# The Biobank Rare Variant consortium powers the discovery of rare genetic associations through global collaboration

**DOI:** 10.64898/2026.05.21.26353759

**Authors:** Duncan S Palmer, Barney Hill, Sam Hodgson, Maarja Jõeloo, Georgios Kalantzis, Athanasios Kousathanas, Satoshi Koyama, Wenhan Lu, Shinichi Namba, Zachary B Rodriguez, Jonathan A Shortt, Kyuto Sonehara, Nicholas Vartanian, Ha My T Vy, Isaac A Wade, Samantha L White, Nikolas A Baya, Nathalie Chami, Ron Do, Karol Estrada, Sarah Finer, Giulio Genovese, Jeremy Guez, Yuval Itan, Masahiro Kanai, Frederik H Lassen, Koichi Matsuda, Loukas Moutsianas, Gina M Peloso, Priit Palta, Daniel J Rader, Augusto Rendon, Ghislain Rocheleau, Omid Sadeghi-Alavijeh, Margaret Sunitha Selvaraj, Roelof AJ Smit, Dapeng Wang, Emilie M Wigdor, Zhi Yu, Colorado Center for Personalized Medicine, Estonian Biobank Research Team, Genes & Health Industry Consortium1, Genes & Health Research Team, Penn Medicine BioBank, The BioBank Japan Project, Christopher R Gignoux, Henrike Heyne, Ruth JF Loos, Hilary C Martin, Lili Milani, Pradeep Natarajan, Yukinori Okada, Nikita Pozdeyev, David A van Heel, Anurag Verma, Wei Zhou, Konrad J Karczewski, Cecilia M Lindgren, Benjamin M Neale

**Affiliations:** Big Data Institute, Li Ka Shing Centre for Health Information and Discovery, University of Oxford, Oxford, UK; Department of Statistics, University of Oxford, Oxford, UK; Program in Medical and Population Genetics, Broad Institute of MIT and Harvard, Cambridge, MA, USA; The Pioneer Centre for SMARTbiomed, Big Data Institute, Li Ka Shing Centre for Health Information and Discovery, University of Oxford, Oxford, UK; Centre for Human Genetics, University of Oxford, Oxford, UK; Wolfson Institute of Population Health, Queen Mary University of London, London, UK; Estonian Genome Centre, Institute of Genomics, University of Tartu, Tartu, Estonia; Wellcome Sanger Institute, Hinxton, UK; Genomics England, London, UK; Personalized Medicine, Mass General Brigham, Boston, MA, USA; Heart and Vascular Institute, Mass General Brigham, Boston, MA, USA; Cardiovascular Disease Initiative, Broad Institute of MIT and Harvard, Cambridge, MA, USA; Department of Medicine, Harvard Medical School, Boston, MA, USA; Analytic and Translational Genetics Unit, Massachusetts General Hospital, Boston, MA, USA; Stanley Center for Psychiatric Research, Broad Institute of MIT and Harvard, Cambridge, MA, USA; Department of Genome Informatics, Graduate School of Medicine, The University of Tokyo, Tokyo, Japan; Laboratory for Systems Genetics, RIKEN Center for Integrative Medical Sciences, Yokohama, Japan; Division of Translational Medicine and Human Genetics, Department of Medicine, Perelman School of Medicine, University of Pennsylvania, Philadelphia, PA, USA; Department of Biomedical Informatics, University of Colorado, Anschutz Medical Campus, Aurora, CO, USA; The Charles Bronfman Institute for Personalized Medicine, Icahn School of Medicine at Mount Sinai, New York, NY, USA; Nuffield Department of Population Health, Medical Sciences Division, University of Oxford, Oxford, UK; Department of Biomedical Informatics, University of Colorado Anschutz, Aurora, CO, USA; Translational Genomics, Maze Therapeutics, South San Francisco, CA, USA; Stanley Center, Broad Institute of MIT and Harvard, MA, USA; Department of Genetics and Genomic Sciences, Icahn School of Medicine at Mount Sinai, New York, NY, USA; Center for Computational and Integrative Biology, Massachusetts General Hospital, Boston, MA, USA; Laboratory of Clinical Genome Sequencing, Department of Computational Biology and Medical Sciences, Graduate School of Frontier Sciences, The University of Tokyo, Tokyo, Japan; Laboratory of Genome Technology, Human Genome Center, Institute of Medical Science, The University of Tokyo, Tokyo, Japan; Department of Biostatistics, Boston University School of Public Health, Boston, MA, USA; Department of Genetics, Perelman School of Medicine, University of Pennsylvania, Philadelphia, PA, USA; Centre for Kidney and Bladder Health, University College London, London, UK; Center for Genomic Medicine, Massachusetts General Hospital, Boston, MA, USA; Cardiovascular Research Center, Massachusetts General Hospital, Boston, MA, USA; Novo Nordisk Foundation Center for Basic Metabolic Research, University of Copenhagen, Copenhagen, Den-mark; National Heart and Lung Institute, Imperial College London, London, UK; Institute of Developmental and Regenerative Medicine, University of Oxford, Oxford, UK; Department of Paediatrics, University of Oxford, Oxford, UK; Clinical and Translational Epidemiology Unit, Massachusetts General Hospital, Boston, MA, USA; University of Colorado Anschutz, Aurora, CO, USA; Colorado Center for Personalized Medicine, University of Colorado Anschutz, Aurora, CO, USA; Human Medical Genetics and Genomics Program, University of Colorado Anschutz, Aurora, CO, USA; University of Colorado Cancer Center, University of Colorado Anschutz, Aurora, CO, USA; Hasso Plattner Institute, Digital Engineering Faculty, University of Potsdam, Potsdam, Germany; Windreich Department of Artificial Intelligence and Human Health, Icahn School of Medicine at Mount Sinai, New York, NY, USA; Finnish Institute for Molecular Medicine, University of Helsinki, Helsinki, Finland; Laboratory of Statistical Immunology, Immunology Frontier Research Center (WPI-IFReC), The University of Osaka, Suita, Japan; Premium Research Institute for Human Metaverse Medicine (WPI-PRIMe), The University of Osaka, Suita, Japan; Division of Endocrinology, Diabetes and Metabolism, University of Colorado Anschutz, Aurora, CO, USA; Blizard Institute, Queen Mary University of London, 4 Newark Street, London, UK; Division of Informatics, Department of Biostatistics, Epidemiology and Informatics, Perelman School of Medicine, University of Pennsylvania, Philadelphia, PA, USA; Psychiatric and Neurodevelopmental Genetics Unit, Center for Genomic Medicine, Massachusetts General Hospital, Boston, MA, USA; Novo Nordisk Foundation Center for Genomic Mechanisms of Disease, Broad Institute of MIT and Harvard, Cambridge, MA, USA; Nuffield Department of Women’s & Reproductive Health, University of Oxford, Oxford, UK

## Abstract

Rare coding variants can have large effects on disease risk and provide direct routes from human genetics to disease mechanisms and therapeutic targets, but their discovery is constrained by sample size, particularly for low-prevalence diseases. Here we establish the Biobank Rare Variant Analysis (BRaVa) consortium, a global rare variant association resource that integrates sequencing and linked health-record data from ten biobanks and cohorts comprising over 1.2 million individuals across diverse ancestries.

We performed gene-based meta-analyses of rare coding variation across 33 clinical endpoints and 11 quantitative traits. Aggregating evidence across biobanks and ancestries identified 514 gene-trait associations, including 31 not previously reported in prior studies or curated association resources following systematic literature review. Notably, 36.1% of gene-level associations were undetectable in any individual biobank, and 91 emerged only through cross-ancestry meta-analysis, demonstrating that federated integration enables discovery beyond the reach of single cohorts. Similar gains were observed at the variant level, where 25.0% of phenotype-locus associations were detectable only through meta-analysis.

Effect size estimates were correlated across ancestries with concordant directions of effect, supporting the generalizability of rare variant associations. The identified signals implicate pathways involved in transcriptional and epigenetic regulation, metabolism, vascular and epithelial biology, and immune function, highlighting rare coding variation as an engine for biological discovery across medical record phenotypes. For example, damaging variation in *ANKRD12* implicates inflammatory transcriptional dysregulation in asthma and chronic obstructive pulmonary disease, and ultra-rare predicted loss-of-function variants in *NAA15* link protein acetylation processes to type 2 diabetes risk.

BRaVa establishes a scalable framework and freely available community resource for rare variant meta-analysis across global biobanks. Public release of gene- and variant-level association summary statistics provides a reference map of rare coding variant associations to support disease gene discovery, biological interpretation, and therapeutic target prioritization as sequencing-linked health-record resources continue to expand.

## Introduction

Understanding how genetic variation contributes to human disease is central to elucidating disease etiology, identifying therapeutic targets, and enabling risk stratification and prevention. Genome-wide association studies (GWAS) have identified thousands of disease-associated loci^1–7^. However, these efforts have primarily focused on common variation and have been disproportionately conducted in populations of European ancestry, limiting biological insight and equitable translation of genetic discoveries^8–10^.

Rare coding variation provides a complementary and often more directly interpretable route to gene discovery, as deleterious variants can exert larger effects and directly implicate causal genes^11,12^. Advances in sequencing technologies and the expansion of large-scale biobanks have increased sample sizes and improved rare variant detection, enabling more powerful association studies across a broader range of traits^13–16^. Nonetheless, discovery of rare variant associations (particularly for binary disease endpoints and less prevalent conditions) remains challenging within individual cohorts due to limited sample sizes and the inherent sparsity of rare variation.

Large biobanks increasingly combine population-scale sequencing with digitized longitudinal health information, creating powerful resources for rare variant association studies across broad medical phenotypes^13,17,18^. Meta-analysis across multiple biobanks offers a scalable strategy to address the remaining limitations by increasing effective sample size, expanding the spectrum of rare variation interrogated, and enabling discovery beyond the reach of any single study^19^. While large sequencing resources such as the UK Biobank and All of Us have demonstrated the value of population-scale rare variant analyses^13,16–18^, no single biobank captures the full diversity of human populations, disease architectures, or environmental contexts^20^. Integrating data across biobanks offers a path toward addressing these gaps by increasing sample diversity, improving statistical power, and enabling broader representation across ancestries and environments. However, coordinated analysis across biobanks requires harmonization of phenotypes, sequencing data, and governance frameworks, which can complicate data sharing and joint analysis. Collaborative approaches are therefore essential for comprehensive rare variant discovery.

Here, we present BRaVa (**B**iobank **Ra**re **Va**riant Analysis), a global consortium integrating rare variant association results across ten large biobanks spanning globally diverse populations, sequencing designs, and patient recruitment strategies. By aggregating rare variant association evidence, BRaVa enables large-scale discovery of rare coding variant associations across diseases and quantitative traits. This approach substantially increases power relative to individual biobanks and facilitates systematic evaluation of rare variant associations at global scale. We analyze rare coding variation across 33 clinical endpoints and 11 biomarkers and continuous traits, leveraging sequencing data from ∼ 1.2 million individuals. By integrating evidence across biobanks and genetic ancestry groupings, BRaVa establishes a scalable community resource, with public summary statistics to support disease gene discovery, evaluation of rare variant effect-size generalizability across populations, and prioritization of biologically grounded therapeutic targets.

## Results

### Large-scale rare-variant meta-analyses across global biobanks

BRaVa integrates sequencing data across multiple globally distributed biobanks (Extended Data Fig. 1). In our first major collaboration, we conducted rare variant and gene-based meta-analyses across 33 clinical endpoints and 11 biomarkers and continuous traits (Tables S1-S2, Fig. 1) in up to ten biobanks^6,13,14,17,21–27^. Details of the contributing biobanks, including geographic location, sample size, recruitment strategy, broad-scale ancestry composition and sequencing modalities, are provided in Table S3 and Methods: Biobank descriptions.

**Figure 1:**
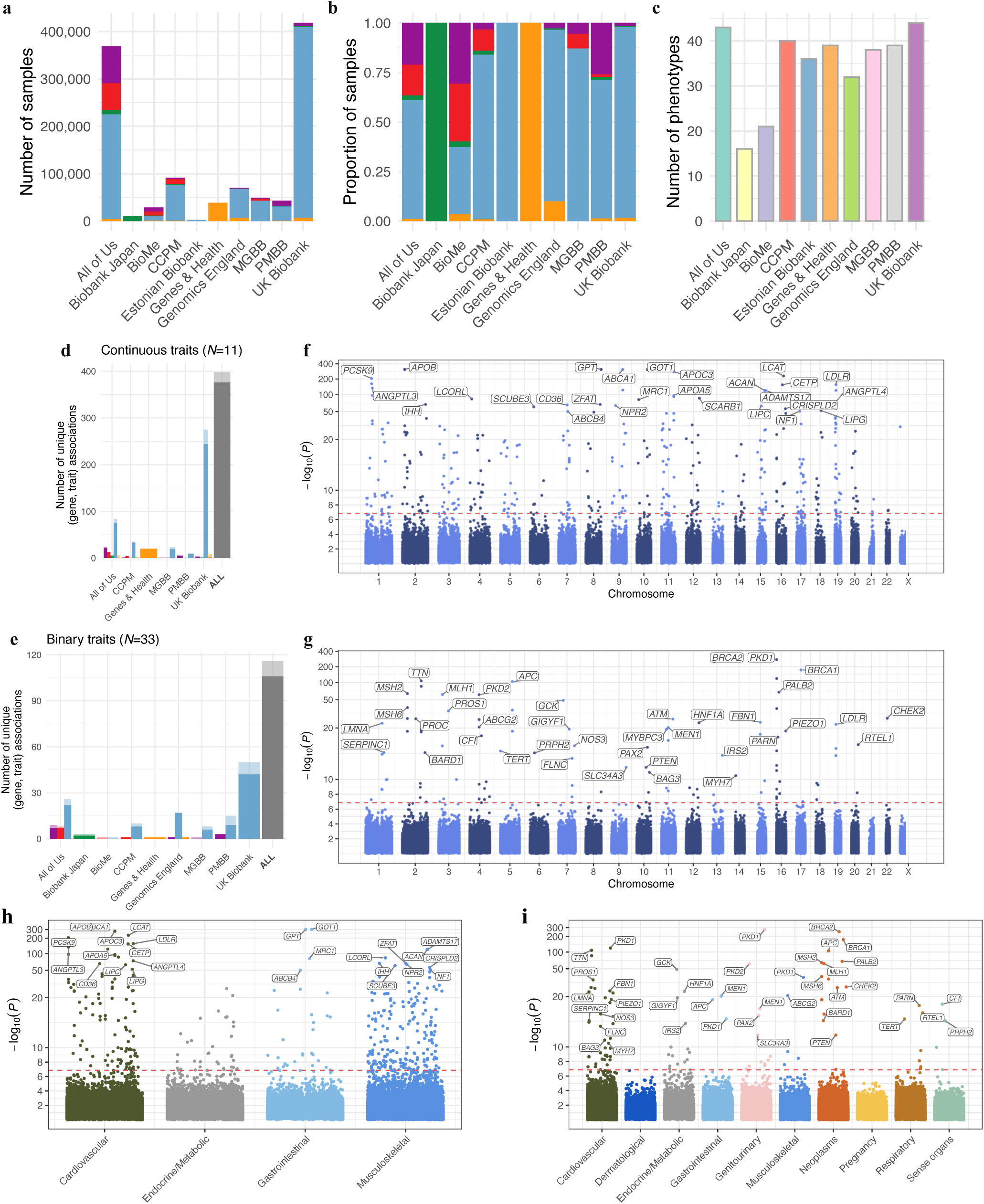
Meta-analysis of ∼1.2 million individuals across ten global biobanks: gene-level. a, Stacked bar plots showing counts and, b, proportions of individuals by broad genetic ancestry. c, Counts of available phenotypes per biobank. Phenotypes were included for a given (biobank, genetic ancestry) tuple when both case and control counts exceeded 100. d-e, Counts of unique gene-trait associations identified within individual biobanks and the overall meta-analysis for continuous and binary traits, respectively. Dark bars denote gene-trait associations for which at least one of 18 tests passed the per-mask significance threshold (*P* < 1.39 × 10^−7^); lighter bars denote associations significant after aggregation (Cauchy *P* < 2.5 × 10^−6^). f-g, Summaries of gene-based association results for continuous and binary traits, respectively. For each gene-trait pair, we report the most significant result across test type (burden, SKAT, SKAT-O), MAF cutoff ∈ 0.1%, 0.01%, and annotation ∈ pLoF, damaging missense or protein altering, pLoF or damaging missense or protein altering. Red dotted lines indicate the experiment-wise significance threshold for gene masks (*P* < 1.39 × 10^−7^). h-i, Corresponding plots stratified by ICD chapter. Equivalent analyses restricted to broad genetic ancestry subsets and non-European groups are shown in Figs. S4-S5.

To summarize the genetic ancestries represented within BRaVa, we projected participants onto the principal component space defined by the 1000 Genomes^28^ (1000G) and Human Genome Diversity Project^29^ (HGDP) reference panels (Extended Data Fig. 2, Figs. S1-S2). Following the approach used in the Global Biobank Meta-analysis Initiative^20^ (GBMI), individuals were assigned to broad continental ancestry groups using these reference datasets: African (AFR), admixed American (AMR), East Asian (EAS), European (EUR), and Central and South Asian (SAS). As in prior large-scale genetic studies, these groupings were used as an analytic strategy to improve calibration and reduce potential confounding due to population stratification in rare variant association analyses, and should not be interpreted as biologically discrete or socially meaningful categories. Association analyses were performed within ancestry- and biobank-specific subsets prior to meta-analysis. We refer to these hereafter using three-letter codes, following prior work. BRaVa is not yet globally representative (Tables S4-S8); most individuals assigned to AMR and AFR groups were recruited through US-based biobanks, whereas most assigned to SAS were recruited through UK-based cohorts. Continental African and Central/South American populations remain underrepresented. Despite these limitations, BRaVa provides one of the broadest cross-ancestry resources currently available for rare variant association analyses. Across all contributing biobanks, BRaVa includes approximately 112,000 individuals assigned to AFR, 80,000 to AMR, 25,000 to EAS, 844,000 to EUR, and 59,000 to SAS.

Disease prevalence varied by biobank and recruitment strategy (Table S9, Fig. S3). As in GBMI, biobanks recruiting from hospitals displayed increased prevalence for a subset of diseases as compared to biobanks recruiting participants at the population-level (Wilcoxon signed rank test *P* < 0.05 for 14 out of the 33 examined diseases and procedures)^20^.

Phenotypes were nominated for analysis by members of the consortium, and harmonized based on ICD and SNOMED mappings^30–33^ (Methods: Phenotype definitions). Phenotype definitions were shared across the consortium to harmonize phenotype curation (Table S1-S2). A subset of nominated phenotypes was selected for analysis based on representation across biobank–ancestry subcohorts. Specifically, we retained disease traits with at least 100 cases in at least ten biobank–ancestry subcohorts spanning at least five biobanks, or with a case prevalence >1% across ancestries in UK Biobank (Methods: Phenotype definitions; Table S6-S7). To ensure the quality and reliability of association results, we also conducted stringent sample curation and quality control of genetic data, following collaboratively defined principles (Methods: Quality control guidance for sequencing data).

### Harmonized rare-variant association testing framework

We implemented a harmonized, multi-ancestry analysis workflow for genome- and exome-sequencing data^34^, performing association analyses within ancestry- and biobank-specific subsets prior to meta-analysis to improve calibration and reduce potential confounding due to population stratification, which is particularly important for rare variant analyses. Scalable generalized mixed models implemented in SAIGE^35^ and SAIGE-GENE+^36,37^ were used to account for sample relatedness and case-control imbalance^35–39^.

To increase power in gene-level rare variant association testing at maximum minor allele frequencies (MAF) of 0.1%, and 0.01%, we aggregate putatively deleterious variants within genes using functional annotation masks: we consider predicted loss-of-function and damaging missense or protein-altering variants^11,40–43^ (Methods: Annotation definitions for gene-level association testing, Association testing).

We used synonymous gene-based tests as an internal calibration control, as synonymous variants are not expected to be enriched for association signals. As such, inflation in these statistics flags residual confounding, technical artifacts, or phenotype imbalance rather than true genetic signal. We quantified inflation within each (biobank, genetic ancestry) for each phenotype using upper-tail genomic inflation metrics which are sensitive to tail inflation in rare variant tests where median-based metrics can be deflated, particularly for binary traits with low case counts. We excluded (biobank, genetic ancestry) strata exceeding predefined thresholds (*λ*_95_, *λ*_99_, *λ*_99.9_ > 1.3), and meta-analyzed well-calibrated results (Methods: Summary statistic quality control; Fig. S6, Table S10). Across synonymous gene-based tests, median *λ*_95_ was 0.97 (95% quantiles [0.86, 1.05]), and 6/834 (0.72%) of strata were excluded by this filtering procedure.

Age-related somatic mutations driving clonal hematopoiesis (CHIP) can confound rare germline variant signals. To increase robustness in interpretation of germline effects, we evaluated the major canonical CHIP genes (*DNMT3A*, *TET2*, and *ASXL1*; Fig. S7) but excluded them from downstream analyses and figures. Other CHIP-associated genes were retained to preserve potential true germline associations.

After restricting to well-calibrated association results within (biobank, genetic ancestry) pairs (Fig. S6), we compared gene-level effect estimates across genetic ancestries (Fig. S8, Table S11). As observed previously^19,44–46^, effect size estimates were strongly correlated across ancestries: estimates from individuals of European genetic ancestry were significantly correlated with those from all non-European ancestry groups (Bonferroni-corrected *P* < 0.001 [0.05/48] across 48 tests spanning combinations of ancestry group, phenotype class (binary/continuous), variant annotation mask, and MAF cutoff; see Methods: Cross-ancestry comparison of gene-level burden effect sizes; Table S11). Further, among gene-phenotype associations with gene-burden *P* < 1.0 × 10^−4^ in both ancestry groups being compared, directions of effect were uniformly concordant.

Robust genetic association results despite differences in genetic ancestry and recruitment across biobanks motivated the use of fixed-effects meta-analysis across all (biobank, genetic ancestry) pairs for each analyzed phenotype (Extended Data Fig. 3). We used inverse-variance weighted meta-analysis for variant and gene-level burden test statistics (though inverse-variance and Stouffer’s method^47^ gave highly concordant results (Fig. S9)), and Stouffer’s method^47^ to meta-analyze variance (SKAT^48^) and hybrid (SKAT-O^49^) -based gene-level test statistics in up to ten biobanks (Methods: Meta-analysis). The resultant distribution of genomic inflation of meta-analyzed test statistics were well calibrated (Fig. S10). Experiment-wide significance thresholds were defined separately for gene-level, gene-mask-level, and variant-level analyses. For gene-level analyses, we used a threshold of Cauchy *P* < 0.05/20, 000 = 2.5 × 10^−6^, accounting for approximately 20,000 genes tested across 18 correlated masks of putatively damaging rare variation (Extended Data Fig. 3). For gene-mask-level analyses, we applied a Bonferroni threshold of *P* < 0.05/(20, 000 × 18) = 1.39 × 10^−7^. For variant-level analyses, significance was defined as *P* < 0.05/2, 746, 957 = 1.82 × 10^−8^, where 2,746,957 represents the maximum number of variants tested for any trait.

Following the initial meta-analysis, we evaluated the extent to which sample overlap within and between biobanks, and tagging of rare variation by nearby common variants, could inflate false discovery rates. First, to assess the impact of relatedness within cohorts, we compared meta-analysis results based on Stouffer’s method using weights assuming unrelated samples to those incorporating effective sample size estimates derived from genetic relatedness (Methods: Meta-analysis). Association results were highly consistent between approaches, likely reflecting the limited degree of relatedness in the largest contributing cohorts, including UKB^13^ and AoU^18^ (Fig. S11). Then, as a sensitivity analysis, we repeated gene-level meta-analysis using a cumulative allele frequency (CAF)-weighted version of Stouffer’s method to account for differences in the aggregate contribution of rare variants within each gene across (biobank, ancestry) pairs (Methods: Cumulative allele frequency weighted Stouffer’s meta-analysis for SKAT and SKAT-O). Results were highly concordant with the primary analysis (Fig. S12), indicating that our findings are robust to alternative weighting schemes.

Finally, we sought to determine the potential impact of sample overlap between cohorts on our findings. Existing overlap-correction methods that operate on summary statistics were developed for common variant analyses^50,51^ but they can yield biased results when applied to rare variant association tests (Methods: Caveats of METAL sample overlap correction for rare variant analyses; Figs. S13-S15). To guard against this, we adopted an empirical strategy to estimate sample overlap based on correlations of gene-level association statistics derived from synonymous variation (30 < MAC < 1000) under the 0.1% MAF mask. Correlations were incorporated into the overlap correction only when effect sizes were positively correlated (*P* < 0.01; Figs. S16-S20). Synonymous burden statistics provide an approximately null signal, so correlations in gene-level effects of synonymous variant burden across cohorts primarily reflect sample overlap rather than shared genetic architecture. Restricting to positive correlations avoids spurious negative estimates driven by noise, which would otherwise imply implausible ‘anti-overlap’ and bias the correction. These correlation estimates were then used as weights in a METAL-style overlap correction adapted for gene-based meta-analysis (Fig. S21). We note that the METAL overlap correction weights are not optimal^52,53^ (see Methods: Accounting for overlap in fixed effects meta-analysis for details), but have been shown to perform well in practice^54^. Following adjustment for sample overlap between cohorts, we saw slight attenuation of signal, as expected, but retained 96.9% of significant associations in the primary analysis (Cauchy *P* < 2.5 × 10^−6^).

### Increased discovery through cross-ancestry meta-analysis

Aggregating ten biobanks in BRaVa substantially increased sample sizes for genetic association studies in the analyzed traits, leading to an increase in power for genetic discovery. Meta-analysis across all biobank-ancestry pairs identified 514 unique gene-phenotype associations involving putatively deleterious variation (Fig. 1). Of these, 186 (36.1%) were not significant in any individual biobank (Fig. 4a, Figs. S22-S33, Tables S12-S15). Meta-analyses restricted to broad continental ancestries are shown in Figs. S4-S5.

Considering meta-analysis at the level of broad continental genetic ancestry, the combined analysis across all (biobank, ancestry) pairs identified the largest number of associations (Fig. 2a), totaling 514. Of these, 91 were detected only in the cross-ancestry meta-analysis and were not significant in any single-ancestry meta-analysis, indicating that increased aggregate sample size and cross-ancestry integration materially enhance discovery. Restricting the analysis to the two largest resources (UKB and AoU) reproduced most associations; however, 18.9% (97/514, among which 93 were nominally significant [Cauchy *P* < 0.05] in at least two biobanks) were not captured (Fig. 2b, Fig. S34), demonstrating the additional gain achieved through federation across the full set of BRaVa cohorts.

**Figure 2:**
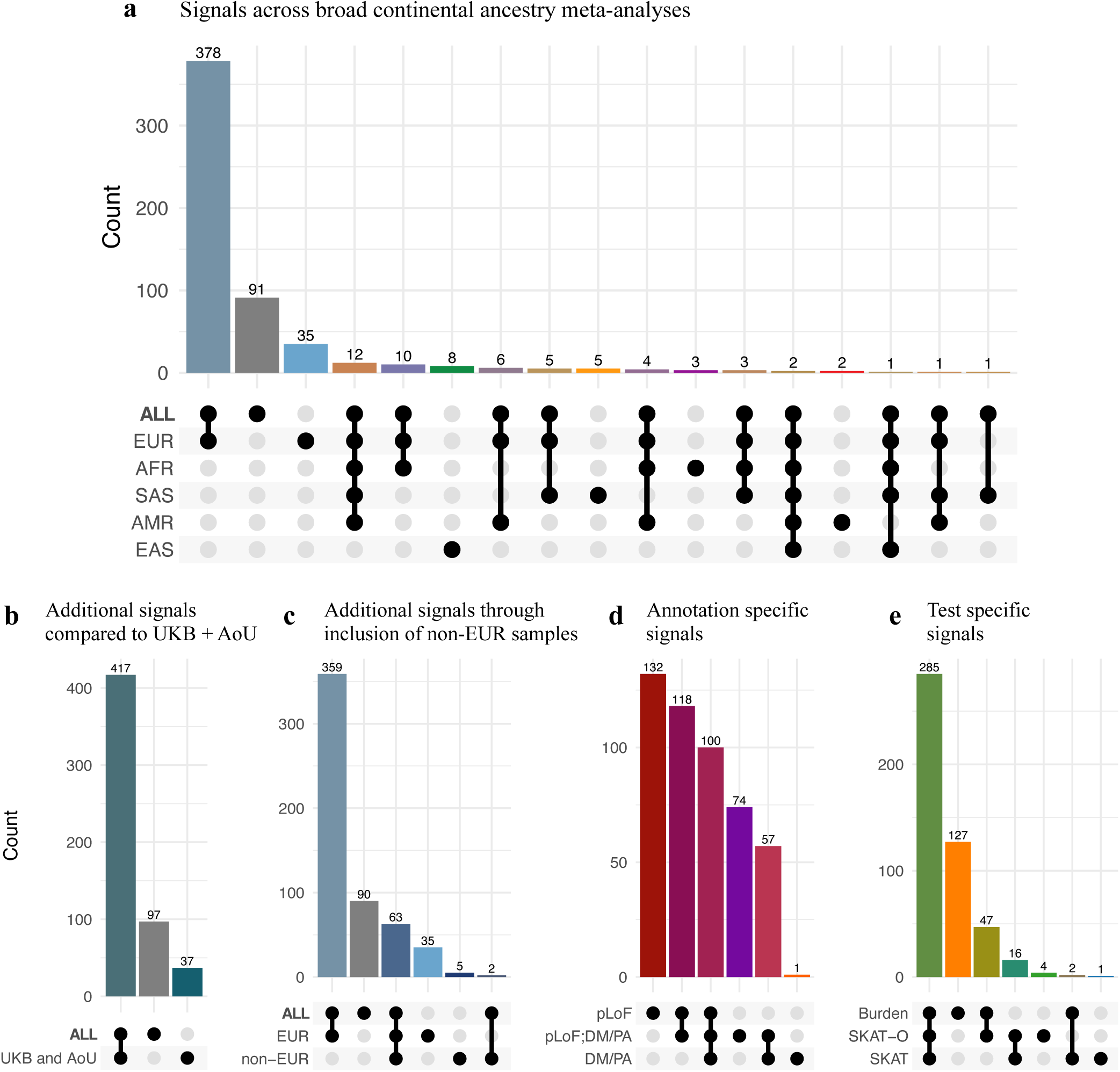
Summary of unique gene-phenotype associations. a, Upset plot of unique experiment-wise significant genephenotype associations (Cauchy *P* = 2.5 × 10^−6^), split by meta-analysis of results within genetic ancestry. ‘ALL’ comprises results after meta-analyzing across all (biobank, genetic ancestry) pairs. Areas within inset Venn diagrams are scaled according to the count of unique gene-trait associations. b, Corresponding upset plots with inset Venn diagrams showing counts of unique gene-trait associations comparing meta-analysis of all (biobank, ancestry) pairs with (biobank, ancestry) pairs restricted to UKB and AoU. c, as in b, showing counts of unique gene-trait associations unique to and shared between meta-analysis results across all (biobank, ancestry) pairs, and those restricted to European and non-European ancestry labels. d, Upset plot with inset Venn diagram, displaying the breakdown of experiment-wise significant gene-mask associations (*P* < 1.4 × 10^−7^) split by annotation mask (pLoF = predicted loss of function, DM/PA = damaging missense or protein altering, pLoF; DM/PA = union of predicted loss of function and damaging missense or protein altering). e, Upset plot with inset Venn diagram, displaying the breakdown of experiment-wise significant gene-mask associations (*P* < 1.4 × 10^−7^) split by class of test.

Ancestry composition and mask/test choice also influenced discovery (Fig. 2c-e). Inclusion of non-European samples contributed associations not observed in European-only meta-analyses (Fig. 2c), demonstrating a measurable gain from broader ancestry representation, largely reflecting increased aggregate sample size. Predicted loss-of-function (pLoF) masks accounted for the largest share of associations, whereas damaging missense/protein-altering (DM/PA) and union masks identified partially overlapping but distinct sets of genes (Fig. 2d). The broader union mask recovered additional associations beyond pLoF alone, consistent with increased sensitivity at a less stringent ‘deleteriousness’ threshold. Although directional burden tests captured the majority of signals, variance-component approaches (SKAT and SKAT-O) identified a smaller number of additional associations (Fig. 2e), consistent with allelic heterogeneity and non-directional effects at a subset of loci. The predominance of pLoF-driven discoveries likely reflects the clearer functional consequences of truncating variation, whereas partially overlapping DM/PA and union-mask signals highlight both the value and remaining difficulty of modelling damaging missense variation. Together, these results indicate that gene-level discovery benefits from combined scale, ancestry diversity, and complementary annotation and testing strategies.

At the variant level (MAF < 0.1%), we identified 371 significant phenotype-variant associations (*P* < 1.8 × 10^−8^), at 128 unique phenotype-locus pairs (Methods: Locus definition for variant-level results, Fig. 3, Tables S16-S17). Of these 371 phenotype-variant associations, 68 (18.3%), spanning 32 phenotype-locus pairs, were not significant in any individual (biobank, genetic ancestry) analysis. The larger relative increase in discoveries at the gene level highlights the additional power gained by aggregating rare variants according to predicted deleteriousness. Broad continental ancestry specific variant meta-analysis summaries are shown in Figs. S35-S40.

**Figure 3:**
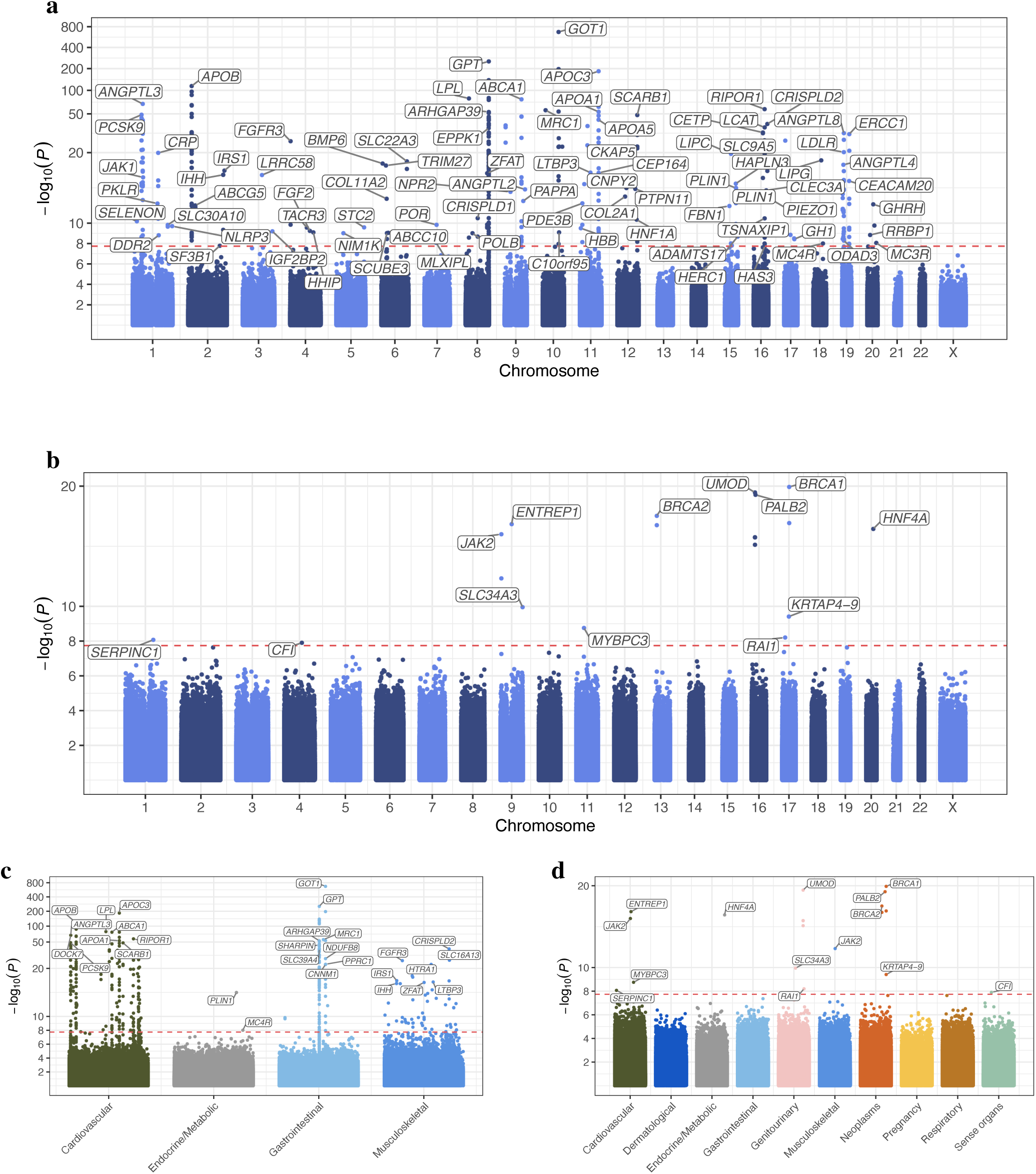
Meta-analysis of ∼1.2 million individuals across ten global biobanks: variant level, MAF. < **0.1%**. a-b, Summary of variant level analysis results for continuous and binary traits, respectively. For genes with multiple significant associations, only the most significant (lowest P value) is labeled; genes without an assigned symbol are not labeled. The red dotted horizontal line denotes the experiment-wise significance threshold for variant-level association testing (*P* = 1.8 × 10^−8^). c-d, display the corresponding plots, split by ICD chapter on the *x*-axis. Plots restricted to meta-analysis within broad genetic ancestry subsets, and across non-European broad ancestry groups are displayed in Figs. S35-S40.

To assess whether gene-level rare variant associations are independent of nearby common variant signals, or instead reflect tagging of common GWAS loci, we performed iterative conditioning on common variant associations within 500 kb of each associated gene within (biobank, genetic ancestry) subcohorts. Conditioning was carried out independently within each ancestry, and the resulting sets of significant variants were combined across biobanks by taking their union before meta-analyzing as before (Methods: Common variant conditioning; Figs. S41-S43).

Results were largely consistent with those found without iterative conditioning on common variants, with 1.2% of gene-mask-level significant (*P* < 1.39 × 10^−7^) associations dropping below significance thresholds (Fig. S44), indicating that the majority of rare variant signals could not be explained by nearby common variant associations.

### Rare variant meta-analysis reveals known and novel gene-level associations across diverse phenotypes

Across binary and quantitative traits, cross-biobank rare variant meta-analysis identified gene-level associations that were not detectable within individual cohorts, spanning cardio-metabolic, vascular, immune, proliferative, lipid, and anthropometric phenotypes. These findings highlight rare coding contributions that emerge only through large-scale aggregation. To systematically assess the novelty of these discoveries, we used an AI-based agent to evaluate evidence from both biomedical literature and the Open Targets platform^55^ (Methods: Novelty assessment of gene-phenotype associations; Fig. S45), followed by manual review. This informed the selection of highlighted associations, chosen based on biological relevance or novelty from among significant results (Table S12-S17).

We recapitulated several associations recently reported by Jurgens *et al*.^19^, including rare deleterious variation in *MIB1* with type 2 diabetes (T2D) (pLoF or DM/PA, MAF < 0.1%, burden *P* = 5.62×10^−10^; Cauchy *P* = 5.70×10^−9^, Fig. 4c) and in *UBR3* associated with both hypertension (pLoF, MAF < 0.1%, burden *P* = 3.44 × 10^−10^; Cauchy *P* = 3.87 × 10^−9^) and T2D (pLoF, MAF < 0.1%, burden *P* = 1.1 × 10^−9^; Cauchy *P* = 1.10 × 10^−8^, Fig. 4c). We extend these findings by identifying additional associations of rare putatively damaging variation in *MIB1* with waist-to-hip ratio adjusted for body mass index (BMI; WHRadjBMI) (pLoF, MAF < 0.1%, burden *P* = 1.22 × 10^−9^; Cauchy *P* = 1.75 × 10^−8^), and HDL cholesterol (pLoF, MAF < 0.01%, burden *P* = 2.05 × 10^−7^; Cauchy *P* = 1.34 × 10^−6^), suggesting broader pleiotropic effects of MIB1 across metabolic phenotypes. In addition, we identify an association between the *UBR3* paralog *UBR2* and BMI (pLoF, MAF < 0.01%, burden *P* = 7.36 × 10^−9^; Cauchy *P* = 9.59 × 10^−8^), consistent with prior reports^56,57^.

**Figure 4:**
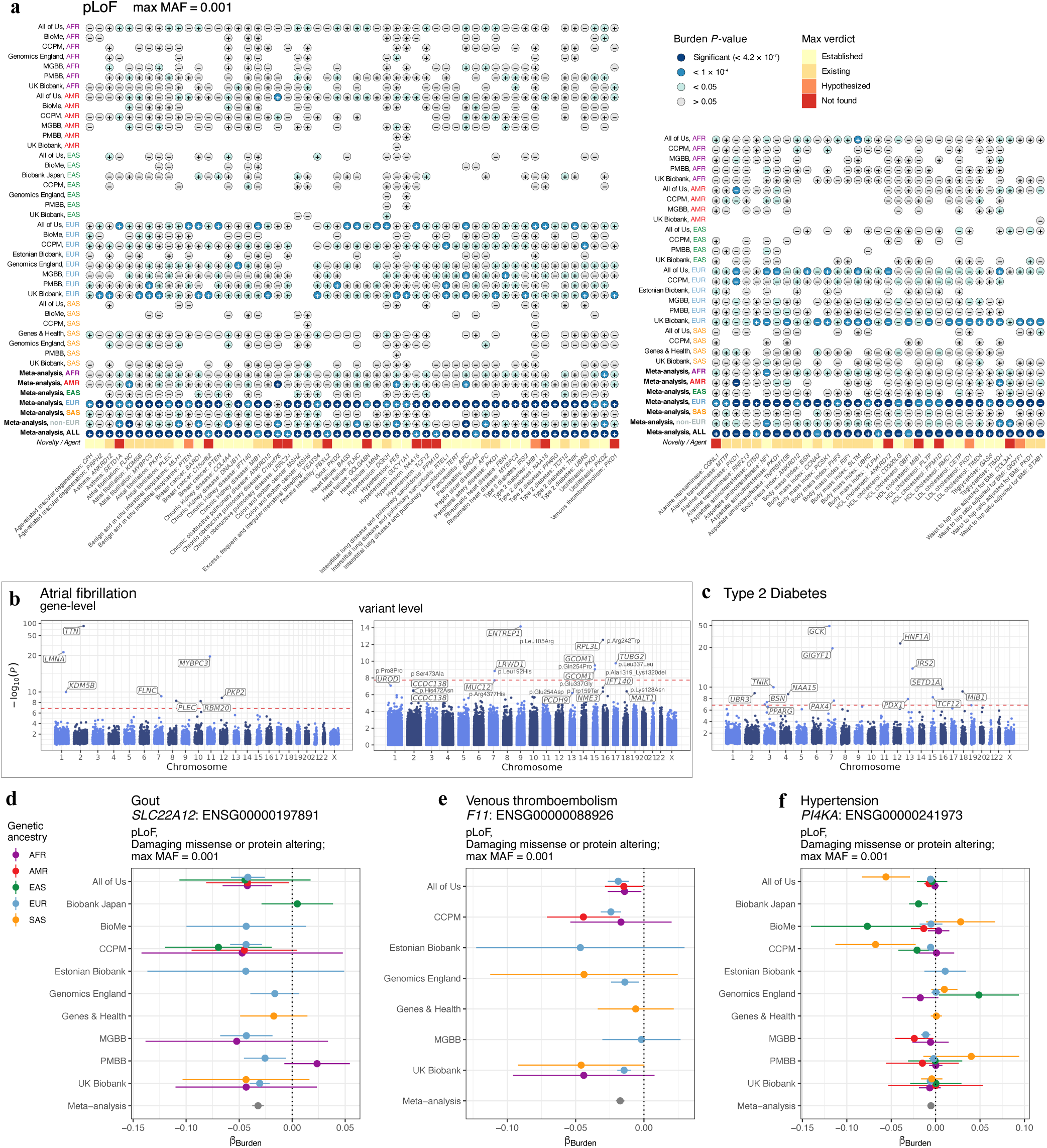
Federated meta-analysis reveals gene-level associations absent in individual biobank–ancestry analyses, demonstrates substantial power gains, and highlights protective effects of rare damaging variants. a, Gene-level burden associations for pLoF variants (MAF < 0.1%) that are significant in meta-analysis but not in any individual (biobank, ancestry) analysis. The left plot displays results for binary traits, and the right for continuous traits. Circles correspond to results for each (biobank, ancestry) combination or meta-analysis (*y*-axis), and are colored by significance level as indicated in the legend. The symbol within each circle denotes the direction of effect (positive or negative *β*_Burden_). The bottom row summarizes prior evidence for each association using classifications from an AI-based agent (Lu *et al.*, 2026), ranging from not found to established. b-c, Manhattan-style plots demonstrating the gain in power from federated meta-analysis. b, Atrial fibrillation: gene-level (left) and variant-level (right) association results. c, T2D: gene-level results. Horizontal dashed lines indicating the significance threshold for gene-mask testing (*P* < 1.4 × 10^−7^). d-f, forest plots of gene-level burden associations across cohorts and ancestries for selected examples showing protective effects of burden of pLoF and damaging missense or protein altering variation (MAF < 0.1%) against disease. Points represent burden effect-size estimates (*β*_Burden_) with standard errors across individual biobanks and meta-analysis (gray), colored by genetic ancestry. d, *SLC22A12* (gout), an established therapeutic target for urate lowering. e, *F11* (venous thromboembolism), a target of emerging anticoagulant therapies, and f, *PI4KA* (hypertension).

We also observed associations with *YEATS4* that align with prior reports linking inherited variation in this gene to uterine phenotypes^58^. Although we did not recapitulate the reported association with benign neoplasms of the uterus^19^, we detected an association with excess menstrual bleeding (pLoF, MAF < 0.1%, burden *P* = 1.14 × 10^−8^; Cauchy *P* = 2.05 × 10^−7^), suggesting potentially related reproductive effects.

Rare variant- and gene-level associations for atrial fibrillation reported by Choi *et al*.^59^ were consistent with our findings (Fig. 4b). All five reported genes met our significance threshold with concordant effect directions, and stronger statistical support in the present study consistent with increased sample size. Similarly, we recapitulated three of the five variant level associations with increased statistical support (Fig. 4b). The remaining two reported associations were not replicated in Choi *et al*.^59^ nor driven by a coding variant unique to Asian ancestries with relatively limited sample size across the BRaVa cohorts (chr10:103611618:A:C, AC = 176, *P* = 0.08) and inconsistent direction of effects between EAS (BBJ and AoU), and SAS (G&H) cohorts.

Rare coding variation in *KEAP1*, a regulator of NRF2-mediated oxidative stress responses^60^, was associated with HDL cholesterol (pLoF or DM/PA, MAF < 0.1%, burden *P* = 4.77 × 10^−8^; Cauchy *P* = 7.97 × 10^−7^). Although *KEAP1* has previously been implicated in adiposity-related traits, including fat distribution^61^ and WHRadjBMI^62^, which we also observe (pLoF or DM/PA, MAF < 0.1%, burden *P* = 1.73 × 10^−12^; Cauchy *P* = 3.07 × 10^−11^), our findings extend its metabolic associations to circulating lipid levels, suggesting a broader role for redox regulation in metabolic homeostasis.

Anthropometric traits further implicated pathways involved in protein translation and growth signaling. Rare pLoF variants in *GIGYF1* were associated with WHRadjBMI (pLoF, MAF < 0.1%, burden *P* = 9.28 × 10^−9^; Cauchy *P* = 1.63 × 10^−7^), connecting insulin/IGF signaling and regulation of protein translation to body fat distribution independent of overall adiposity. Previous exome sequencing studies have reported male-specific associations between *GIGYF1* and WHRadjBMI^15,62^, with cross-sex associations approaching exome-wide significance^63^; our results further support a role for this pathway in body fat distribution.

Beyond *GIGYF1*, our rare variant meta-analysis highlighted likely causal genes at several anthropometric loci previously implicated primarily by common variants. For example, rare coding variation in *LCORL* was significantly associated with height (pLoF, MAF < 0.1%, SKAT *P* = 5.23 × 10^−84^; Cauchy *P* = 7.28 × 10^−83^). While GWAS have strongly linked this locus to human stature, our rare variant evidence provides strong human genetic support that aligns with compelling cross-species genetic evidence, including a nonsynonymous variant affecting body frame size in cattle^64,65^ and an eQTL explaining approximately 20% of height variation in horses^66,67^.

Similarly, for BMI, we identified associations for *BSN* and *CALCR*. The associations at *BSN* (BMI: pLoF, MAF < 0.1%, burden *P* = 1.13 × 10^−11^; Cauchy *P* = 1.97 × 10^−10^, T2D: pLoF, MAF < 0.1%, burden *P* = 4.18 × 10^−8^; Cauchy *P* = 7.43 × 10^−7^) support recent findings linking protein-truncating variants in this gene to severe adult-onset obesity and pleiotropic metabolic complications, including T2D and fatty liver disease^68^. Furthermore, the *CALCR* association (pLoF or DM/PA, MAF < 0.1%, burden *P* = 4.68 × 10^−8^; Cauchy *P* = 8.35 × 10^−7^) is particularly notable as it helps resolve a known GWAS locus to *CALCR*, which encodes an established drug target.

This finding complements murine neurocircuitry and human transcriptomic studies highlighting the critical role of *CALCR*-expressing neurons in regulating energy balance and appetite^69,70^.

A strong vascular association was observed for *PKD1*, where rare pLoF variants were associated with venous thromboembolism (pLoF, MAF < 0.1%, burden *P* = 3.10 × 10^−11^; Cauchy *P* = 3.55 × 10^−10^). *PKD1* encodes polycystin-1, a protein central to renal and vascular biology^71,72^. Mutations in *PKD1* are the predominant cause of autosomal dominant polycystic kidney disease. This finding potentially extends polycystin-mediated signaling into thrombotic disease susceptibility.

Among cardiometabolic outcomes, rare pLoF variation in *NAA15* was associated with T2D (pLoF, MAF < 0.1%, burden *P* = 1.63 × 10^−9^; Cauchy *P* = 2.09 × 10^−8^) and hypertension (pLoF, MAF < 0.1%, SKAT *P* = 5.17 × 10^−8^; Cauchy *P* = 2.33 × 10^−7^). *NAA15* encodes a core component of the N-terminal acetyltransferase complex^73^, implicating co-translational protein modification and proteostasis in metabolic and blood-pressure regulation. Rare pLoF variation in *TCF12*, a basic helix-loop-helix transcription factor, was similarly associated with T2D (pLoF, MAF < 0.1%, burden *P* = 5.73 × 10^−9^; Cauchy *P* = 5.59 × 10^−8^) and hypertension (pLoF, MAF < 0.1%, burden *P* = 5.98×10^−9^; Cauchy *P* = 8.11×10^−8^). Multiple GWAS (in up to → 2.5 million individuals) report genome-wide significant associations with T2D and glaucoma at the *TCF12* locus^5,20,55,74,75^.

Liver biomarker analyses identified novel signals at two paralogous tight-junction scaffold genes. Rare coding variation in *CGN* and *CGNL1* was associated with alanine transaminase (*CGN*: pLoF, MAF < 0.1%, burden *P* = 1.70 × 10^−7^; Cauchy *P* = 2.87 × 10^−6^, *CGNL1*: pLoF, MAF < 0.1%, burden *P* = 5.33 × 10^−8^; Cauchy *P* = 9.08 × 10^−7^). Both genes, which sit on different chromosomes, have been found to be associated with *γ*-glutamyl transferase^15^, another liver biomarker, indicating a consistent relationship with markers of liver function.

*CGN* and *CGNL1* encode structurally related proteins involved in epithelial junctional organization and cytoskeletal regulation. Notably, *CGN* and *CGNL1* are highly and moderately expressed in liver^76^, respectively, and common (MAF *>* 1%) intronic variants near *CGNL1* have shown suggestive associations with alcohol consumption in AFR ancestry-specific GWAS^4^. The independent associations at two related genes across two liver biomarkers support a role for junctional and cytoskeletal pathways in liver function.

Burden of rare pLoF variants (MAF < 0.1%) in *PDZK1* was nominally associated with increased HDL cholesterol in European genetic ancestry participants (burden *P* = 5.03 × 10^−7^). *PDZK1* encodes a scaffolding protein required for hepatic expression of the HDL receptor SR-BI (*SCARB1*), and PdzK1 knockout mice exhibit elevated circulating HDL levels due to impaired hepatic HDL uptake^77^. Although extensive mechanistic and animal model evidence supports a central role for *PDZK1* in HDL regulation^78,79^, human genetic support has been limited. The positive association observed here is consistent with the phenotype observed in PdzK1 knockout models and provides direct human genetic evidence implicating *PDZK1* in plasma HDL cholesterol homeostasis.

Immune-mediated phenotypes highlighted several genes with roles in chromatin modification and transcriptional regulation. Rare pLoF variants in *SETD1A* were associated with asthma (pLoF, MAF < 0.1%, burden *P* = 8.15 × 10^−9^; Cauchy *P* = 9.61 × 10^−8^). *SETD1A* encodes a core H3K4 methyltransferase required for promoter activation and transcriptional regulation. Given the central role of epigenetic regulation in T-cell differentiation, cytokine expression, and airway epithelial homeostasis^80,81^, these findings implicate *SETD1A* in asthma susceptibility and suggest that chromatin regulation may contribute to disease risk. Notably, *SETD1A* is also strongly implicated in schizophrenia through rare coding variation, suggesting that disruption of chromatin regulatory pathways may have pleiotropic effects across neuropsychiatric and immune-mediated phenotypes. Similarly, rare pLoF burden in *ANKRD12*, a nuclear receptor-interacting transcriptional co-regulator, was associated with asthma (pLoF, MAF < 0.1%, burden *P* = 5.28 × 10^−8^; Cauchy *P* = 9.12 × 10^−7^) and chronic obstructive pulmonary disease (pLoF, MAF < 0.1%, burden *P* = 3.24 × 10^−10^; Cauchy *P* = 3.55 × 10^−9^), consistent with a broader role for altered transcriptional and epigenetic regulation, which may drive lung inflammation through dysregulated immune gene expression.

Among the experiment-wise significant gene-level burden results (Bonferroni *P* < 4.2 × 10^−7^), putatively damaging coding variation in three genes was associated with reduced disease prevalence (Fig. 4d–f, Table S13, S17). We identified two established protective associations that serve as positive controls. Rare deleterious variation in *SLC22A12*, which encodes the renal urate transporter URAT1, was associated with reduced risk of gout (pLoF or DM/PA, MAF < 0.1%, burden *P* = 3.60 × 10^−9^), consistent with the mechanism of uricosuric therapies that lower serum urate by inhibiting URAT1^82–84^. Similarly, rare deleterious variation in *F11*, encoding coagulation factor XI, was associated with reduced risk of venous thromboembolism (pLoF or DM/PA, MAF < 0.1%, burden *P* = 2.14 × 10^−8^), in line with the emerging use of factor XI inhibitors as anticoagulants^85,86^. In addition, rare putatively deleterious genetic variation in *PI4KA* was also associated with reduced risk of hypertension (pLoF or DM/PA, MAF < 0.1%, burden *P* = 2.98 × 10^−7^), representing a novel protective signal for blood pressure regulation. Given the established role of PI4KA in maintaining the plasma membrane phosphoinositide pools required for G-protein-coupled receptor signaling^87^, and experimental evidence that PI4KA inhibition leads to severe in vivo toxicity, including sudden death consistent with cardiovascular collapse^88^, these findings suggest that reduced PI4KA activity may influence pathways relevant to blood pressure regulation. Overall, these results illustrate how protective rare variant associations can directly nominate targets for therapeutic inhibition.

Our analysis also recapitulated and extended several gene-phenotype associations in which gene-level effects oppose the phenotypic correlations observed between traits (Table S18). For example, we identified the well-characterized *ANGPTL3* signal, where rare pLoF variants drive reductions in both HDL cholesterol and triglycerides (HDLC pLoF, MAF < 0.1%, burden *P* = 1.07 × 10^−32^; triglycerides: pLoF, MAF < 0.1%, burden *P* = 3.17 × 10^−126^), despite these traits being negatively correlated at the population level (Pearson *r* = −0.41, *P* < 1 × 10^−300^, Fig. S46a,b)^89^. Similarly, we observed the recently described *GIGYF1* association, where burden of rare pLoF variation is associated with increased adiposity (WHR adjusted for BMI: pLoF, MAF < 0.1%, burden *P* = 9.28 × 10^−9^) alongside reduced LDL cholesterol (pLoF, MAF < 0.1%, burden *P* = 3.17 × 10^−14^), in contrast to their typical positive phenotypic relationship (Pearson *r* = 0.088; *P* < 1 × 10^−300^; Fig. S46a,c)^90,91^. At *G6PC1*, rare putatively damaging variation was associated with both increased triglycerides (pLoF or DM/PA, MAF < 0.1%, burden *P* = 1.49 × 10^−32^) and reduced alanine transaminase (pLoF or DM/PA, MAF < 0.1%, burden *P* = 3.06 × 10^−7^), discordant with their positive phenotypic correlation (Pearson *r* = 0.21, *P* < 1 × 10^−300^, Fig. S46a,d). This pattern is consistent with the metabolic consequences of impaired hepatic glucose export, where accumulated glucose-6-phosphate is channeled into *de novo* lipogenesis, driving hypertriglyceridemia^92^ (Fig. S46d). Together, these findings highlight how rare coding variation can reveal gene-specific effects that are not apparent from population-level trait correlations.

Positive selection in spermatogonia has recently been shown to increase the effective mutation rate of specific genes through clonal expansion of mutant germline progenitors^93,94^. By increasing the transmission and population frequency of otherwise rare deleterious variants, clonal expansions in spermatogonia (CES) may increase the representation of such variants in population sequencing datasets and thereby influence the landscape of rare variant discovery. We therefore asked whether genes previously implicated in CES were enriched among BRaVa significant associations. We observed substantial overlap between BRaVa significant genes and genes previously implicated in positive selection in spermatogonia, with 13 such genes represented across 19 gene–phenotype pairs. BRaVa-significant genes were enriched for CES-related gene sets (adjusting for gene length, phyloP score^95^, and DepMap gene essentiality^96^). This included the candidate loss-of-function CES driver set of Seplyarskiy *et al*.^93^ (LoF-2), a subset of genes with elevated *de novo* mutation rates that nonetheless retain high functional polymorphism in the general population (OR = 24.2, *P* = 1.11 × 10^−8^; Table S19; Methods: Spermatogonia gene-set enrichment testing), as well as a larger overlapping gene set reported by Neville *et al*.^94^ (OR = 8.72, *P* = 8.56 × 10^−9^). These overlapping CES-related gene sets included canonical signaling genes such as *FGFR3* and *PTPN11*, but also multiple candidate loss-of-function CES drivers, including *MIB1*, *PPM1D*, *NF1*, *PTEN* and *SMAD6*. Several of these genes were associated with multiple traits, including *MIB1* across renal, lipid and metabolic phenotypes, and *PPM1D* across cancer, lipid and respiratory traits.

Collectively, these results demonstrate that large-scale rare variant meta-analysis across global biobanks reveals biologically coherent gene-trait associations spanning metabolic, cardiovascular, immune, and anthropometric domains, and provides a scalable framework for uncovering rare coding contributors to complex human disease.

## Discussion

BRaVa integrates rare variant association results across ten globally distributed biobanks, demonstrating that coordinated meta-analysis materially increases power to detect gene-level associations beyond those identifiable within any single cohort. These gains, observed across both disease endpoints and continuous traits, underscore the central role of scale and cross-population integration in rare variant discovery. Importantly, this framework is inherently scalable and can be extended as additional biobanks and sequencing resources emerge, enabling progressively well-powered analyses of less prevalent traits.

Beyond increasing discovery, these results provide insight into the biological architecture of complex disease. Many of the associations identified implicate genes involved in transcriptional and epigenetic regulation, metabolic homeostasis, vascular and epithelial biology, and immune function, highlighting the contribution of core regulatory pathways to disease risk. For example, associations at genes such as *SETD1A* and *ANKRD12* point to roles for chromatin and transcriptional control in inflammatory phenotypes, while signals at *NAA15*, *KEAP1*, and *GIGYF1* implicate protein modification, oxidative stress pathways, and translational regulation in metabolic traits. In several cases, we observe associations spanning multiple phenotypic domains, suggesting pleiotropic effects of rare coding variation. For instance, rare deleterious variation in *MIB1*, previously associated with T2D^19^, was also associated with chronic kidney failure and metabolic traits, potentially reflecting pleiotropy, mediation through T2D, or correlated disease ascertainment within cardiometabolic phenotypes. Given that *MIB1* regulates Notch signaling, a pathway implicated in kidney development and fibrosis, a direct role in kidney biology is also plausible. Additional signals at genes such as *PKD1*, *CGN/CGNL1*, and *PDZK1* further implicate vascular function, epithelial junction organization, and lipid transport pathways in disease susceptibility.

Interpretation of novelty in rare variant association studies also requires care. AI-assisted literature curation we employed was designed to summarize prior evidence for gene–trait relationships broadly and did not distinguish between burden-based association evidence, Mendelian disease reports, common variant associations, or other forms of biological support. Consequently, some associations classified as previously reported may nevertheless represent novel rare variant burden associations identified at biobank scale. For example, genes such as *NPPC* and *ACVR1C* have established biological links to height and adiposity-related phenotypes, respectively, yet population-scale burden evidence for these associations has remained limited.

The enrichment of CES-implicated genes among our significant findings suggests that positive selection in spermatogonia may influence the landscape of rare variant discovery. By elevating the *de novo* mutation rate at these loci, germline selection may increase the population frequency of otherwise highly constrained variants, and thus the power to detect associations relative to non-CES genes with similar effect sizes. In this setting, CES genes may achieve stronger statistical support not because they have larger phenotypic effects, but because they are represented by more carriers. At the same time, many CES genes function in core growth and developmental pathways, so their recurrence in our results may reflect a combination of increased mutational input and genuine biological pleiotropy. Together, these observations suggest that germline selection can influence which genes emerge from large-scale rare variant association studies.

Despite differences in recruitment strategy, phenotype ascertainment, and sequencing platforms across contributing studies, effect size estimates were generally consistent across biobanks and ancestry groups. These findings suggest that, while heterogeneity in study design is unavoidable in large consortia, careful harmonization and filtering can provide robust, interpretable rare genetic associations. However, pragmatic choices, such as minimum case count thresholds and allele frequency cutoffs, are necessary to ensure calibration and stability. These choices may exclude biologically meaningful signals near analytic boundaries, underscoring an inherent trade-off in large-scale rare variant studies.

Phenotype definitions were harmonized across biobanks using ICD and SNOMED mappings, but are not identical across healthcare systems due to differences in coding practices and ascertainment. As a result, residual heterogeneity in phenotype definition and prevalence is expected, and we observed variation in disease prevalence across biobanks consistent with differences in recruitment. Such heterogeneity may reduce power, particularly for rare variant analyses where the number of individuals carrying qualifying variants per gene is limited. In this setting, heterogeneity is more likely to attenuate true signals than to induce spurious associations, increasing the risk of false negatives. These effects are partially mitigated by mixed model association testing and stringent calibration filtering based on synonymous variation, although residual heterogeneity across cohorts is likely to remain. Heterogeneity in ascertainment and phenotype definition may also affect the magnitude of estimated effect sizes, which represent averages across cohorts with differing recruitment strategies, disease prevalence, and healthcare contexts. Accordingly, these estimates may not directly translate to risk prediction or penetrance in individual populations.

More broadly, these considerations reflect structural limitations of consortium-scale rare variant analyses, where heterogeneity in data generation, phenotype definition, and cohort overlap must be balanced against gains in statistical power.

This study also highlights important methodological considerations for large-scale rare variant meta-analysis. Overlap correction strategies developed for common variant meta-analysis, such as those implemented in METAL^50^, can lead to biased results in rare variant settings if used without caution. These observations emphasize the need for overlap-aware methods tailored specifically to rare variant aggregation and motivate continued methodological development as sequencing consortia scale further and overlap across studies becomes unavoidable.

Continued advances in variant deleteriousness prediction will improve power for gene-based association testing by more accurately prioritizing variants with functional impact^97–100^. As such predictors combine diverse sources of information, such as protein structure, multi-omics data and regional constraint^101–103^, improved annotation of damaging missense variation is likely to enhance signal-to-noise in aggregation tests and improve biological interpretation of significant rare variant associations.

Rare coding variation can provide compelling human genetic support for therapeutic target discovery, as illustrated by protective loss-of-function alleles in genes such as *PCSK9*, *ANGPTL3*, and *APOC3*^89,104,105^, which have helped motivate lipid-lowering therapeutic strategies. Rare variants that confer protection against disease therefore represent particularly valuable translational opportunities, as loss-of-function or protective alleles can provide direct human genetic support for therapeutic target inhibition. In contrast to activating strategies, pharmacologic inhibition is often more tractable in drug development, making protective genetic associations especially informative for target prioritization. However, therapeutic targeting of these pathways requires evaluation within a broader phenome-wide context, as loss-of-function variants may be pleiotropic and increase risk for other diseases. Characterizing these multi-phenotype effects requires very large whole-genome sequencing resources linked to health record data to capture sufficient carrier counts, underscoring the value of federated efforts such as BRaVa.

Gene-based meta-analysis methods that explicitly account for LD have the potential to further improve power and interpretability^106–109^. However, their application in large, multi-cohort consortia is currently constrained by data-sharing limitations, as LD for ultra-rare genetic variation can often not be shared due to privacy concerns.

Beyond these methodological considerations, several genomic regions remain unassessed in our current analyses. Future work will aim to systematically evaluate the Y chromosome, the mitochondrial genome, and structurally complex loci with established pleiotropic effects, such as the HLA and KIR regions.

Finally, BRaVa illustrates the complementary roles of rare and common variant analyses in elucidating disease biology. Many rare variant associations identified through BRaVa implicate genes with clear functional relevance, offering direct insights into disease mechanisms. Importantly, inclusion of non-European ancestry samples increased power beyond that achievable by European-only analyses and enabled evaluation of effect size consistency across populations, reinforcing the scientific value of diverse representation. As additional biobanks join BRaVa and sequencing efforts expand, this framework provides a scalable path toward increasingly comprehensive and equitable rare variant studies, with the potential to accelerate gene discovery and translational insight. By enabling analyses that are not feasible within any single biobank and releasing gene- and variant-level summary statistics to the community, BRaVa provides a shared resource and collaborative engine for future rare variant discovery across the global research community.

## Data and code availability

All meta-analysis results are available for download from google cloud storage at gs://brava-meta-analysis, using Requester Pays. These meta-analysis results consist of ancestry specific, cross-ancestry meta-analysis test statistics at the variant and gene level.

All associated scripts for quality control, meta-analysis, and plotting of results are available at https://github.com/BRaVa-genetics.

Individual-level genetic and phenotypic data are available from participating biobanks subject to their respective data access procedures and approvals.

Data from the National Genomic Research Library (NGRL) used in this research are available within the secure Genomics England Research Environment. Access to NGRL data is restricted to adhere to consent requirements and protect participant privacy. Data used in this research include: multi-sample population aggregate files in pgen format (aggV2), participant demographics such as year of birth and sex and additional electronic health record data (EHR) such as hospital episode statistics (HES) accessed via Labkey application for data release version 18. Access to NGRL data is provided to approved researchers who are members of the Genomics England Research Network, subject to institutional access agreements and research project approval under participant-led governance. For more information on data access, visit: https://www.genomicsengland.co.uk/research.

Access to UK Biobank data can be obtained through application to the UK Biobank Access Management System (https://www.ukbiobank.ac.uk/enable-your-research/apply-for-access).

Individual-level data from the All of Us Research Program are available to approved researchers through the All of Us Researcher Workbench under the program’s data access policies and procedures (https://www.researchallofus.org/). Access requires completion of the All of Us researcher registration, training processes, and institutional eligibility requirements.

## Acknowledgments

This research was conducted using the UK Biobank Resource (application 11867). D.S.P is supported by a Wellcome Trust Investigator Award (WTIA) (221782/Z/20/Z) and the Pioneer Centre for Statistical and Computational Methods for Advanced Research to Transform Biomedicine (SMARTbiomed). C.M.L. was supported by the Li Ka Shing Foundation, NIHR Oxford Biomedical Research Centre, NIH (1P50HD104224-01), Gates Foundation (INV-024200) and WTIA (221782/Z/20/Z). W.Z. was supported by NHGRI (K99/R00HG012222). We acknowledge the Pioneer Center for SMARTbiomed, DNRF grant number P4. The contributions of constituent biobanks were supported by a collection of grants. Biobank-specific acknowledgments and more detailed acknowledgments are provided in Methods: Biobank and cohort acknowledgments.

---

**Extended Data Figure 1:**
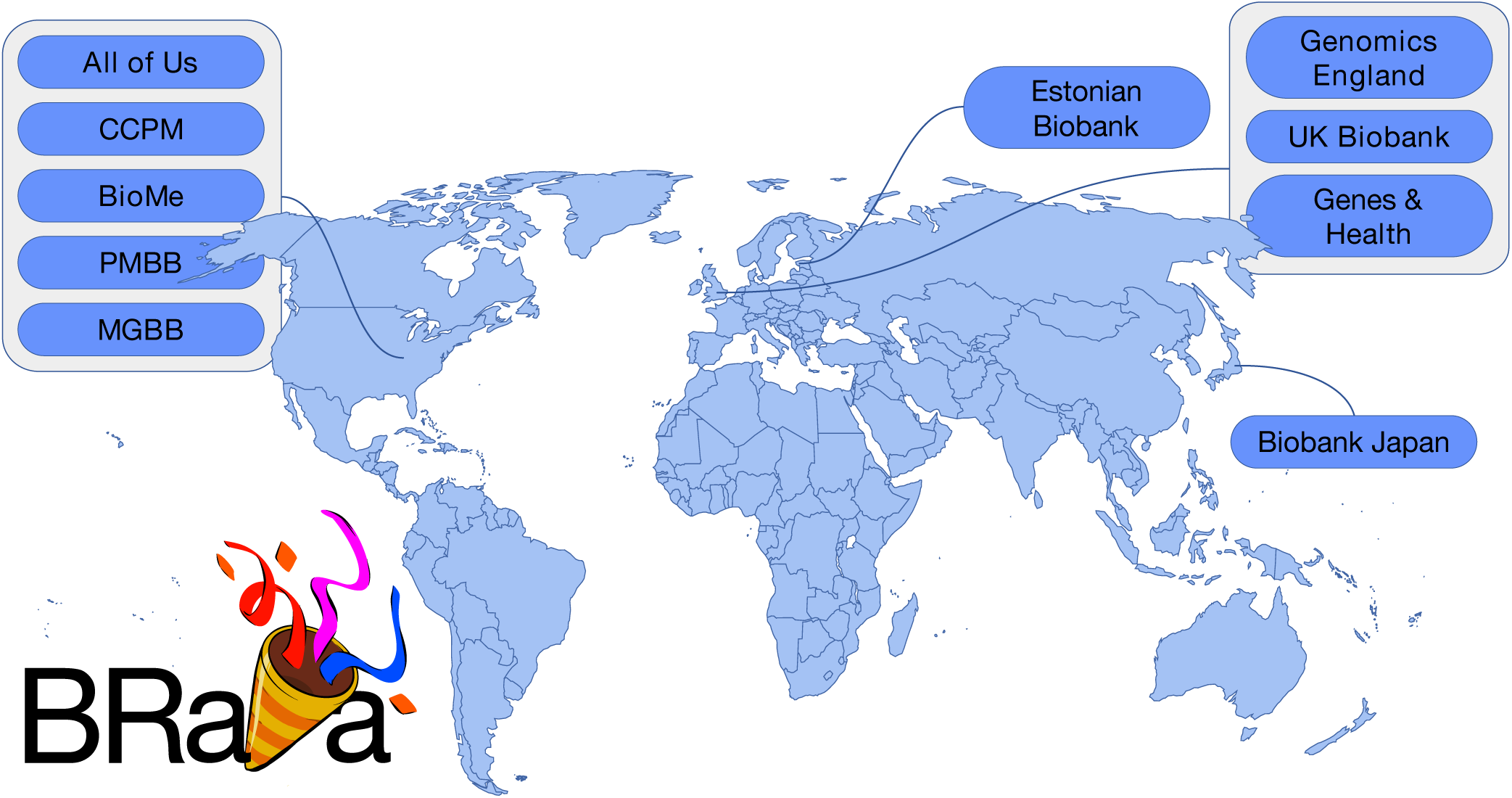
BRaVa consortium members. 10 biobanks and cohorts have contributed to this primary analysis of 44 diseases and traits. The total number of individuals represented across the constituent members of the collaboration now exceeds 1.2 million.

**Extended Data Figure 2:**
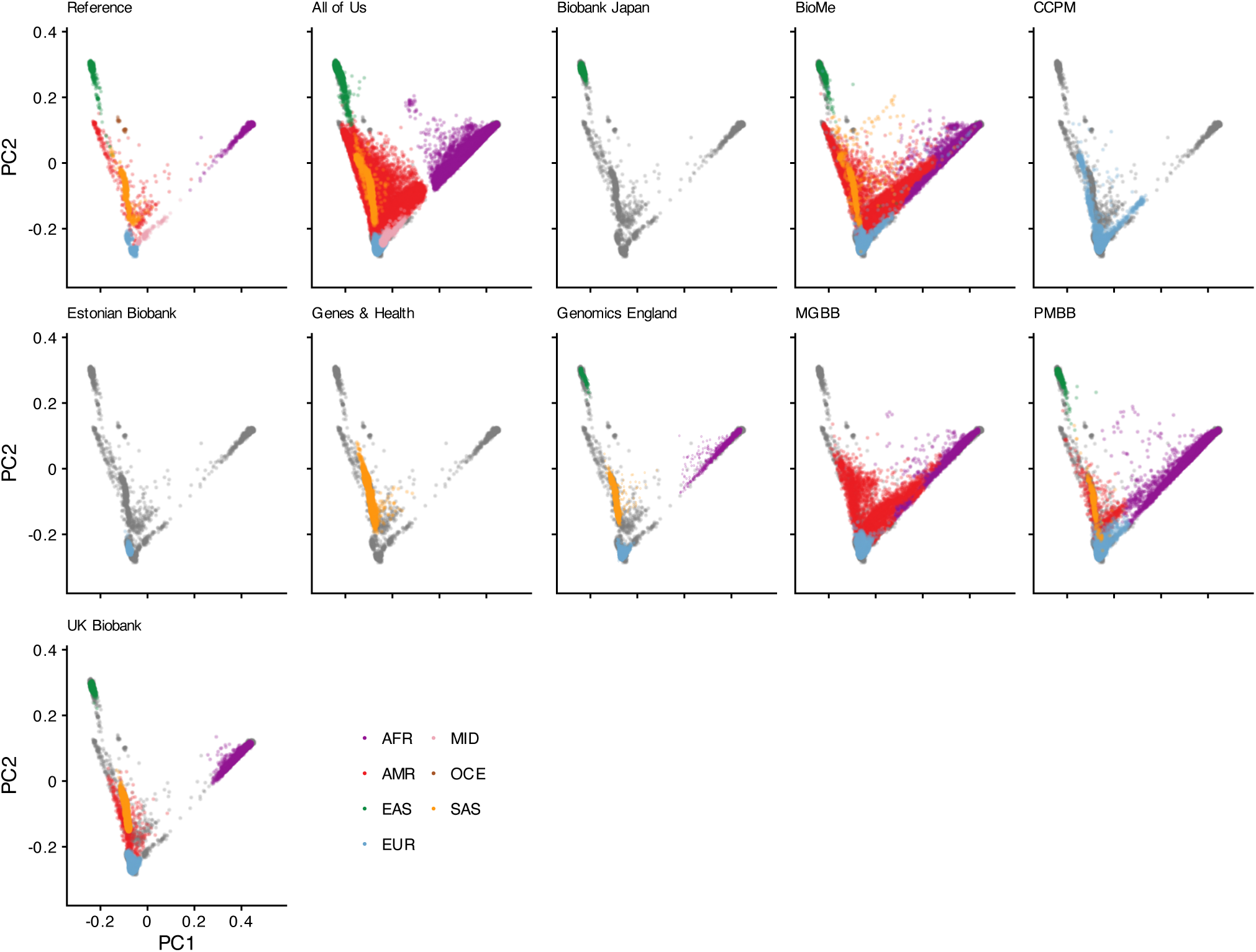
Projection of contributed samples from studies participating within the BRaVa consortium into the PC space defined by 1000G and HGDP. Using the PC space defined by 1000G and HGDP as a reference, analysts projected samples from contributing studies into the space using a common set of 165,000 variants. In each panel (except for the reference), colored points correspond to contributed samples from each cohort. Gray points on each panel (except the reference) denote the reference samples (which are colored in the first panel). Points are colored according to the genetic ancestry label determined by each contributing biobank. As GEL was only able to submit images of the projection of samples into PC space, we overlaid submitted transparent images onto reference samples, rather than plotting them. Here we show projections onto PCs 1 and 2. Figs. S1-S2 display the corresponding plots for PCs 3-6.

**Extended Data Figure 3:**
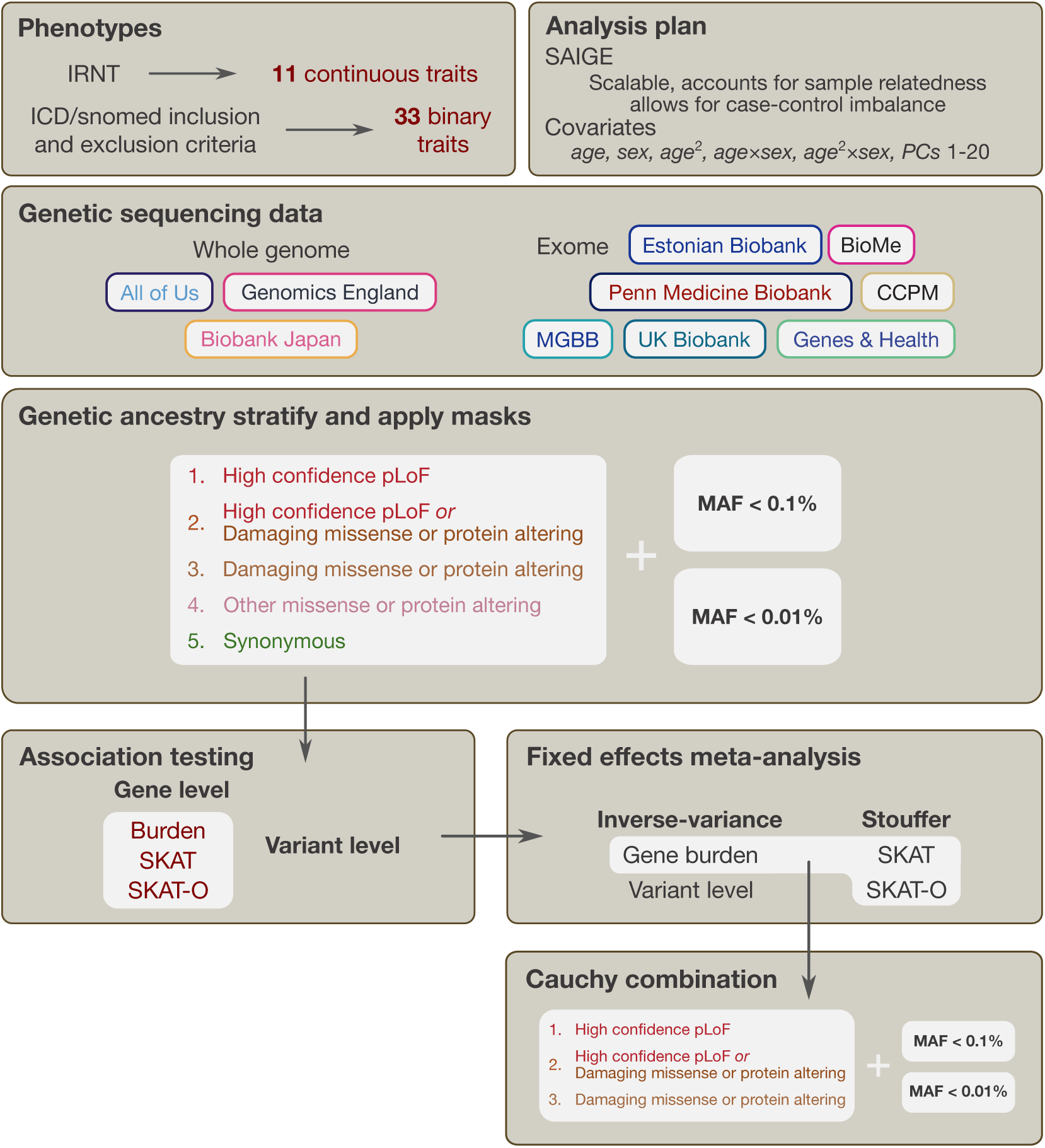
Workflow of the flagship pilot analysis project in BRaVa. Phenotypes were nominated by analysts from across the consortium. 17 analysts from across seven biobanks put forward the collection of the 44 pilot traits analyzed. Ten biobanks carried out analyses and contributed summary statistics data for meta-analysis. Among these, three cohorts analyzed WGS data, and seven analyzed ES data. We applied MAF masks at 0.1% and 0.01% and partitioned exonic variants into five classes (see Methods: Annotation definitions for gene-level association testing, for full details). We then carried out group-based testing at the gene-level, and variant association testing, using SAIGE. Finally, we performed fixed-effects meta-analysis on the resultant summary level data provided by constituent biobank analysts.

## References

1. Buniello, A. et al. The NHGRI-EBI GWAS Catalog of published genome-wide association studies, targeted arrays and summary statistics 2019. en. Nucleic Acids Res. 47, D1005–D1012 (Aug. 2019).

2. Visscher, P. M. et al. 10 years of GWAS discovery: Biology, function, and translation. en. Am. J. Hum. Genet. 101, 5–22 (June 2017).

3. Gaziano, J. M. et al. Million Veteran Program: A mega-biobank to study genetic influences on health and disease. en. J. Clin. Epidemiol. 70, 214–223 (Feb. 2016).

4. Karczewski, K. J. et al. Pan-UK Biobank genome-wide association analyses enhance discovery and resolution of ancestry-enriched effects. en. Nat. Genet. 57, 2408–2417 (Oct. 2025).

5. Verma, A. et al. Diversity and scale: Genetic architecture of 2068 traits in the VA Million Veteran Program. en. Science 385, eadj1182 (19 7 2024).

6. Milani, L. et al. The Estonian Biobank’s journey from biobanking to personalized medicine. en. Nat. Commun. 16, 3270 (May 2025).

7. Walters, R. G. et al. Genotyping and population characteristics of the China Kadoorie Biobank. en. Cell Genom. 3, 100361 (Sept. 2023).

8. Popejoy, A. B. & Fullerton, S. M. Genomics is failing on diversity. en. Nature 538, 161–164 (13 10 2016).

9. Martin, A. R. et al. Human demographic history impacts genetic risk prediction across diverse populations. en. Am. J. Hum. Genet. 100, 635–649 (June 2017).

10. Martin, A. R., Kanai, M., Kamatani, Y., Okada, Y., Neale, B. M. & Daly, M. J. Clinical use of current polygenic risk scores may exacerbate health disparities. en. Nat. Genet. 51, 584–591 (Apr. 2019).

11. Karczewski, K. J. et al. The mutational constraint spectrum quantified from variation in 141,456 humans. en. Nature 581, 434–443 (May 2020).

12. Cirulli, E. T. & Goldstein, D. B. Uncovering the roles of rare variants in common disease through whole-genome sequencing. en. Nat. Rev. Genet. 11, 415–425 (June 2010).

13. Bycroft, C. et al. The UK Biobank resource with deep phenotyping and genomic data. en. Nature 562, 203–209 (Oct. 2018).

14. Nagai, A. et al. Overview of the BioBank Japan Project: Study design and profile. en. J. Epidemiol. 27, S2–S8 (Mar. 2017).

15. Karczewski, K. J. et al. Systematic single-variant and gene-based association testing of thousands of phenotypes in 394,841 UK Biobank exomes. en. Cell Genom. 2, 100168 (14 9 2022).

16. Backman, J. D. et al. Exome sequencing and analysis of 454,787 UK Biobank participants. en. Nature 599, 628–634 (Nov. 2021).

17. All of Us Research Program Genomics Investigators. Genomic data in the All of Us Research Program. en. Nature 627, 340–346 (Mar. 2024).

18. All of Us Research Program Investigators et al. The “All of Us” Research Program. en. N. Engl. J. Med. 381, 668–676 (15 8 2019).

19. Jurgens, S. J. et al. Rare coding variant analysis for human diseases across biobanks and ancestries. en. Nat. Genet. 56, 1811–1820 (Sept. 2024).

20. Zhou, W. et al. Global Biobank Meta-analysis Initiative: Powering genetic discovery across human disease. en. Cell Genom. 2, 100192 (Dec. 2022).

21. 100,000 Genomes Project Pilot Investigators et al. 100,000 genomes pilot on rare-disease diagnosis in health care - preliminary report. en. N. Engl. J. Med. 385, 1868–1880 (Nov. 2021).

22. Finer, S. et al. Cohort Profile: East London Genes & Health (ELGH), a community-based population genomics and health study in British Bangladeshi and British Pakistani people. en. Int. J. Epidemiol. 49, 20–21i (Jan. 2020).

23. Verma, A. et al. The Penn Medicine BioBank: Towards a genomics-enabled learning healthcare system to accelerate precision medicine in a diverse population. en. J. Pers. Med. 12, 1974 (29 11 2022).

24. Wiley, L. K. et al. Building a vertically integrated genomic learning health system: The biobank at the Colorado Center for Personalized Medicine. en. Am. J. Hum. Genet. 111, 11–23 (Apr. 2024).

25. Boutin, N. T. et al. The evolution of a large Biobank at mass general Brigham. en. J. Pers. Med. 12, 1323 (17 8 2022).

26. Karlson, E. W., Boutin, N. T., Hoffnagle, A. G. & Allen, N. L. Building the Partners HealthCare Biobank at partners personalized medicine: Informed consent, return of research results, recruitment lessons and operational considerations. en. J. Pers. Med. 6, 2 (14 1 2016).

27. Belbin, G. M. et al. Toward a fine-scale population health monitoring system. en. Cell 184, 2068–2083.e11 (15 4 2021).

28. 1000 Genomes Project Consortium et al. A global reference for human genetic variation. en. Nature 526, 68–74 (Jan. 2015).

29. Cavalli-Sforza, L. L. The Human Genome Diversity Project: past, present and future. en. Nat. Rev. Genet. 6, 333–340 (Apr. 2005).

30. Donnelly, K. The advanced terminology and coding system for eHealth. Stud Health Technol Inform. 121, 279–290 (2006).

31. Denny, J. C. et al. PheWAS: demonstrating the feasibility of a phenome-wide scan to discover gene-disease associations. en. Bioinformatics 26, 1205–1210 (Jan. 2010).

32. Wu, P. et al. Mapping ICD-10 and ICD-10-CM codes to phecodes: Workflow development and initial evaluation. en. JMIR Med. Inform. 7, e14325 (29 11 2019).

33. World Health Organization. International Statistical Classification of Diseases and Related Health Problems, 10th Revision (ICD-10) (WHO, Geneva).

34. universal-saige github repository https://github.com/BRaVa-genetics/universal-saige.

35. Zhou, W. et al. Efficiently controlling for case-control imbalance and sample relatedness in large-scale genetic association studies. en. Nat. Genet. 50, 1335–1341 (Sept. 2018).

36. Zhou, W. et al. Scalable generalized linear mixed model for region-based association tests in large biobanks and cohorts. en. Nat. Genet. 52, 634–639 (June 2020).

37. Zhou, W. et al. SAIGE-GENE+ improves the efficiency and accuracy of set-based rare variant association tests. en. Nat. Genet. 54, 1466–1469 (Oct. 2022).

38. Dey, R., Schmidt, E. M., Abecasis, G. R. & Lee, S. A fast and accurate algorithm to test for binary phenotypes and its application to PheWAS. en. Am. J. Hum. Genet. 101, 37–49 (June 2017).

39. Heinze, G. & Schemper, M. A solution to the problem of separation in logistic regression. en. Stat. Med. 21, 2409–2419 (30 8 2002).

40. McLaren, W. et al. The Ensembl Variant Effect Predictor. en. Genome Biol. 17, 122 (June 2016).

41. Ioannidis, N. M. et al. REVEL: An ensemble method for predicting the pathogenicity of rare missense variants. en. Am. J. Hum. Genet. 99, 877–885 (Oct. 2016).

42. Rentzsch, P., Witten, D., Cooper, G. M., Shendure, J. & Kircher, M. CADD: predicting the deleteriousness of variants throughout the human genome. en. Nucleic Acids Res. 47, D886–D894 (Aug. 2019).

43. Jaganathan, K. et al. Predicting Splicing from Primary Sequence with Deep Learning. en. Cell 176, 535–548.e24 (Jan. 2019).

44. Brown, B. C., Ye, C. J., Price, A. L. & Zaitlen, N. Transethnic genetic-correlation estimates from summary statistics. en. Am. J. Hum. Genet. 99, 76–88 (July 2016).

45. Galinsky, K. J. et al. Estimating cross-population genetic correlations of causal effect sizes. en. Genet. Epidemiol. 43, 180–188 (Mar. 2019).

46. Liu, J. Z. et al. Association analyses identify 38 susceptibility loci for inflammatory bowel disease and highlight shared genetic risk across populations. en. Nat. Genet. 47, 979–986 (Sept. 2015).

48. Stouffer, S. A., Suchman, E. A., Devinney, L. C., Star, S. A. & Williams, R. M. The American Soldier, Volume I: Adjustment During Army Life (1949).

48. Wu, M. C., Lee, S., Cai, T., Li, Y., Boehnke, M. & Lin, X. Rare-variant association testing for sequencing data with the sequence kernel association test. en. Am. J. Hum. Genet. 89, 82–93 (15 7 2011).

49. Lee, S. et al. Optimal unified approach for rare-variant association testing with application to small-sample case-control whole-exome sequencing studies. en. Am. J. Hum. Genet. 91, 224–237 (Oct. 2012).

50. Willer, C. J., Li, Y. & Abecasis, G. R. METAL: fast and efficient meta-analysis of genomewide association scans. en. Bioinformatics 26, 2190–2191 (Jan. 2010).

51. https://genome.sph.umich.edu/w/images/7/7b/METAL_sample_overlap_method_2017-11-15.pdf. Accessed: 2026-1-21.

52. Lin, D.-Y. & Sullivan, P. F. Meta-analysis of genome-wide association studies with overlapping subjects. en. Am. J. Hum. Genet. 85, 862–872 (Dec. 2009).

53. Genovese, G. score: Tools to work with GWAS-VCF summary statistics files en.

54. Nguyen, T. et al. A resource of “bottom-line” variant associations for 1,281 complex traits by integrating data across published genome-wide association studies. en. Res. Sq. (22 1 2026).

55. Buniello, A. et al. Open Targets Platform: facilitating therapeutic hypotheses building in drug discovery. en. Nucleic Acids Res. 53, D1467–D1475 (June 2025).

56. Akbari, P. et al. Sequencing of 640,000 exomes identifies GPR75 variants associated with protection from obesity. en. Science 373, eabf8683 (Feb. 2021).

57. Wang, Q. et al. Rare variant contribution to human disease in 281,104 UK Biobank exomes. en. Nature 597, 527–532 (Sept. 2021).

58. Välimäki, N. et al. Inherited mutations affecting the SRCAP complex are central in moderate-penetrance predisposition to uterine leiomyomas. en. Am. J. Hum. Genet. 110, 460–474 (Feb. 2023).

59. Choi, S. H. et al. Sequencing in over 50,000 cases identifies coding and structural variation underlying atrial fibrillation risk. en. Nat. Genet. 57, 548–562 (Mar. 2025).

60. Ding, C. et al. Research progress on the role and inhibitors of Keap1 signaling pathway in inflammation. en. Int. Immunopharmacol. 141, 112853 (15 11 2024).

61. Akbari, P. et al. Multiancestry exome sequencing reveals INHBE mutations associated with favorable fat distribution and protection from diabetes. en. Nat. Commun. 13, 4844 (23 8 2022).

62. Deaton, A. M. et al. Rare loss of function variants in the hepatokine gene INHBE protect from abdominal obesity. en. Nat. Commun. 13, 4319 (27 7 2022).

63. Baya, N. A. et al. Combining evidence from human genetic and functional screens to identify pathways altering obesity and fat distribution. en. Am. J. Hum. Genet. (29 8 2025).

64. Setoguchi, K. et al. The SNP c.1326T>G in the non-SMC condensin I complex, subunit G (NCAPG) gene encoding a p.Ile442Met variant is associated with an increase in body frame size at puberty in cattle. en. Anim. Genet. 42, 650–655 (Dec. 2011).

65. Bouwman, A. C. et al. Meta-analysis of genome-wide association studies for cattle stature identifies common genes that regulate body size in mammals. en. Nat. Genet. 50, 362–367 (Mar. 2018).

66. Signer-Hasler, H. et al. A genome-wide association study reveals loci influencing height and other conformation traits in horses. en. PLoS One 7, e37282 (16 5 2012).

67. Metzger, J., Schrimpf, R., Philipp, U. & Distl, O. Expression levels of LCORL are associated with body size in horses. en. PLoS One 8, e56497 (13 2 2013).

68. Zhao, Y. et al. Protein-truncating variants in BSN are associated with severe adult-onset obesity, type 2 diabetes and fatty liver disease. en. Nat. Genet. 56, 579–584 (Apr. 2024).

69. Cheng, W. et al. Calcitonin receptor neurons in the mouse nucleus tractus solitarius control energy balance via the non-aversive suppression of feeding. en. Cell Metab. 31, 301–312.e5 (Apr. 2020).

70. Tadross, J. A. et al. A comprehensive spatio-cellular map of the human hypothalamus. en. Nature 639, 708–716 (Mar. 2025).

71. Kim, K., Drummond, I., Ibraghimov-Beskrovnaya, O., Klinger, K. & Arnaout, M. A. Polycystin 1 is required for the structural integrity of blood vessels. en. Proc. Natl. Acad. Sci. U. S. A. 97, 1731–1736 (15 2 2000).

72. Perrone, R. D., Malek, A. M. & Watnick, T. Vascular complications in autosomal dominant polycystic kidney disease. en. Nat. Rev. Nephrol. 11, 589–598 (Oct. 2015).

73. Aksnes, H., Ree, R. & Arnesen, T. Co-translational, post-translational, and non-catalytic roles of N-terminal acetyltransferases. en. Mol. Cell 73, 1097–1114 (21 3 2019).

74. Gharahkhani, P. et al. Genome-wide meta-analysis identifies 127 open-angle glaucoma loci with consistent effect across ancestries. en. Nat. Commun. 12, 1258 (24 2 2021).

75. Sakaue, S. et al. A cross-population atlas of genetic associations for 220 human phenotypes. en. Nat. Genet. 53, 1415–1424 (Oct. 2021).

76. Ghoussaini, M. et al. Open Targets Genetics: systematic identification of trait-associated genes using large-scale genetics and functional genomics. en. Nucleic Acids Res. 49, D1311–D1320 (Aug. 2021).

77. Kocher, O. et al. Influence of PDZK1 on lipoprotein metabolism and atherosclerosis. en. Biochim. Biophys. Acta 1782, 310–316 (May 2008).

78. Trigatti, B. L. SR-B1 and PDZK1: partners in HDL regulation. en. Curr. Opin. Lipidol. 28, 201–208 (Apr. 2017).

79. Kocher, O. & Krieger, M. Role of the adaptor protein PDZK1 in controlling the HDL receptor SR-BI. en. Curr. Opin. Lipidol. 20, 236–241 (June 2009).

80. Tumes, D. J., Papadopoulos, M., Endo, Y., Onodera, A., Hirahara, K. & Nakayama, T. Epigenetic regulation of T-helper cell differentiation, memory, and plasticity in allergic asthma. en. Immunol. Rev. 278, 8–19 (July 2017).

81. Alashkar Alhamwe, B., Miethe, S., Pogge von Strandmann, E., Potaczek, D. P. & Garn, H. Epigenetic regulation of airway epithelium immune functions in asthma. en. Front. Immunol. 11, 1747 (18 8 2020).

82. Enomoto, A. et al. Molecular identification of a renal urate anion exchanger that regulates blood urate levels. en. Nature 417, 447–452 (23 5 2002).

83. Sakiyama, M. et al. The effects of URAT1/SLC22A12 nonfunctional variants, R90H and W258X, on serum uric acid levels and gout/hyperuricemia progression. en. Sci. Rep. 6, 20148 (29 1 2016).

84. Guo, W. et al. Mechanisms of urate transport and uricosuric drugs inhibition in human URAT1. en. Nat. Commun. 16, 1512 (Oct. 2025).

85. Presume, J., Ferreira, J. & Ribeiras, R. Factor XI inhibitors: A new horizon in anticoagulation therapy. en. Cardiol. Ther. 13, 1–16 (Mar. 2024).

86. Preis, M. et al. Factor XI deficiency is associated with lower risk for cardiovascular and venous thromboembolism events. en. Blood 129, 1210–1215 (Feb. 2017).

87. Balla, T. Phosphoinositides: tiny lipids with giant impact on cell regulation. en. Physiol. Rev. 93, 1019–1137 (July 2013).

88. Bojjireddy, N. et al. Pharmacological and genetic targeting of the PI4KA enzyme reveals its important role in maintaining plasma membrane phosphatidylinositol 4-phosphate and phosphatidylinositol 4,5-bisphosphate levels. en. J. Biol. Chem. 289, 6120–6132 (28 2 2014).

89. Dewey, F. E. et al. Genetic and pharmacologic inactivation of ANGPTL3 and cardiovascular disease. en. N. Engl. J. Med. 377, 211–221 (20 7 2017).

90. Deaton, A. M. et al. Gene-level analysis of rare variants in 379,066 whole exome sequences identifies an association of GIGYF1 loss of function with type 2 diabetes. en. Sci. Rep. 11 (Mar. 2021).

91. Zhao, Y. et al. GIGYF1 loss of function is associated with clonal mosaicism and adverse metabolic health. en. Nat. Commun. 12, 4178 (July 2021).

92. Chou, J. Y., Jun, H. S. & Mansfield, B. C. Glycogen storage disease type I and G6Pase- deficiency: etiology and therapy. en. Nat. Rev. Endocrinol. 6, 676–688 (Dec. 2010).

93. Seplyarskiy, V. et al. Hotspots of human mutation point to clonal expansions in spermatogonia. en. Nature 647, 429–435 (Nov. 2025).

94. Neville, M. D. C. et al. Sperm sequencing reveals extensive positive selection in the male germline. en. Nature 647, 421–428 (Nov. 2025).

95. Pollard, K. S., Hubisz, M. J., Rosenbloom, K. R. & Siepel, A. Detection of nonneutral substitution rates on mammalian phylogenies. en. Genome Res. 20, 110–121 (Jan. 2010).

96. Arafeh, R., Shibue, T., Dempster, J. M., Hahn, W. C. & Vazquez, F. The present and future of the Cancer Dependency Map. en. Nat. Rev. Cancer 25, 59–73 (Jan. 2025).

97. Cheng, J. et al. Accurate proteome-wide missense variant effect prediction with AlphaMissense. en. Science 381, eadg7492 (22 9 2023).

98. Fiziev, P. P. et al. Rare penetrant mutations confer severe risk of common diseases. en. Science 380, eabo1131 (Feb. 2023).

99. Frazer, J. et al. Disease variant prediction with deep generative models of evolutionary data. en. Nature 599, 91–95 (Nov. 2021).

100. Clarke, B. et al. Integration of variant annotations using deep set networks boosts rare variant association testing. en. Nat. Genet. 56, 2271–2280 (Oct. 2024).

101. Samocha, K. E., Kosmicki, J. A., Karczewski, K. J., et al. Regional missense constraint improves variant deleteriousness prediction. BioRxiv (2017).

102. Fang, Z. et al. AlphaFold 3: an unprecedent opportunity for fundamental research and drug development. en. Precis. Clin. Med. 8, baf015 (Sept. 2025).

103. Gerges, S. et al. Genetics-to-structure multiscale analysis identifies disrupted calcium homeostasis as a mechanism of psychiatric disease. Genetics (27 8 2025).

104. Cohen, J. C., Boerwinkle, E., Mosley Jr, T. H. & Hobbs, H. H. Sequence variations in PCSK9, low LDL, and protection against coronary heart disease. en. N. Engl. J. Med. 354, 1264–1272 (23 3 2006).

105. TG and HDL Working Group of the Exome Sequencing Project, National Heart, Lung, and Blood Institute et al. Loss-of-function mutations in APOC3, triglycerides, and coronary disease. en. N. Engl. J. Med. 371, 22–31 (Mar. 2014).

106. Joseph, T. A. et al. Computationally efficient meta-analysis of gene-based tests using summary statistics in large-scale genetic studies. en. Nat. Genet. 57, 3193–3200 (Dec. 2025).

107. Park, E. et al. Scalable and accurate rare variant meta-analysis with Meta-SAIGE. en. Nat. Genet. 57, 3185–3192 (Dec. 2025).

108. Lee, S., Teslovich, T. M., Boehnke, M. & Lin, X. General framework for meta-analysis of rare variants in sequencing association studies. en. Am. J. Hum. Genet. 93, 42–53 (Nov. 2013).

109. Li, X. et al. Powerful, scalable and resource-efficient meta-analysis of rare variant associations in large whole genome sequencing studies. en. Nat. Genet. 55, 154–164 (Jan. 2023).

