## Supplementary Materials for "The Biobank Rare Variant consortium powers the discovery of rare genetic associations through global collaboration"

Step 1: Fit the null generalized linear model (linear or logistic for continuous and binary traits, respectively) 51

Step 2: Perform association testing for each variant or set of variants (burden and variance based tests) . 52

**Summary statistic quality control** **53**

**Cross-ancestry comparison of gene-level burden effect sizes** **53**

Data preparation . . . . . . . . . . . . . . . . . . . . . . . . . . . . . . . . . . . . . . . 54

Deming regression analysis . . . . . . . . . . . . . . . . . . . . . . . . . . . . . . . . . . 54

**Meta-analysis** **55**

**Cumulative allele frequency weighted Stouffer's meta-analysis for SKAT and SKAT-O** **57**

**Caveats of METAL sample overlap correction for rare variant analyses** **58**

**Accounting for overlap in fixed effects meta-analysis** **60**

Sample size based meta-analysis . . . . . . . . . . . . . . . . . . . . . . . . . . . . . . . 60

Determining optimal weights when there is sample overlap . . . . . . . . . . . . . . . . . . . . 61

**Locus definition for variant-level results** **64**

**Common variant conditioning** **64**

**Novelty assessment of gene–phenotype associations** **65**

**Biobank and cohort acknowledgments** **65**

**Supplementary Figures** **72**

**Supplementary Tables** **113**

**References** **114**

### Biobank descriptions

#### All of Us

The All of Us Research Program, launched by the NIH, is building a cohort of at least one million participants from across the United States, with a focus on including populations that have historically been under-represented in biomedical research[1]. All of Us collects biospecimens, electronic health records (EHRs), physical measurements, and survey data. To date, ~400,000 individuals have been whole-genome sequenced using Illumina NovaSeq 6000 machines and following the manufacturer's best practices.

#### Biobank Japan

Biobank Japan, established in 2003, has collected genetic and clinical data from approximately 270,000 participants diagnosed with 51 targeted diseases. Among these individuals, a subset of ~10,000 have been whole-genome sequenced.

#### Bio*Me*

Run by the Icahn School of Medicine at Mount Sinai, Bio*Me* consists of genetic sequencing data from over 31,000 participants in the Mount Sinai Health System (MSHS) in New York, NY, linked to EHRs. The cohort is one of the most ethnically diverse in the United States. Participant recruitment into Bio*Me* has been ongoing since 2007 and occurs predominantly through ambulatory care practices across the MSHS.

#### Colorado Center for Personalized Medicine

Part of the University of Colorado Anschutz Medical Campus, the Colorado Center for Personalized Medicine (CCPM) biobank comprises data from over 250,000 participants linked to EHR information. Participant recruitment occurs through an electronic consent opt-in model through the UC Health My Health Connection Portal. Among these, a subset of samples have been exome-sequenced in partnership with the Regeneron Genetics Center using the Twist platform.

#### Estonian Biobank

The Estonian Biobank is a population-based biobank managed by the University of Tartu, comprising genetic and health data from over 200,000 participants - around 20% of Estonia's adult population. It supports research on gene-environment interactions, disease risk prediction, and personalized medicine.

#### Genes & Health

Genes & Health is a long-term, community-based study of British Pakistani and British Bangladeshi individuals aged 16 years and older living in the UK. Since recruitment started in 2015, more than 60,000 individuals have contributed their genetic, health, and lifestyle data. Since recruitment started in 2015, more than 60,000 individuals have contributed their genetic, health, and lifestyle data, though here we focus on a pilot subset of 39,148 individuals who had both genotype and ES data.

#### Genomics England

Established in 2013, Genomics England completed the '100,000 genomes project' (100kGP) from NHS patients with rare diseases, cancers, and infectious diseases. 100kGP is a large-scale clinical sequencing initiative within the UK's National Health Service (NHS). It focuses on collecting WGS and clinical data from NHS patients with rare diseases, cancers, and infections. In the case of rare disease, 100kGP also recruits relatives in a family-based study design.

#### Mass General Brigham Biobank

Mass General Brigham Biobank (MGBB) consists of genetic and clinical data from more than 135,000 participants. It links biospecimens with EHR data from a multicenter health system in Massachusetts, USA.

#### Penn Medicine Biobank

The Penn Medicine Biobank (PMBB) has enrolled over 60,000 participants to date, collecting biospecimens and linking them to EHR data, including diagnoses, labs, procedures, medications, and structured data in Observational Medical Outcomes Partnership (OMOP). Among these, a subset of ~45,000 have been exome-sequenced in partnership with the Regeneron Genetics Center using the Illumina NovaSeq xGen exome capture from Integrated DNA technologies.

#### UK Biobank

The UK Biobank (UKB) is a large population-based prospective cohort study from the UK comprising genetic, health, and lifestyle data from more than 500,000 individuals aged 40–69 at recruitment (2006–2010).

### Phenotype definitions

Phenotypes were assigned based on ICD9, ICD10, and SNOMED based case inclusion and control exclusion criteria detailed in Table S1. These definitions were a combination of phecodes, ICD based encodings of phenotypes provided by the global health data exchance (GHDx), and manual definitions provided by disease experts within the consortium. We created scripts to take in comma delimited vectors of hierarchical disease encoding per individual, to automate case/control status for each trait[2]. Study participants were labeled a phecode if they had one or more of the phecode-specific ICD-9, ICD-10, or SNOMED codes. To assign cases and controls, we first set all individuals to controls, individuals with control exclusion criteria are set to missing, and finally individuals with ICD/SNOMED case inclusion codes are set to cases.

We are assume the hierarchical structure of the ICD based encodings, so if an analyst includes a two digit ICD code in the case inclusion criteria, then we assume that all ICD codes below that node in the hierarchy are included in the case criteria. This also allows simplification of the regular expression e.g. for acute appendicitis, the control exclusion criteria:

```
K36, K38, K38.0, K38.1, K38.2, K38.3, K38.8, K38.9, K37, K36, K38.3, K38.8, K38.2, K38.0,
K38.9, K38, K38.1
```

is mapped to the regular expression:

```
(^|\s)K36|(^|\s)K37|(^|\s)K38
```

For a collection of sex-specific traits and breast cancer (Table S1), only females were included in the study samples. The procedure endpoint, hip replacement was defined based on the OPSC.

As ICD/SNOMED/OPSC based definitions were not available for all constituent biobanks, we detail any deviation from the proposed BRaVa phenotype definitions:

#### Biobank Japan

We defined phenotypes based on the doctors' diagnoses at the cooperative hospitals of Biobank Japan and past medical histories extracted from the medical records. Both of the data sources were stored in the form of the disease names.

#### Bio*Me*

Phenotype definitions were pulled directly from the BRaVa definitions for ICD based definitions. Quantitative phenotypes also followed BRaVa's definitions, but were not pulled directly as their ascertainment requires more care. To ensure quality control, the BioMe team ran their in house ICD extraction pipeline multiple times and studied extracted participants. Doing so isolated errors in ICD definitions, which were communicated back to the consortium.

##### Genes & Health

Genes & Health binary trait phenotype data was ascertained from routine electronic health record participant data from primary care (in the form of SNOMED codes) and secondary care (in the form of ICD10 codes). SNOMED codes were mapped to 3- and 4-digit ICD10 codes within Genes & Health as previously described[3]. Quantitative trait data was also ascertained from routine electronic health record data from both primary and secondary care data sources. Data was processed as previously described[3].

Following case and control definitions, we wished to define a collection of traits that are present across a large proportion of biobanks and genetic ancestries across the BRaVa consortium, to best showcase the increased power garnered through federated meta-analysis. To do this, we restricted nominated traits to the subset that satisfy:

**Binary traits:** $> 100$ cases in at least five biobanks/cohorts and at least ten (biobank, genetic ancestry) pairs OR $> 1\%$ case prevalence in UK Biobank across all genetic ancestries.

**Continuous traits:** Total count $> 200,000$ across all (biobank, genetic ancestry) sub-cohorts.

#### Quality control guidance for sequencing data

To ensure consistency across biobanks, general guidelines for variant- and sample-level quality control, and ancestry were collaboratively defined in advance. Each biobank then performed quality control independently, following these shared principles, which we describe in this section. To support practical implementation, analysts were allowed to apply biobank-specific parameters or thresholds if deemed appropriate.

##### Sequencing Coverage

Sequencing coverage is left to the user's discretion, taking into account sample size and cost considerations, with priority given to maximizing sample size.

##### Batch Effects

Batch effects may occur when sequencing is performed on different platforms for cases and controls, or if controls are obtained from other studies. Sequencing all samples using the same sequencing platform and joint-calling together will reduce batch effects. If subsets of samples have been sequenced using disparate platforms, we recommend flagging samples appropriately. We recommend that samples be joint-called whenever possible. In all cases, we suggest plotting all QC metrics separately by batch to ensure even quality across platforms. Preferably, all case-control and quantitative analysis should be carried out on joint-called samples. Using internal or external datasets (e.g. gnomAD) as controls without joint-calling will introduce bias and as such we did not accept such data as part of this analysis.

#### Platform Usage

We recommend using a GDPR-compliant cloud computing platform for the full analysis workflow to facilitate ease of use, sharing, and efficiency. The Google Cloud Platform is recommended.

#### Variant Calling

##### Read Processing

We recommend processing sequencing reads (`.fastq/.bam/.cram` files) using GATK. For `.fastq` files, paired reads should be represented in two files one with first in pair reads, one with second in pair reads (interleaved `.fastq` files must be converted).

If samples are multiplexed (recommended), the following metadata descriptors should be provided per sample:

- Readgroup (unique identifier)
- Sample name
- Library name (e.g. `Solexa-*`)
- Platform unit (e.g. `<flowcell>.<lane>.<barcode>`)
- Run date
- Platform name (e.g. Illumina)
- Sequencing center (e.g. BI, BCM, NYGC)

Suggestions in this section were adapted from the Broad Centers for Common Disease Genomics project[4].

##### Quality Metrics

We recommend computing quality metrics (e.g. % chimeras) using CollectHsMetrics. Target intervals should be extracted from the calling intervals available from:

```
gs://gcp-public-data-broad-references
```

Suggested flags:

**Software: CollectHsMetrics**

**Standardized flags**

`-I $input_bam`

`-R $ref_fasta`

`-TARGET_INTERVALS $wes_coverage_interval_list`

`-BAIT_INTERVALS $wes_coverage_interval_list`

`-O $metrics_filename`

For whole-genome sequencing, padded exon windows may be used when analyzing the exome.

#### Alignment

We follow the functional equivalence standard described by Regier *et al.*[5].

We recommend BWA-MEM v0.7[6], aligning to the GRCh38DH reference genome[7]. Samples processed with alternative reference builds may be lifted over, with artifacts removed post-liftover.

**Software: bwa**

**Standardized flags**

`-K 100000000` to achieve deterministic alignment results (note: this is a hidden option)

`-Y` to force soft-clipping rather than default hard-clipping of supplementary alignments

Include a .alt file for consumption by BWA-MEM; do not perform post-processing of alternate alignments

Do **not** use `-M` since it causes split-read alignments to be marked as 'secondary' rather than 'supplementary' alignments, violating the .bam specification

**Optional flags**

`-p` (for interleaved fastq)

`-C` (append `.fasta/.fastq` comment to SAM output)

`-v` (logging verbosity)

`-t` (threading)

`-R` (read group header line)

Note that the `-M` flag should not be used.

Upon successful alignment, we suggest an additional post-alignment step. In order to reduce false positive calls due to bacterial contamination randomly aligning to the human genome, reads and their mates may be marked by setting 0x4 bit in the .sam flag if the following conditions apply:

1. The primary alignment has less than 32 aligned bases.
2. The primary alignment is soft clipped on both sides.

The original mapping information will be encoded in a Previous Alignment (PA) tag on the marked reads using the same format as the SA tag in the `.bam` specification[8].

##### Marking Duplicates

We recommend using Picard[9] for duplicate marking. Orphan alignments are marked as duplicates if another read shares the same alignment, and the unmapped mate of duplicate orphan reads should also be marked as duplicate.

##### Base Quality Score Recalibration

As part of the GATK suite of tools, we suggest that this step uses GATK BaseRecalibrator where recalibration files can be copied from the GATK hg38 bundle bucket[10], specifically:

- `Homo_sapiens_assembly38.dbsnp138.vcf`
- `Mills_and_1000G_gold_standard.indels.hg38.vcf.gz`
- `Homo_sapiens_assembly38.known_indels.vcf.gz`

Flags that can be used include the following:

**Software: BaseRecalibrator**

**Standardized flags**

```
-R $ref_fasta
-I $input_bam
-O $recalibration_report_filename
-knownSites "Homo_sapiens_assembly38.dbsnp138.vcf"
-knownSites "Mills_and_1000G_gold_standard.indels.hg38.vcf.gz"
-knownSites "Homo_sapiens_assembly38.known_indels.vcf.gz"
```

**Optional flags**

```
-rf BadCigar
-preserve_qscores_less_than 6
-disable_auto_index_creation_and_locking_when_reading_rods
-disable_bam_indexing
-nct
-useOriginalQualities
-downsample_to_fraction 0.1
-L chr1 -L chr2 -L chr3 -L chr4 -L chr5 -L chr6 -L chr7 -L chr8 -L chr9 -L chr10 -L
chr11 -L chr12 -L chr13 -L chr14 -L chr15 -L chr16 -L chr17 -L chr18 -L chr19 -L chr20
-L chr21 -L chr22
```
(For efficiency)

#### Base Quality Score Binning Scheme

To reduce file size, we recommend four-bin quality score compression using ApplyBQSR with:

**Software: ApplyBQSR**

**Standardized flags**

```
-SQQ 10 -SQQ 20 -SQQ 30
-I $input_bam
-O $output_bam
-bqsr $recalibration_report
-R $ref_fasta
-disable_indel_quals
```

**Optional flags**

```
-globalQScorePrior -1.0
-preserve_qscores_less_than 6
-useOriginalQualities
-nct
-rf BadCigar
-createOutputBamMD5
-addOutputSAMProgramRecord
```

##### Haplotype Caller

Variant calling should be performed using GATK HaplotypeCaller. Resulting `.gvcf` files are used for joint discovery. Recommended flags for this step include:

**Software: HaplotypeCaller**

**Standardized flags**

```
-R $ref_fasta
-I $input_bam
-L $interval_list
-O $output_filename
-contamination 0
-ERC GVCF
-G StandardAnnotation
-G StandardHCAnnotation
-G AS_StandardAnnotation
```

#### GVCF Postprocessing

We recommend postprocessing GVCFs using GATK ReblockGVCF to reduce file size and runtime. This sacrifices genotype quality precision at low coverage sites but does not significantly impact downstream analyses. Suggested flags:

**Software: ReblockGVCF**

| **Standardized flags** |
|---|
| `-drop-low-quals` |
| `-do-qual-approx` |
| `-floor-blocks -GQB 10 -GQB 20 -GQB 30 -GQB 40 -GQB 50 -GQB 60` |

#### Joint Discovery and VQSR Filtering

When preparing joint calling and VQSR filtering scripts, we recommend coding indels to be run first because of the nature of indels being few compared to SNPs. Therefore the indel run would be completed first before the SNP model. Suggested tools and associated flags are as follows:

**Software: GenomicsDBImport** Note that if joint calling is required for more than 50,000 (exomes) or 5,000 (genomes) samples, we recommend Hail's vds combiner[11].

| **Standardized flags** |
|---|
| `-batch-size 50` |
| `-L $interval_list` |
| `-reader-threads` 5 |
| `-merge-input-intervals` |
| `-consolidate` |

**Software: GenotypeGVCFs**

**Standardized flags**

-R $ref_fasta

-O $output_vcf_filename

-D $dbsnp_vcf

-G StandardAnnotation

-only-output-calls-starting-in-intervals

-use-new-qual-calculator

-V $workspace

-L $interval

-merge-input-intervals

**Software: VariantRecalibrator**

**Standardized flags**

-V $sites_only_variant_filtered_vcf

-O $recalibration_filename

-tranches-file $tranches_filename

-trust-all-polymorphic

-tranche $sep=' -tranche ' recalibration_tranche_values

-an $sep=' -an ' recalibration_annotation_values

-mode SNP (This tool should be run once for SNPs and once for indels)

-sample-every-Nth-variant $downsampleFactor (Needed for samples higher than ~150,000 exomes /~15,000 genomes)

-output-model $model_report_filename

-max-gaussians 6 (Conceptually, this flag will help with modeling variation between different sequencing platforms. It may need further tweaking on user end)

-use-allele-specific-annotations

-resource:hapmap,known=false,training=true,truth=true,prior=15 $hapmap_resource_vcf

-resource:omni,known=false,training=true,truth=true,prior=12 $omni_resource_vcf

-resource:1000G,known=false,training=true,truth=false,prior=10 $one_thousand_genomes_resource_vcf

-resource:dbsnp,known=true,training=false,truth=false,prior=7 $dbsnp_resource_vcf

**Software: ApplyVQSR**

**Standardized flags**

`-O $recalibrated_vcf_filename`

`-V tmp.indel.recalibrated.vcf`

`-recal-file $snps_recalibration`

`-tranches-file $snps_tranches`

`-truth-sensitivity-filter-level $snp_filter_level`

`-create-output-variant-index true`

`-mode SNP` This tool should be run once for SNPs and once for indels.

`-use-allele-specific-annotations`

Resulting .vcf files can then be used as input to subsequent QC described in the sample and variant quality control section of the methods.

#### Sample and Variant Quality Control

These guidelines were circulated to all analysts in the consortium to preparation of an analysis callset for the primary BRaVa discovery analysis.

##### WES Interval QC

Optionally filter to intervals where at least 85% of samples have mean coverage ≥ 20×. Failing intervals may be flagged as `fail_interval_qc`.

##### Genotype QC

Genotypes should be removed based on genotype quality, depth, and allele balance thresholds specific to zygosity class.

Remove the following:

- If homozygous reference, at least one of:
  - Genotype quality < 20
  - Depth < 10
- If heterozygous, at least one of:

    - (Reference allele depth + alternative allele depth)/depth < 0.8
    - (Alternative allele depth)/depth < 0.2
    - Reference phred-scaled genotype posterior < 20
    - Depth < 10
- If homozygous variant, at least one of:
    - (Alternative allele depth)/depth < 0.8
    - Reference phred-scaled genotype posterior < 20
    - Depth < 10

##### Initial Variant QC

Variants should be removed if they:

- Fall in low-complexity regions
- Fail VQSR
- Fall outside padded target intervals (50 bp)
- Are invariant

##### Initial Sample QC

Samples should be removed based on thresholds determined from empirical distributions of sample call rate, mean depth, mean genotype quality, contamination, and chimerism where available. The last two metrics can be extracted from the GATK/Picard metadata.

##### Create high quality common variant subset to determine ancestry and remove sex-swaps

Export common variants (allele frequency between 0.01 to 0.99) with high call rate (> 0.98) to and prune to pseudo-independent SNPs, e.g. using `-indep 50 5 2` in plink[12].

##### Impute sexes of the individuals

Impute the sexes of sample with this pruned set of variants on the X, and create a list of samples with incorrect or unknown sex as defined by:

- Sex is 'unknown' in the phenotype files.
- $F$-statistic > 0.8 and the sex is 'female' in the phenotype file.
- $F$-statistic < 0.2 and the sex is 'male' in the phenotype file.

We recommend plotting the resultant output to check that the data sit within the above thresholds.

In order to refine inferred sex, take each sample's fraction of chromosome Y coverage, normalised by the sample's chromosome 20 coverage. Aneuploidies can be determined in samples that impute female but have normalised chrY fraction $> 0.1$, and those that impute male but have normalised chrY fraction $< 0.1$. If sex was imputed as missing, set sex to ambiguous.

##### Determine relatedness between samples

Compute IBD between all pairs of individuals using the pruned set of variants in the autosome, and create a sample list of individuals such that no pair exceeds $\hat{\pi} > 0.2$.

##### Define ancestry labels using either gnomAD, 1000G, and/or HGDP

For WGS data, use the gnomAD SNP loadings to project samples and train a random forest classifier.

If imputed genotype data is available for the same samples, more accurate clustering will be achieved by filtering 1000G/HGDP for HM3 SNPs, LD-pruning, running PCA on the 1000G/HGDP samples using these variants, and projecting in the samples from the dataset under analysis.

If only exome data is available, it will be more straightforward to intersect 1000G/HGDP variants with the collection of high quality common variants evaluated in 'Create high quality common variant subset to determine ancestry and remove sex-swaps', and LD-prune. Ensure that you ~10,000s of SNPs are available following LD pruning, otherwise the resultant clustering will be poor.

Run PCA on unrelated 1000G/HGDP after variant filtering above, and project in the samples using the SNP loadings.

Train a random forest on the super populations of 1000G/HGDP genomes and predict super populations of samples in the cohort under analysis.

##### Final Variant QC

Within ancestry-stratified unrelated samples, variants should be removed if call rate $< 0.97$ or Hardy–Weinberg equilibrium $P < 1 \times 10^{-10}$ (following removal of related samples defined earlier).

##### Final Sample QC

Outliers should be removed using median absolute deviation thresholds for the following sample level metrics:

- Number of SNP alternate alleles
- Transition/transversion ratio
- Insertion/Deletion allele ratio
- Number of insertion alternate alleles
- Number of deletion alternate alleles
- Heterozygous/homozygous call ratio

#### Annotation definitions for gene-level association testing

We annotated genetic variants using the Ensembl Variant Effect Predictor (VEP) version 105[13], with the LOFTEE plugin against genome build GRCh38 (v1.04_GRCh38)[14]. The MANE select transcript was annotated and assigned as the 'canonical' transcript. For consequence annotations, we assign with respect to the MANE select transcript, and the small number of 'canonical' transcripts in genes without a defined MANE select transcript. For any putative loss of function (LoF) variants, we annotated with LOFTEE (HC, LC, and flags). We used a series of *in silico* deleteriousness metrics to define a collection of broad annotation categories for our gene-based association tests: CADD (v1.6)[15], REVEL[16], using dbNSFP4.3[17], and spliceAI v1.3[18]. Following annotation with each of these metrics and consequence with VEP, we applied the following rules to assign a single variant category for each variant:

1. **High confidence pLoF:** high-confidence LoF variants (LOFTEE HC)
2. **Damaging missense/protein-altering:** any variant not categorised in (1) (High confidence pLoF) with at least one of
   (a) Variant annotated as missense/start-loss/stop-loss/in-frame indel and (REVEL $\geq 0.773$ or CADD $\geq 28.1$ (or both))
   (b) Any variant with SpliceAI DS $\geq 0.2$ where SpliceAI DS is max (DS_AG, DS_AL, DS_DG, DS_DL).
   (c) Low-confidence LoF variants (LOFTEE LC)
3. **Other missense/protein-altering:**
   (a) Missense/start-loss/stop-loss/in-frame indel not categorised in (2) (Damaging missense/protein-altering).

4. **Synonymous:** synonymous variants with SpliceAI DS < 0.2 in the gene (control set).

Lower numbers in the categorization take precedence, so e.g. if a variant is categorized as HC by LOFTEE and has a SpliceAI DS score ≥ 0.2, it should receive a 'high confidence pLoF' annotation. Note also that if a variant has multiple consequence annotations with respect to the MANE select transcript, it receives the most deleterious among them. REVEL and CADD cutoffs chosen are based on the PP3 moderate criteria[19]. Code to annotate according to these criteria, pinned to the specified versions to ensure consistency, and create group-files ready for input into SAIGE is available in our variant annotation github repository[20].

#### Association testing

Association testing Using the annotations defined in the previous section, we carried out association testing at the variant and gene-level using Scalable and Accurate Implementation of GEneralized mixed model (SAIGE), scalable genetic association testing software that allows for relatedness, case-control imbalance of binary traits, and the robust analysis of rare variation. Binary traits were analyzed if both the number of cases and controls exceeded 100. Covariates included in the both-sex analyses were *age*, $age^2$, *sex*, $age \times sex$, $age^2 \times sex$ and *20 genetic PCs*. For sex-specific analyses, we included *age*, $age^2$, and *20 genetic PCs*. SAIGE analysis consists of three steps. We provided a docker container pinned to SAIGE 1.3.6 and provided scripts hard-coding the options described below for each step of the analysis pipeline[21].

##### Step 0: Compute the GRM for the mixed effects term

Biobanks first extracted an LD-pruned set of variants from genotyping array data or WGS data, which were then passed to createSparseGRM.R in SAIGE with the following options:

- `-numRandomMarkerforSparseKin=5000`
- `-relatednessCutoff=0.05`

In addition, biobanks constructed a set of plink files containing randomly chosen variants above and below MAC 20, which were used to estimate the 'variance ratio' in the SAIGE analysis.

##### Step 1: Fit the null generalized linear model (linear or logistic for continuous and binary traits, respectively)

Next, biobanks fit the null model using step1_fitNULLGLMM.R in SAIGE, with the following flags:

- `-useSparseGRMtoFitNULL TRUE`
- `-skipVarianceRatioEstimation FALSE`

- `-invNormalize TRUE` (for quantitative traits)
- `-isCateVarianceRatio TRUE`

##### Step 2: Perform association testing for each variant or set of variants (burden and variance based tests)

**Gene level** We carried out group-based tests using SAIGE-gene+ for each gene, with 1%, 0.1%, and 0.01% MAF cutoffs for each phenotype (`-maxMAF_in_groupTest=0.0001, 0.001, 0.01`). Using the annotations defined above, we performed group-based testing for each gene using the following annotations using the `-annotation_in_groupTest` flag:

- High confidence pLoF
- Damaging missense/protein-altering
- Other missense/protein-altering
- Synonymous
- High confidence pLoF or damaging missense/protein-altering

Other flags we set for group-based testing were:

- `-minMAF 0`
- `-minMAC 0.5`
- `-LOCO FALSE`
- `-is_Firth_beta TRUE`
- `-pCutoffforFirth 0.1`
- `-is_output_moreDetails TRUE`
- `-is_fastTest TRUE`
- `-is_output_markerList_in_groupTest TRUE`
- `-is_single_in_groupTest TRUE`

These parameters support group-based association testing (Burden, SKAT, and SKAT-O) across the defined phenotypes in five broad continental ancestry groups (AFR, AMR, EAS, EUR, SAS), using up to four functional variant classes defined in the annotation section (High confidence pLoF, damaging missense/protein-altering, other missense/protein-altering, synonymous) and three maximum minor allele frequency thresholds (0.01, 0.001, and 0.0001) following broad ancestry grouping. Variant level Variant level test statistics for all variants with MAF $< 1\%$ were generated through the `-is_single_in_groupTest TRUE` flag applied when performing gene level association testing described above. Throughout each of the three steps, anything not mentioned in the above collection of flags was set at the default value in SAIGE. We enforce the choice of 'optional' flags through a series of scripts for each step, which hard-code the flags and wrap the relevant SAIGE function. Our github repository, universal-saige[21], provides a README and step-by-step walkthroughs.

#### Summary statistic quality control

Following association testing we performed a series of automated investigations of inflation, removing any (biobank, phenotype) results with synonymous $\lambda_{95}$ or $\lambda_{99} > 1.3$ from subsequent meta-analysis, as well as manual inspection of QQ plots for additional removal. A summary plot of the genomic inflation statistics split by biobank is displayed in Fig. S6. In gene-based association testing, a small subset of association analyses, we observed widespread inflation. This was driven by the use of `-isCovariateOffset` leading to spurious inflation in edge cases. Turning off this option (which we use for computational efficiency) removed the inflation of the test statistics. The reason for this is that when the `-isCovariateOffset` flag is used, SAIGE fits a null model without random effects (no sample relatedness accounted for), in which fixed effect coefficients of covariates are estimated. These estimates are then included in the null random effect model as an offset, rather than iteratively updating them as random effects are estimated. This approach has been used in several biobank-scale GWAS methods to increase computational efficiency. However, the simplified model fitting procedure ignores covariance between covariates (fixed effects) and random effects. In the small number of case where such inflation was observed, biobanks reran the analysis with the `-isCovariateOffset` flag set to false.

In the Genes & Health cohort, we compared results from the SAIGE-BRaVa pipeline to those obtained from an independent pipeline for both single-variant and gene burden testing run by the core Genes & Health team using REGENIE software. Results were similar; for example, the top five single variant and gene-based hits for Type 2 Diabetes were the same in both pipelines.

#### Cross-ancestry comparison of gene-level burden effect sizes

To assess the consistency of gene-level rare variant effect size estimates across genetic ancestries, we compared ancestry-specific burden effect sizes to those obtained from European (EUR) meta-analysis.

#### Data preparation

Gene-level association summary statistics were obtained from BRaVa meta-analyses across all contributing biobanks. Analyses were restricted to burden tests using inverse-variance weighted meta-analysis. For each phenotype, we identified (gene, phenotype, mask) tupes with $P < 0.05/(20,000 \cdot 2 \cdot 3) = 4.2 \times 10^{-7}$ in the EUR meta-analysis and used these to define the set of associations included in downstream comparisons.

For each phenotype, corresponding gene-level summary statistics were extracted from ancestry-specific meta-analyses (AFR, AMR, EAS, SAS, and non-European combined analyses). Summary statistics were harmonized across datasets by matching on gene (Region), annotation mask (Group), minor allele frequency threshold (max MAF), and phenotype. Extracted variables included burden effect sizes, standard errors, heterogeneity statistics, and meta-analysis weights. Phenotypes were classified as binary (case–control) or continuous traits based on predefined phenotype groupings.

#### Deming regression analysis

To quantify concordance between ancestry-specific and EUR effect size estimates, we performed Deming regression, which accounts for measurement error in both variables. For each combination of:

- Phenotype type (binary vs continuous),
- Ancestry group (AFR, AMR, EAS, SAS, non-European),
- Variant annotation mask (pLoF, damaging missense/protein-altering [DM/PA], and combined pLoF;DM/PA),
- Maximum minor allele frequency threshold (0.1% and 0.01%),

we fitted a Deming regression model of the form:

$$\beta_{\text{ancestry}} = \alpha + \gamma \cdot \beta_{\text{EUR}} + \varepsilon,$$

where $\beta_{\text{EUR}}$ denotes the effect size from the European meta-analysis, and $\beta_{\text{ancestry}}$ denotes the corresponding ancestry-specific estimate.

Measurement error in both variables was accounted for by incorporating the reported standard errors from the burden tests. For each model, we estimated the slope, intercept, and their standard errors. Ninety-five percent confidence intervals were calculated assuming asymptotic normality.

To ensure stability of estimates, analyses were restricted to strata with at least 40 gene–phenotype pairs.

We tested whether the regression slope differed from zero, corresponding to evidence for a non-zero relationship between ancestry-specific and EUR effect size estimates.

We then applied a Bonferroni correction across the remaining tests ($n = 48$) to define significance, corresponding to all combinations of ancestry group, phenotype type, variant annotation mask, and minor allele frequency threshold that met inclusion criteria (at least 40 gene–phenotype pairs per stratum).

Results are summarized in Table S11.

#### Meta-analysis

We performed fixed effects meta-analysis using weighted inverse-variance weighted meta-analysis for variant and burden testing at the gene-level, and Stouffer's method for variance and hybrid gene-level testing (SKAT and SKAT-O), weighting by effective sample size. (biobank, ancestry) tuples for a given phenotype were included in the meta-analysis if they contained at least 100 cases. For a given (trait, gene, annotation) tuple, we meta-analyzed across (biobank, genetic ancestry) using Stouffer's method, computing weighted $Z$-scores for each (biobank, genetic ancestry):

$$\widetilde{Z}_{b,g} = \sqrt{N_{\mathrm{eff},b,g}}\Phi^{-1}\left(1 - P_{b,g}\right),$$

where, $N_{\mathrm{eff},b,g}$ is the effective sample size of genetic ancestry $g$ in biobank $b$, and $P_{b,g}$ is the associated $P$-value. These weighted $Z$-scores are then combined into an overall $Z$-score:

$$Z = \frac{\sum_{b,g} \widetilde{Z}_{b,g}}{\sqrt{\sum_{b,g} N_{\mathrm{eff},b,g}}}.$$

The overall $P$-value for meta-analyzed SKAT and SKAT-O association tests is then

$$P = 1 - \Phi\left(|Z|\right).$$

The effective sample sizes were estimated by relatedness using the sparse GRM defined in step 0 of the association testing pipeline. To estimate phenotype specific effective population size estimates we used the extractNglmm.R script in the SAIGE repository. Briefly, the approach uses preconditioned conjugate gradients to estimate effective sample size under the (sparse) GRM-induced correlation. For inverse-variance weighting of gene-level burden test statistics we combined gene-level association test statistics for each (trait, gene, annotation):

$$P = 1 - 2\Phi\left(|Z|\right), \quad Z = \frac{\sum_{b,g} \frac{\beta_{b,g}}{SE_{b,g}^2}}{\sqrt{\sum_{b,g} \frac{1}{SE_{b,g}^2}}}.$$

We carried out a series of meta-analyses, across all continuous and disease traits with:

1. All biobanks across all genetic ancestries.

2. All biobanks stratified by each genetic ancestry label (AFR, AMR, EAS, EUR, SAS).

3. All biobanks restricted to non-EUR genetic ancestry labels.

4. Leave one-biobank out meta-analyses.

In each case, meta-analysis was carried out across all available (biobank, genetic ancestry) pairs.

To determine $N_{\text{eff},b,g}$ for each (biobank, genetic ancestry) pair, we used a null generalized linear mixed model framework with genetic relatedness accounted for via a covariance matrix derived from the genotype data used to fit the mixture component in SAIGE runs. For each phenotype, we first fit a null model including specified covariates (and optional transformations such as inverse-normalization for quantitative traits and QR-based orthogonalization of covariates). Sample inclusion was restricted to individuals with both genotype and phenotype data, with optional sex-specific analyses.

To avoid explicit construction or inversion of the full covariance matrix, we employed a preconditioned conjugate gradient (PCG) algorithm to solve linear systems of the form $\Sigma x = \mathbf{1}$, thereby obtaining $\Sigma^{-1}\mathbf{1}$, where $\Sigma$ represents the model-implied covariance structure incorporating genetic relatedness. PCG is an iterative method suitable for large, sparse, symmetric positive-definite matrices, and enables efficient approximation of matrix–vector products without direct matrix inversion.

From the PCG solution, we computed the scalar quantity $\mathbf{1}^T\Sigma^{-1}\mathbf{1}$, which reflects the effective information content of the sample under the fitted model. For binary traits, the effective sample size was further scaled by $4p(1-p)$, where $p$ is the proportion of cases in the sample, yielding

$$N_{\text{eff}} = 4p(1-p)\,\mathbf{1}^T\Sigma^{-1}\mathbf{1}.$$

For quantitative traits, the effective sample size was defined as

$$N_{\text{eff}} = \mathbf{1}^T\Sigma^{-1}\mathbf{1}.$$

We used a sparse genetic relationship matrix (GRM) to improve computational efficiency. Convergence of the PCG algorithm was controlled via predefined tolerance and iteration limits.

To assess the impact of assuming independence of samples within each cohort on resultant association test statistics, we compared this estimate of $N_{\text{eff}}$ to that obtained under an independence assumption, namely:

$$N_{\text{eff}} = N$$

for continuous traits, and

$$N_{\text{eff}} = \frac{4}{\frac{1}{N_{\text{case}}} + \frac{1}{N_{\text{control}}}}.$$

Results are displayed in Fig. S11.

#### Cumulative allele frequency weighted Stouffer's meta-analysis for SKAT and SKAT-O

As a sensitivity analysis, we performed meta-analysis of SKAT and SKAT-O gene-level association results using Stouffer's method that also incorporates combined allele frequency (CAF) weighting.

$$\widetilde{Z}_{\text{CAF},b,g} = \sqrt{2\ \text{CAF}_{b,g}(1 - \text{CAF}_{b,g})\ N_{\text{eff},b,g}}\ \Phi^{-1}\left(1 - P_{b,g}\right),$$

where ($N_{\text{eff},b,g}$) is the effective sample size and ($P_{b,g}$) is the $P$-value for the corresponding SKAT or SKAT-O test. The CAF term, ($\text{CAF}_{b,g}$), denotes the combined allele frequency of variants included in the corresponding gene–annotation group for ancestry ($g$) within biobank ($b$).

These weighted $Z$-scores were then combined across (biobank, genetic ancestry) pairs as:

$$Z_{\text{CAF}} = \frac{\sum_{b,g} \widetilde{Z}_{\text{CAF},b,g}}{\sqrt{\sum_{b,g} 2\ \text{CAF}_{b,g}(1 - \text{CAF}_{b,g})\ N_{\text{eff},b,g}}}.$$

The corresponding meta-analyzed $P$-value was computed as:

$$P = 1 - \Phi\left(|Z_{\text{CAF}}|\right).$$

For some biobanks, ancestry-specific CAF values could not be directly shared. In these instances we used proxy estimates derived from available summary statistics or external reference panels. In particular, for each genetic ancestry where CAF information was available, we used the largest such cohort to define reference CAFs for each (gene, annotation) pair, which we use for weights in the meta-analysis. In instances where a (gene, annotation) result is present, but an estimate of CAF is not available, we assume there is a singleton in that (gene, annotation) tuple. We note that the variance approximation $2\text{CAF}_{b,g}(1 - \text{CAF}_{b,g})$ is reasonable when the constituent variants comprising the CAF are low in frequency, but breaks down for very large genes, larger max MAF cutoffs, or variant annotation categories that comprise a large number of variants (e.g. the union of pLoF, damaging missense or protein altering, other missense or protein altering, and synonymous). In these instances, it is possible to obtain a negative variance approximation. To allow for reasonable variance estimates across CAFs, we used a collection of approximations, dependent of the CAF. First, define $\lambda = 2$ CAF and let $n_{\text{rare}}$ and $n_{\text{ultra}}$ denote the number of rare (MAC > 10) and ultra-rare (MAC ≤ 10) variants within the mask, respectively (available in our SAIGE-gene output).

**Sparse regime.** For $\lambda < 0.05$, use a Bernoulli approximation, as described above (the majority of cases):

$$\text{Var} \approx 2\ \text{CAF}\ (1 - \text{CAF})$$

**Intermediate regime with rare variants.** For $0.05 \leq \lambda < 0.2$ and $n_{\text{rare}} > 0$, assume that the total allele frequency is distributed approximately evenly across rare variants, implying:

$$\text{Var} \approx 2 \left( \text{CAF} - \frac{\text{CAF}^2}{n_{\text{rare}}} \right).$$

**General case (mixture of rare and ultra-rare variants).** Otherwise, split the cumulative allele frequency into contributions from rare and ultra-rare variants:

$$\text{CAF}_{\text{rare}} = \text{CAF}\ \frac{n_{\text{rare}}}{n_{\text{rare}} + n_{\text{ultra}}}, \quad \text{CAF}_{\text{ultra}} = \text{CAF}\ \frac{n_{\text{ultra}}}{n_{\text{rare}} + n_{\text{ultra}}},$$

and model the rare component as a sum of independent contributions:

$$\text{Var}_{\text{rare}} \approx 2 \left( \text{CAF}_{\text{rare}} - \frac{\text{CAF}_{\text{rare}}^2}{n_{\text{rare}}} \right),$$

Finally, include the ultra-rare component using a Poisson approximation for carrier status:

$$c = 1 - \exp(-2\ \text{CAF}_{\text{ultra}}), \quad \text{Var}_{\text{ultra}} \approx c\ (1 - c).$$

The total variance for the mixture is then estimated by the sum of these contributions: $\text{Var} \approx \text{Var}_{\text{rare}} + \text{Var}_{\text{ultra}}$.

#### Caveats of METAL sample overlap correction for rare variant analyses

There are a collection of practical limitations of the sample overlap correction implemented in METAL (using `OVERLAP ON`), which are particularly relevant in the context of rare variant or gene-based analyses with differential variant missingness across cohorts.

First, METAL implements overlap correction using a iterative meta-analysis scheme, in which studies are incorporated one at a time. As a consequence, the resulting estimates depend on the order in which studies are provided if there is differential variant missingness between them (Fig. S13-S14). This occurs because, at each step, the method updates correlation estimates and effective sample size based on the subset of variants shared between the current aggregate and the next study. When variant availability differs across cohorts (which will almost certainly occur for rare variants) this induces a progressively changing set of variants contributing to the overlap estimation, such that later-added studies are conditioned on a different subset of variants than earlier ones.

This leads to two related issues:

- The overlap correction is not symmetric across studies, and results may vary depending on input ordering.
- The procedure implicitly relies on variant intersections that shrink or change across iterations, which can distort correlation estimates and downstream effective sample size calculations when missingness is non-uniform.

Second, for rare variant analyses, we observed that the estimation of cross-study correlation (used for overlap correction) can become biased. This is partly because many variants (or genes) are effectively underpowered, yielding test statistics that deviate from the assumed null behavior (Fig. S15). METAL estimates cross-study correlation using variants with $|Z| < z_{\text{cutoff}}$ (1 by default), implicitly assuming that this subset is approximately drawn from the null and that, under the null, test statistics follow a standard normal distribution: $N(0, 1)$. It then estimates the correlation parameter using a truncated bivariate normal likelihood based on these $Z$-scores. However, in settings with low case counts and small gene sizes, the true null distribution can deviate substantially from the asymptotic $N(0, 1)$ approximation due to discreteness of the underlying $2 \times 2$ contingency tables and case–control imbalance. As a result, even under the null, test statistics may display non-zero mean or skewness (Fig. S15).

Because the overlap estimator is based on the joint distribution of $Z$-scores within this truncated region, it depends critically on these assumptions. When they are violated, deviations from mean-zero calibration can induce spurious positive correlation across cohorts even in the absence of true sample overlap (since $E[Z_1 Z_2] = \text{Cov}(Z_1, Z_2) + E[Z_1]E[Z_2]$). This leads to inflated overlap estimates, reduced $N_{\text{eff}}$, and a corresponding loss of statistical power.

Third, the current implementation in METAL uses a sub-optimal weighting scheme for overlap correction rather than the optimal generalized least squares solution (Methods: Accounting for overlap in fixed effects meta-analysis). While METAL's approach preserves type I error by correctly adjusting the variance of the standard sample-size-weighted sum, it fails to apply the optimal inverse-covariance weights to the study estimates themselves. This approximation leads to statistical inefficiency and mis-estimation of the effective sample size, ultimately resulting in a loss of power compared to the optimal matrix formulation.

As a result, METAL's overlap correction should be applied with caution in settings involving rare variants, gene-based tests, or substantial cross-cohort differences in variant inclusion. In particular, sensitivity to study ordering and instability of correlation estimates can materially affect results.

#### Accounting for overlap in fixed effects meta-analysis

For study $i$, let

$$
\begin{aligned}
N_i &: \text{ effective sample size,} && (1)\\
\beta_i &: \text{ effect size estimate,} && (2)\\
\mathrm{SE}_i &: \text{ standard error,} && (3)\\
P_i &: \ P\text{-value,} && (4)\\
\Delta_i &: \text{ direction of effect,} && (5)\\
Z_i &= \frac{\beta_i}{\mathrm{SE}_i}. && (6)
\end{aligned}
$$

Finally, let $R$ denote the correlation matrix of $Z$-scores across studies.

#### Sample size based meta-analysis

*Inputs:*

$$N_i,\ P_i,\ \Delta_i. \tag{7}$$

*Intermediates:*

$$Z_i = \Phi^{-1}\left(\frac{P_i}{2}\right)\Delta_i, \tag{8}$$

$$w_i = \sqrt{N_i}. \tag{9}$$

*No sample overlap:*

$$N = \sum_i w_i^2, \quad Z = \frac{\sum_i Z_i w_i}{\sqrt{N}}, \quad P = 2\Phi(-|Z|). \tag{10}$$

Cochran's $Q$ statistic: $Q = \sum_i Z_i^2 - Z^2$. *Sample overlap:*

$$N = \sum_{i,j} w_i R_{i,j}^{-1} w_j, \quad Z = \frac{\sum_{i,j} Z_i R_{i,j}^{-1} w_j}{\sqrt{N}}, \quad P = 2\Phi(-|Z|). \tag{11}$$

Cochran's $Q$ statistic: $Q = \sum_{i,j} Z_i R_{i,j}^{-1} Z_j - Z^2$.

##### Inverse Variance Based Meta-analysis

*Inputs:*

$$\beta_i,\ \mathrm{SE}_i \tag{12}$$

*Intermediates:*

$$Z_i = \frac{\beta_i}{\mathrm{SE}_i},\ w_i = \frac{1}{\mathrm{SE}_i}. \tag{13}$$

*No overlap:*

$$N = \sum_i w_i^2, \quad \beta = \frac{\sum_i \beta_i w_i^2}{N}, \quad \mathrm{SE} = \frac{1}{\sqrt{N}}, \quad Z = \frac{\beta}{\mathrm{SE}}, \quad P = 2\Phi(-|Z|). \tag{14}$$

*Sample overlap:*

$$N = \sum_{i,j} w_i R_{i,j}^{-1} w_j, \quad \beta = \frac{\sum_{i,j} \beta_i w_i R_{i,j}^{-1} w_j}{N}, \quad \mathrm{SE} = \frac{1}{\sqrt{N}}, \quad Z = \frac{\beta}{\mathrm{SE}} \quad P = 2\Phi(-|Z|). \tag{15}$$

The formulae implemented in `METAL` do not use the optimal inverse-covariance weights suggested by Lin and Sullivan (2009). Assuming $\mathrm{Var}(Z_1) = \mathrm{Var}(Z_2) = 1$ and $\mathrm{Cov}(Z_1, Z_2) = r_{12}$, both approaches yield $\mathrm{Var}(Z) = 1$.

#### Determining optimal weights when there is sample overlap

Following Lin and Sullivan, suppose that there are K studies with potentially overlapping subjects. For $k = 1, \ldots, K$, let $\hat{\eta}_k$ be the estimator of a common genetic effect $\eta$ from the $k$th study. For meta-analysis of $K$ studies with sample overlap the optimal weights for the meta-analyzed test statistic $\hat{\eta} = \sum_{k=1}^{K} a_k \hat{\eta}_k$ are

$$(a_1, \ldots, a_K)^\top = \frac{\Omega^{-1}\mathbf{1}}{\mathbf{1}^\top \Omega^{-1} \mathbf{1}}. \tag{16}$$

where $\mathbf{1}$ is a $K \times 1$ vector of 1's and $\Omega$ is the (estimated) covariance matrix of $(\hat{\eta}_1 \ldots \hat{\eta}_k)$. For us,

$$\hat{\eta}_k = \frac{Z_k}{\sqrt{N_k}}. \tag{17}$$

We wish to determine these weights as a function of available information. First, consider the entries of $(a_1, \ldots, a_K)^\top$. The denominator,

$$\mathbf{1}^\top \Omega^{-1} \mathbf{1} = \sum_{i,j} \Omega_{ij}^{-1} \tag{18}$$

is simply the sum of all entries of $\Omega^{-1}$. We can write $\Omega$ as

$$\Omega = \mathrm{Cov}\left(\frac{Z_1}{\sqrt{N_1}}, \ldots, \frac{Z_K}{\sqrt{N_K}}\right). \tag{19}$$

Then

$$\Omega = D^{-1/2} R D^{-1/2}, \tag{20}$$

where $D = \mathrm{diag}(N_1, \ldots, N_K)$.

Hence,

$$\Omega^{-1} = D^{1/2} R^{-1} D^{1/2}. \tag{21}$$

Now, letting $w_i = \sqrt{N_i}$,

$$\sum_{i,j} \Omega^{-1}_{ij} = \sum_{i,j} w_i w_j R^{-1}_{ij}. \tag{22}$$

So $a_k$ is

$$\frac{\sum_j w_k w_j R^{-1}_{k,j}}{\sum_{i,j} w_i w_j R^{-1}_{i,j}}. \tag{23}$$

Now, how do we obtain the meta-analysis $Z$-score, and $\sqrt{N}$?

Set

$$\begin{aligned}
\frac{Z}{\sqrt{N}} &= \sum_{k=1}^{K} \frac{Z_k}{\sqrt{N_k}} a_k \qquad (24)\\
&= \sum_{k=1}^{K} \frac{Z_k}{\sqrt{N_k}} \frac{\sum_j w_k w_j R^{-1}_{k,j}}{\sum_{i,j} w_i w_j R^{-1}_{i,j}} \qquad (25)\\
&= \frac{\sum_{k=1}^{K} Z_k \sum_j w_j R^{-1}_{k,j}}{\sum_{i,j} w_i w_j R^{-1}_{i,j}}. \qquad (26)
\end{aligned}$$

Now, let's determine $\sqrt{N}$ using the fact that $\mathrm{Var}(Z) = 1$. $Z$ is of the form

$$Z = \sqrt{N} \sum_k Z_k b_k, \tag{27}$$

where $b_k = \frac{\sum_j w_j R^{-1}_{kj}}{\sum_{i,j} w_i w_j R^{-1}_{ij}}$.

$$\mathrm{Var}(Z) = \mathbb{E}\left[Z^2\right] - \mathbb{E}\left[Z\right]^2 = \mathbb{E}\left[Z^2\right]. \tag{28}$$

Expanding,

$$\mathrm{Var}(Z) = N\mathbb{E}\left[\sum_{k,\ell} Z_k Z_\ell b_k b_\ell\right] \tag{29}$$

$$= N \sum_{k,\ell} R_{k,\ell} b_k b_\ell. \tag{30}$$

Substituting in for $b_k$,

$$\mathrm{Var}(Z) = \frac{N \sum_{k,\ell} R_{k,\ell} \left(\sum_j w_j R^{-1}_{k,j}\right) b_\ell}{\sum_{i,j} w_i w_j R^{-1}_{i,j}} \tag{31}$$

$$= \frac{N \sum_{\ell} \left(\sum_j w_j \left(\sum_k R^{-1}_{j,k} R_{k,\ell}\right)\right) b_\ell}{\sum_{i,j} w_i w_j R^{-1}_{i,j}}. \tag{32}$$

The innermost sum is 0 if $j \neq \ell$ and 1 if $j = \ell$, so we can rewrite as

$$\mathrm{Var}(Z) = \frac{N \sum_\ell w_\ell b_\ell}{\sum_{i,j} w_i w_j R^{-1}_{i,j}} \tag{33}$$

$$= \frac{N \sum_\ell w_\ell \sum_i w_i R^{-1}_{i,\ell}}{\left(\sum_{i,j} w_i w_j R^{-1}_{i,j}\right)^2} = \frac{N \sum_{i,j} w_i w_j R^{-1}_{i,j}}{\left(\sum_{i,j} w_i w_j R^{-1}_{i,j}\right)^2} = \frac{N}{\sum_{i,j} w_i w_j R^{-1}_{i,j}}. \tag{34}$$

Now, since $\mathrm{Var}(Z) = 1$, we can rearrange for $N$.

$$N = \sum_{i,j} w_i w_j R^{-1}_{i,j}. \tag{35}$$

So, the combined statistic is

$$\frac{Z}{\sqrt{N}} = \frac{\sum_{k=1}^{K} Z_k \sum_{j=1}^{K} w_j R^{-1}_{k,j}}{\sum_{i,j} w_i w_j R^{-1}_{i,j}}. \tag{36}$$

Overall, we have

$$w_i = \sqrt{N_i}, \tag{37}$$

$$N = \sum_{i,j} w_i w_j R^{-1}_{i,j}, \tag{38}$$

$$Z = \sum_{k=1}^{K} Z_k \frac{\sum_j w_j R^{-1}_{kj}}{\sqrt{N}}. \tag{39}$$

#### Locus definition for variant-level results

Loci were defined for each phenotype and chromosome using a distance-based clustering procedure. For each significant variant association ($P < 8.0 \times 10^{-9}$) with genomic position $p$, we considered the genomic interval $[p - w,\ p + w]$, where $w = 500{,}000$ base pairs. Intervals were truncated at the lower bound of 1 bp and the upper bound of the respective chromosome length where applicable.

Within each chromosome, all such intervals were merged by taking the union of overlapping intervals. The resulting merged intervals defined our locus boundaries, with start and end coordinates given by the minimum start and maximum end positions of the constituent intervals.

Variants were then assigned to a locus if their physical position $p$ fell within the merged locus boundaries.

#### Common variant conditioning

Following the initial gene-based analysis and subsequent meta-analysis, we carried out common variant conditioning for all significant gene-phenotype pairs (Cauchy $P < 2.5 \times 10^{-6}$). For each (biobank, ancestry, phenotype) tuple, we determine a collection of variants to condition on to account for signal driven by common variation nearby. To do this, we created an iterative conditioning pipeline using snakemake. For each MAF mask, we carry out association analysis of all variants with MAF greater than the MAF of the mask within 500 kb of the gene. If any variant association has an association $P$-value $< T$, we add it to a set of conditioning variants and condition on them using the `-condition` flag within SAIGE, and iteratively rerun until no variant in the region is associated ($P$-value $< T$) with the trait. This procedure is carried out for all (ancestry, biobank) pairs. To determine an appropriate $P$-value cutoff ($T$), we performed iterative conditioning with a series of cutoffs across a subset of the biobanks in BRaVa. We selected a cutoff of $P$-value $< 1 \times 10^{-5}$, for (biobank, ancestry) tuples exceeding 100,000 samples, and $P$-value $< 1 \times 10^{-3}$ otherwise (Figs. S41-S43). A more stringent threshold was used in larger cohorts to limit the inclusion of weakly associated variants in the conditioning set, which can otherwise induce collinearity and numerical instability in the conditional test statistic, leading to spuriously inflated associations. Following iterative conditioning within each constituent (biobank, ancestry) tuple, biobanks provide the resultant conditioning variant lists. We then determine the union of these lists centrally for each genetic ancestry. The resultant variant lists are then shared back. Biobanks then perform final gene-based association analysis conditioning on these variants. To guard against collinearity in the variants used for conditioning, we first perform linkage disequilibrium pruning, ensuring that no-pair of variants in the set have $r^2 > 0.9$.

#### Novelty assessment of gene–phenotype associations

We assessed the novelty of gene-phenotype associations using the agentic framework developed in Lu *et al.* (2026). Briefly, the pipeline integrates evidence from two complementary sources: the biomedical literature (via PubMed) and the Open Targets platform. This dual-source strategy mitigates limitations of each resource, as GWAS findings are often under-reported in abstracts, while Open Targets may not capture mechanistic studies, case reports, or animal-model evidence.

Given a gene and phenotype as input, the pipeline independently evaluates evidence from each source and assigns a novelty classification - Not Found, Hypothesized, Existing, or Established. For the literature component, gene aliases (from HGNC) and phenotype synonyms (generated by a language-model–based synonym expansion step) are used to construct both phenotype-specific and agnostic PubMed searches. Retrieved titles and abstracts are then assessed by a novelty-classification agent.

For Open Targets, association scores for the gene across all phenotypes are retrieved and filtered to remove low-confidence signals. Phenotypes matching or closely related to the queried trait are identified using a combination of string matching and model-assisted synonym selection. Novelty labels are assigned based on the maximum association score among matched phenotypes.

Final novelty assignments are obtained by conservatively combining the literature- and Open Targets-based verdicts, taking the most established classification. Consistent with prior observations, Open Targets tended to identify a broader set of associations, while literature-based analysis more often captured mechanistic or contextual evidence not reflected in structured databases.

#### Biobank and cohort acknowledgments

##### All of Us

We gratefully acknowledge All of Us participants for their contributions, without whom this research would not have been possible. We also thank the National Institutes of Health's All of Us Research Program for making available the participant data examined in this study.

##### Biobank Japan

BioBank Japan was supported by the Tailor-Made Medical Treatment program of the Ministry of Education, Culture, Sports, Science, and Technology (MEXT), the Japan Agency for Medical Research and Development (AMED). S.N. was supported by JSPS KAKENHI (26K18279), AMED (JP24tm0424228, JP24tm0524009, JP25kk0305032, and JP256f0137004), Japan Foundation for Applied Enzymology, and The University of Tokyo Pandemic Preparedness, Infection and Advanced Research (UTOPIA) Center. Y.O. was supported by JSPS KAKENHI (25H01057), and AMED (JP24km0405217, JP24ek0109594, JP24ek0410113, JP24kk0305022, JP223fa627001, JP223fa627002, JP223fa627010, JP223fa627011, JP22zf0127008, JP24tm0524002, JP24wm0625504, JP24gm1810011), JST Moonshot R&D (JPMJMS2021, JPMJMS2024), Ono Pharmaceutical Foundation for Oncology, Immunology, and Neurology, Bioinformatics Initiative of Osaka University Graduate School of Medicine, Center for Infectious Disease Education and Research (CiDER), and Center for Advanced Modality and DDS (CAMaD), Osaka University, RIKEN TRIP initiative (AGIS).

#### Bio*Me*

The Mount Sinai Bio*Me* Biobank has been supported by The Andrea and Charles Bronfman Philanthropies and in part by Federal funds from the NHLBI and NHGRI (U01HG00638001; U01HG007417; X01HL134588). We thank all participants in the Mount Sinai Biobank. We also thank all our recruiters who have assisted and continue to assist in data collection and management and are grateful for the computational resources and staff expertise provided by Scientific Computing at the Icahn School of Medicine at Mount Sinai.

#### Colorado Center for Personalized Medicine

We are deeply indebted to the participants who have made The Colorado Center for Personalized Medicine (CCPM) a reality. CCPM would like to thank Richard Zane, Steve Hess, Sarah White, Emily Hearst, Emily Roberts and the entire Health Data Compass team. CCPM is supported through a partnership between UCHealth, the University of Colorado (CU) Anschutz Medical Campus, and the CU School of Medicine. We thank Regeneron Genetics Center for supporting the data generation in this project. CCPM biobank activities were conducted under COMIRB protocol #15-0461.

Colorado Center for Personalized Medicine: Heather D Anderson, Christina L Aquilante, Kelsey Arbogast, Ian M Brooks, Elizabeth E Burke, Emily M Casteel, Joanne B Cole, Curtis R Coughlin II, Jacob Crawford, Kristy Crooks, Erin Culver, Matthew J Fisher, Teresa C Frye, Hunter George, Chris R Gignoux, Elizabeth K Gilliland, Casey S Greene, Emily Hearst, Audrey E Hendricks, Randi K Johnson, Shelby Jones, Dave Kao, Gabrielle A Knortz, Danielle Koffenberger, Santhanagopalan Krishnamoorthy, Lisa Ku, Elizabeth L Kudron, Rashawnda Lacy, Ethan M Lange, Joe A Lesny, Meng Lin, James L Martin, Nicole L McDaniel, Jack Pattee, Nikita Pozdeyev, Alaa Radwan, Nick Rafaels, Sridharan Raghavan, Neda Rasouli, Carolina Sanchez-Wild, Elise L Shalowitz, Hoda Sherif, Johnathan A Shortt, Adrian M Stewart, Carolyn T Swartz, Anna Tanaka, Emily Todd, Katy E Trinkley, Vendant Vohra, and Laura K Wiley.

Please see here for affiliations: `https://docs.google.com/spreadsheets/d/1kDhalCpW2tGPoq6D1SvSmeS0c9jsNPwsiq5R5_xVbeU/edit?usp=sharing`.

#### Estonian Biobank

This research was supported by the European Union through Horizon 2020 research and innovation program under grant no 810645 and through the European Regional Development Fund project no. MOBEC008, by the Estonian Research Council grant PUT (PRG1291, PRG687 and PRG184) and by the European Union through the European Regional Development Fund project no. MOBERA21 (ERA-CVD project DETECT ARRHYTHMIAS, GA no JTC2018-009), Project No. 2014-2020.4.01.15-0012 and Project No. 2014-2020.4.01.16-0125. We would like to acknowledge Dr. Tõnu Esko; Dr. Lili Milani; Dr. Reedik Mägi, Dr. Mari Nelis and Dr. Andres Metspalu, all from the Institute of Genomics, University of Tartu, Tartu, Estonia.

#### Genes & Health

Genes & Health is/has recently been core-funded by Wellcome (WT102627, WT210561), the Medical Research Council (UK) (M009017, MR/X009777/1, MR/X009920/1), Higher Education Funding Council for England Catalyst, Barts Charity (845/1796), Health Data Research UK (for London substantive site), and research delivery support from the NHS National Institute for Health Research Clinical Research Network (North Thames). We acknowledge the support of the National Institute for Health and Care Research Barts Biomedical Research Centre (NIHR203330); a delivery partnership of Barts Health NHS Trust, Queen Mary University of London, St George's University Hospitals NHS Foundation Trust and St George's University of London

Genes & Health is/has recently been funded by Alnylam Pharmaceuticals, Genomics PLC; and a Life Sciences Industry Consortium of AstraZeneca PLC, Bristol-Myers Squibb Company, GlaxoSmithKline Research and Development Limited, Maze Therapeutics Inc, Merck Sharp & Dohme LLC, Novo Nordisk A/S, Pfizer Inc, Takeda Development Centre Americas Inc.

We thank Social Action for Health, Centre of The Cell, members of our Community Advisory Group, and staff who have recruited and collected data from volunteers. We thank the NIHR National Biosample Centre (UK Biocentre), the Social Genetic & Developmental Psychiatry Centre (King's College London), Wellcome Sanger Institute, and Broad Institute for sample processing, genotyping, sequencing and variant annotation. This work uses data provided by patients and collected by the NHS as part of their care and support. This research utilised Queen Mary University of London's Apocrita HPC facility, supported by QMUL Research-IT, `http://doi.org/10.5281/zenodo.438045`

We thank: Barts Health NHS Trust, NHS Clinical Commissioning Groups (City and Hackney, Waltham Forest, Tower Hamlets, Newham, Redbridge, Havering, Barking and Dagenham), East London NHS Foundation Trust, Bradford Teaching Hospitals NHS Foundation Trust, Public Health England (especially David Wyllie), Discovery Data Service/Endeavour Health Charitable Trust (especially David Stables), Voror Health Technologies Ltd (especially Sophie Don), NHS England (for what was NHS Digital) - for GDPR-compliant data sharing backed by individual written informed consent.

Most of all we thank all of the volunteers participating in Genes & Health.

A favourable ethical opinion for the main Genes & Health research study was granted by NRES Committee London - South East (reference 14/LO/1240) on 16 Sept 2014. Queen Mary University of London is the Sponsor, and Data Controller.

Current Genes & Health Research Team: Eamonn Maher Shabana Chaudhary, Joseph Gafton, Karen A Hunt, Shapna Hussain, Kamrul Islam, Mohammed Bodrul Mazid, Elizabeth Owor, Jessry Russell, Nishat Safa, John Solly, Marie Spreckley, David A Van Heel, Jan Whalley,Ishevanhu Zengeya, Emily Mantle, Shaheen Akhtar, Samina Ashraf, Dan Mason, John Wright, Daniel MacArthur, Michael Simpson, Richard C Trembath, Gerome Breen, Raymond Chung, Sang Hyuck Lee, Omar Asgar, Joanne Harvey, Karen Tricker, Caroline Winckley, Hanifa Khatun, Amna Asif, Claudia Langenberg, Grainne Colligan, Ceri Durham, Bill Newman, Ahsan Khan, Hilary Martin, Teng Heng, Matt Hurles, Vivek Iyer, Georgios Kalantzis, Vladimir Ovchinnikov, Iaroslav Popov, Klaudia Walter, Panos Deloukas, David Collier, Ana Angel, Saeed Bidi, Fabiola Eto, Sarah Finer, Chris Griffiths, Sam Hodgson, Benjamin M Jacobs, Rohini Mathur, Caroline Morton, Asma Qureshi, Stuart Rison, Annum Salman, Miriam Samuel, Moneeza K Siddiqui, Daniel Stow, Sabina Yasmin, Julia Zöllner, and Sheik Dowlut.

Please see here for affiliations: `https://docs.google.com/spreadsheets/d/1D9HLbc_m0KdOUN-gS0hymTLewJ36e8ETSEb8Tu_CqWY/edit?gid=0#gid=0`.

#### Genomics England

We gratefully acknowledge the participants of the National Genomic Research Library (NGRL)[22], whose contributions made this research possible. Secure access to the NGRL under project ID RR953 was provided by Genomics England, which delivers the NGRL in partnership with NHS England, and is wholly owned by the UK Department of Health and Social Care. The NGRL contains participants' health data collected by the NHS as part of their care, along with samples and data from their participation in research, for which fully informed consent has been obtained. This includes genomic and clinical data provided through the NHS Genomic Medicine Service, as well as data obtained through research studies, including the 100,000 Genomes Project and the Generation Study, both of which are delivered in partnership with the NHS, and from other research cohorts involving external collaborators.

#### Mass General Brigham Biobank

Biobank samples, genomic data, and health information were obtained from the Mass General Brigham Biobank, a bio-repository of consented patients samples at Mass General Brigham (parent organization of Massachusetts General Hospital and Brigham and Women's Hospital). We are grateful to all of the participants and clinical and research teams who made this work possible. Support for genotyping was provided through MGB Personalized

Medicine.

MGB Biobank Leadership: Elizabeth W. Karlson, MD; Shawn N. Murphy, MD, PhD; Susan A Slaugenhaupt, PhD; Jordan W. Smoller, MD, ScD; Scott T. Weiss, MD, MSc.

#### Penn Medicine BioBank

We acknowledge the Penn Medicine BioBank (PMBB) for providing data and thank the patient participants of Penn Medicine who consented to participate in this research program. We would also like to thank the Penn Medicine BioBank team and Regeneron Genetics Center for providing genetic variant data for analysis. The PMBB is approved under IRB protocol #813913 and supported by Perelman School of Medicine at University of Pennsylvania, a gift from the Smilow family, and the National Center for Advancing Translational Sciences of the National Institutes of Health under CTSA award number UL1TR001878.

**PMBB Leadership Team** Daniel J. Rader, Marylyn D. Ritchie, Michael D. Feldman. *Contributions:* All authors contributed to securing funding, study design and oversight.

**Patient Recruitment and Regulatory Oversight** JoEllen Weaver, Afiya Poindexter, Ashlei Brock, Khadijah Hu-Sain, Yi-An Ko. *Contributions:* JW manage patient recruitment and regulatory oversight of study. AP, AB, KH, YK recruitment and enrollment of study participants.

**Lab Operations** JoEllen Weaver, Meghan Livingstone, Fred Vadivieso, Ashley Kloter, Stephanie DerOhannessian, Teo Tran, Linda Morrel, Ned Haubein, Joseph Dunn. *Contributions:* JW, ML, FV, SD oversight of lab operations. ML, FV, AK, SD, TT, LM perform sample processing. NH, JD are responsible for sample tracking and the laboratory information management system.

**Clinical Informatics** Anurag Verma, Ph.D., Colleen Morse, M.S., Marjorie Risman, M.S., Renae Judy, B.S. Contributions: All authors contributed to the development and validation of clinical phenotypes used to identify study subjects and (when applicable) controls.

**Genome Informatics** Anurag Verma Ph.D., Shefali S. Verma, Ph.D., Yuki Bradford, M.S., Scott Dudek, M.S., Theodore Drivas, M.D., Ph.D. *Contributions:* A.V., S.S.V. are responsible for the analysis design and infrastructure needed for quality control of genotype and exome data. Y.B. performed the analysis. T.D. and A.V. provide variant and gene annotations and their functional interpretation of variants.

We thank PMBB collaborators at Penn for their cooperation in these studies including Katherine Nathanson (Uterine and Thyroid cancer), Peter Merkel (Gout), Joan O'Brien (Primary open-angle glaucoma), Sharlene Day

(Cardimyopathy), Tom Cappola, Julio Chirinos, Nosheen Reza (Heart Failure), Dr. Brett Cucchiara (Stroke), Scott Damrauer (Abdominal aortic aneurysm), Blanca Himes (Asthma, COPD), Lilly McGuire (Acute appendicitis).

##### UK Biobank

Access to data from the UK Biobank was obtained through Application #31063 PIs: Ben Neale, Claire Churchhouse: ‘Methodological extensions to estimate genetic heritability and shared risk factors for phenotypes of the UK Biobank’, and Application #11867 PIs: Cecilia Lindgren, Duncan Palmer: ‘Dissection of the Genetic Susceptibility of Obesity Traits and their Comorbidities’.

##### ICDA

The authors would like to acknowledge the organizing committee of the International Common Disease Alliance (ICDA) for intellectual contributions on the set up of the BRaVa consortium as a contributing activity to the larger effort. We also thank them for the use of their slack platform. We would especially like to thank Sam Bryant from the Stanley Center Data Management team for helping with the Google bucket set up and data sharing.

#### Harmonization of regenie and SAIGE gene-burden effect sizes

A challenge in combining gene-level results from SAIGE and regenie summary statistic output in an inverse-variance weighted meta-analysis is that the two packages use different default variant weighting schemes and aggregation methods at the gene-level, resulting in burden effect sizes ($\beta_{\text{Burden}}$) and standard errors ($SE_{\text{Burden}}$) on different scales. Specifically, while variant-level effect sizes show high concordance between the two methods, SAIGE computes the burden statistic using variant weights derived from a Beta distribution function of the minor allele frequency ($Beta(\text{MAF}; 1, 25)$) by default, whereas regenie defaults to a uniform weighting ($w_i = 1$) independent of MAF.

To place gene-burden effect sizes on the same scale prior to inverse variance weighted meta-analysis, we can reconstruct SAIGE-equivalent gene-burden effect sizes and standard errors using the variant-level summary statistics output by regenie.

For each variant $i$ within a given gene and mask, the Score statistic ($T_i$) and its variance can be determined from the regenie variant-level outputs:

$$T_i = \frac{\chi_i^2}{\beta_i} = \frac{\beta_i}{SE_i^2}$$

$$\text{Var}\,(T_i) = \frac{T_i^2}{\chi_i^2} = \frac{1}{SE_i^2}$$

where $\beta_i$ is the marginal effect size of variant $i$, and $\chi_i^2$ is its associated $\chi_1^2$ statistic.

To mirror the SAIGE burden aggregation, we can assign each variant a weight $w_i$ based on its cohort-specific MAF, evaluating the probability density function of the Beta distribution: $w_i = Beta(\text{MAF}_i; 1, 25)$. The gene-burden effect size on the SAIGE scale and its corresponding standard error can then be constructed by aggregating the weighted variant-level score statistics and variances across all variants $i \ldots n$ in the gene mask:

$$\beta_{\text{Burden}} = \frac{\sum_{i=1}^{n} w_i T_i}{\sum_{i=1}^{n} w_i^2 \text{Var}(T_i)}$$
$$SE_{\text{Burden}} = \frac{1}{\sqrt{\sum_{i=1}^{n} w_i^2 \text{Var}(T_i)}}.$$

This transformation ensures that regenie-derived summary statistics are placed on a consistent scale with SAIGE outputs, enabling valid inverse-variance weighted meta-analysis at the gene-level across cohorts.

#### Spermatagonia gene-set enrichment testing

To assess whether genes under positive selection in spermatogonia (CES genes) are enriched among BRaVa gene-level associations, we performed logistic regression analyses adjusting for gene-level covariates reflecting mutational target size, evolutionary constraint, and functional essentiality.

##### Definition of gene sets and association status

Gene sets corresponding to CES categories (gain-of-function, LoF-1, LoF-2, previously reported CES genes, and Neville *et al.* gene set) were curated from published sources and encoded as binary indicators. Gene-level association results were obtained from BRaVa meta-analyses across quantitative and binary traits. For each gene, we defined a binary outcome variable indicating whether the gene exhibited at least one experiment-wise significant association in any trait.

##### Gene-level covariates

To ensure consistent gene definitions across analyses, we restricted all gene-level covariates to MANE Select transcripts[23] obtained from Ensembl[24] (GRCh38). For each gene, we defined gene length as the total number of coding bases across all exons of the MANE transcript.

Evolutionary constraint was quantified using phyloP scores derived from the 100-way vertebrate alignment[25]. PhyloP scores were extracted from the hg38 bigWig track and averaged across exonic bases of each MANE transcript. To avoid excessive memory usage, phyloP scores were computed in chunks and aggregated using a length-weighted mean across overlapping intervals.

Functional essentiality was measured using CRISPR-based gene effect scores from the DepMap project[26]. For each gene, we computed the median gene effect score across all available cell lines.

#### Statistical analysis

We evaluated enrichment of CES gene sets among BRaVa-associated genes using logistic regression models of the form:

$$\text{logit}\left(P(\text{hit})\right) = \beta_0 + \beta_1 \cdot \text{GeneSet} + \beta_2 \cdot \log_{10}(\text{gene length}) + \beta_3 \cdot \text{phyloP} + \beta_4 \cdot \text{DepMap score}.$$

Separate models were fitted for each CES gene set. Regression coefficients for gene set membership were exponentiated to obtain odds ratios, with corresponding 95% confidence intervals derived from standard errors.

#### Rationale for covariate selection

We avoided reliance on constraint metrics derived from human population variation (e.g., LOEUF[14]), as such measures may be influenced by mutation model misspecification and exhibit systematic biases in genes under positive selection in spermatogonia. Instead, we incorporated orthogonal measures of evolutionary conservation (phyloP) and experimental essentiality (DepMap) to reduce potential circularity and confounding in gene-level enrichment analyses.

#### Supplementary Figures

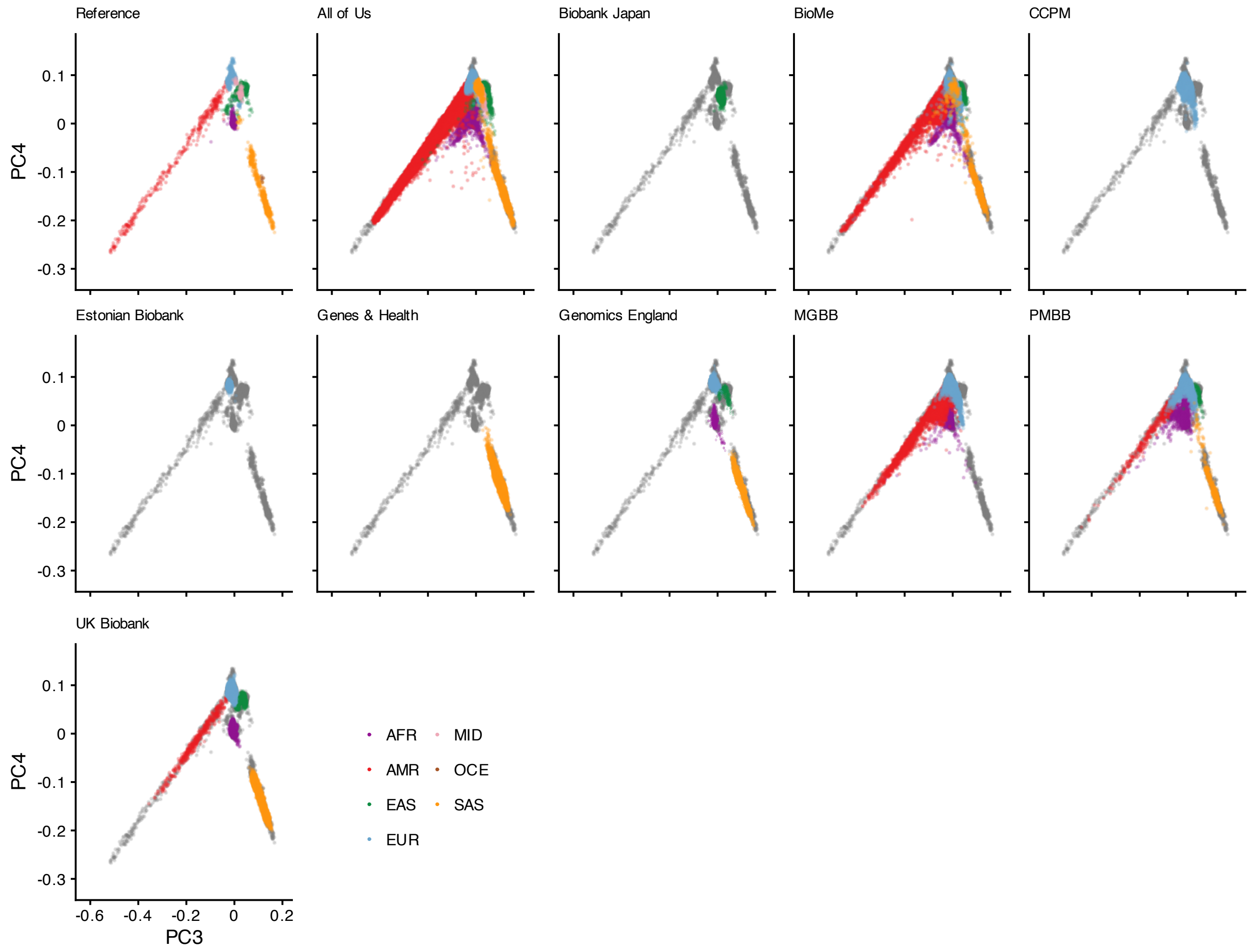

**Figure S1: Projection of contributed samples from studies participating within the BRaVa consortium into the PC space defined by 1000G and HGDP: PCs 3-4.** Using the PC space defined by 1000G and HGDP as a reference, analysts projected samples from contributing studies into the space using a common set of ∼165,000 variants. In each panel (except for the reference), colored points correspond to contributed samples from each cohort. Grey points on each panel (except the reference) denote the reference samples (which are colored in the first panel). Points are colored according to the genetic ancestry label determined by each contributing biobank. As GEL was only able to submit images of the projection of samples into PC space, we overlaid submitted transparent images onto reference samples, rather than plotting them. Here we show projections onto PCs 3 and 4.

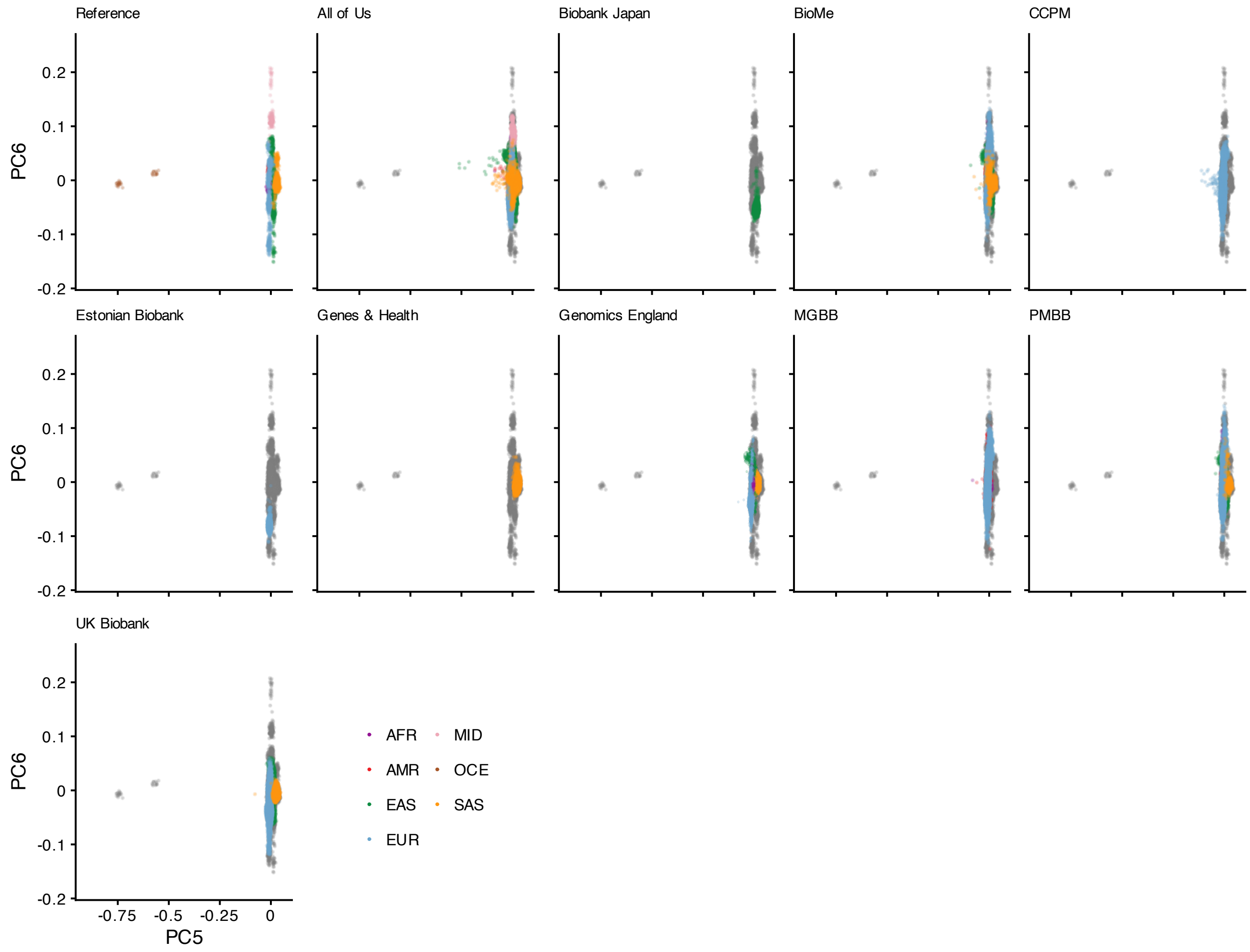

**Figure S2: Projection of contributed samples from studies participating within the BRaVa consortium into the PC space defined by 1000G and HGDP: PCs 5-6.** Using the PC space defined by 1000G and HGDP as a reference, analysts projected samples from contributing studies into the space using a common set of ~165,000 variants. In each panel (except for the reference), colored points correspond to contributed samples from each cohort. Grey points on each panel (except the reference) denote the reference samples (which are colored in the first panel). Points are colored according to the genetic ancestry label determined by each contributing biobank. As GEL was only able to submit images of the projection of samples into PC space, we overlaid submitted transparent images onto reference samples, rather than plotting them. Here we show projections onto PCs 5 and 6.

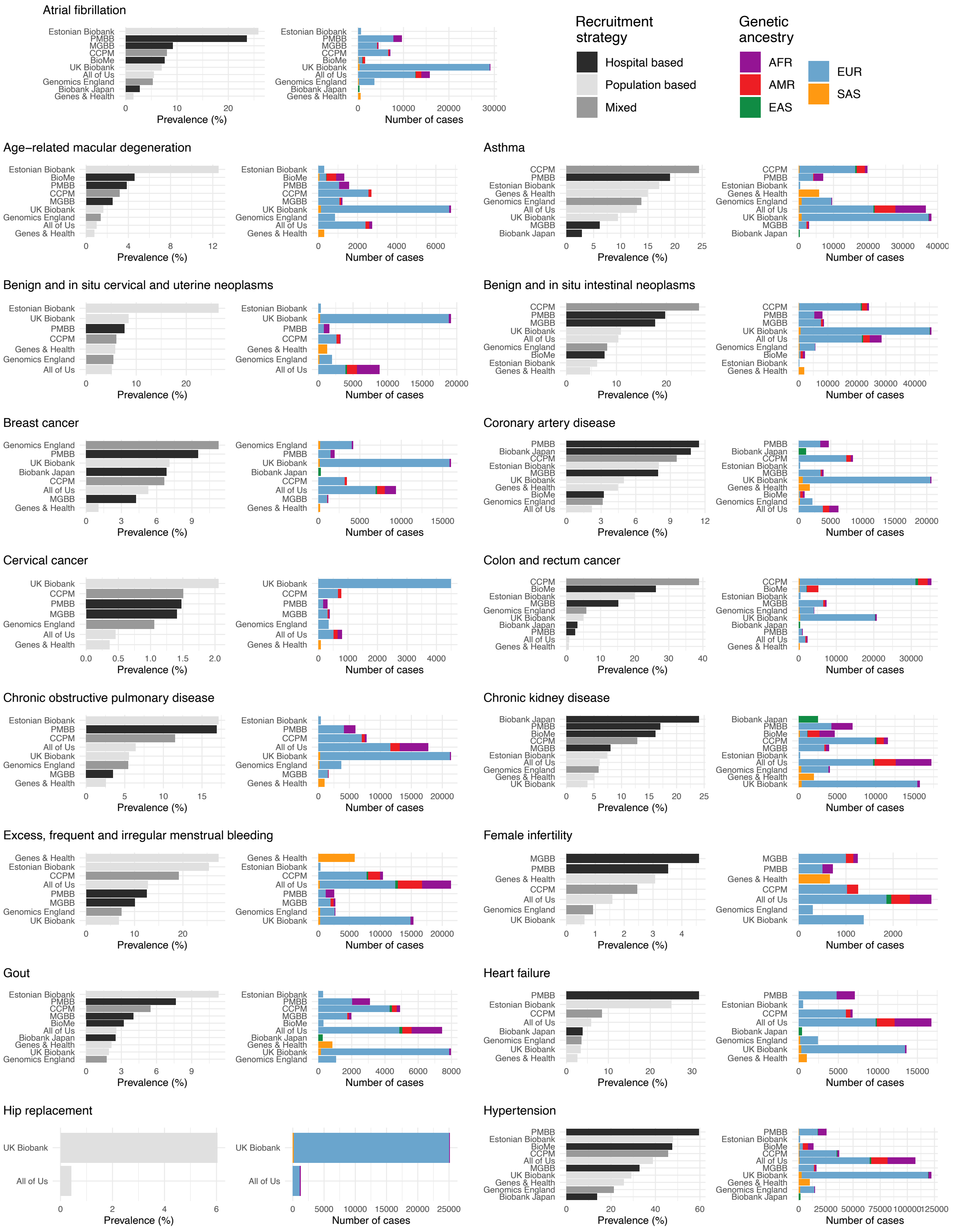
Atrial fibrillation
Recruitment strategy
Hospital based
Population based
Mixed
Genetic ancestry
AFR
AMR
EAS
EUR
SAS
Age−related macular degeneration
Asthma
Benign and in situ cervical and uterine neoplasms
Benign and in situ intestinal neoplasms
Breast cancer
Coronary artery disease
Cervical cancer
Colon and rectum cancer
Chronic obstructive pulmonary disease
Chronic kidney disease
Excess, frequent and irregular menstrual bleeding
Female infertility
Gout
Heart failure
Hip replacement
Hypertension
Prevalence (%)
Number of cases

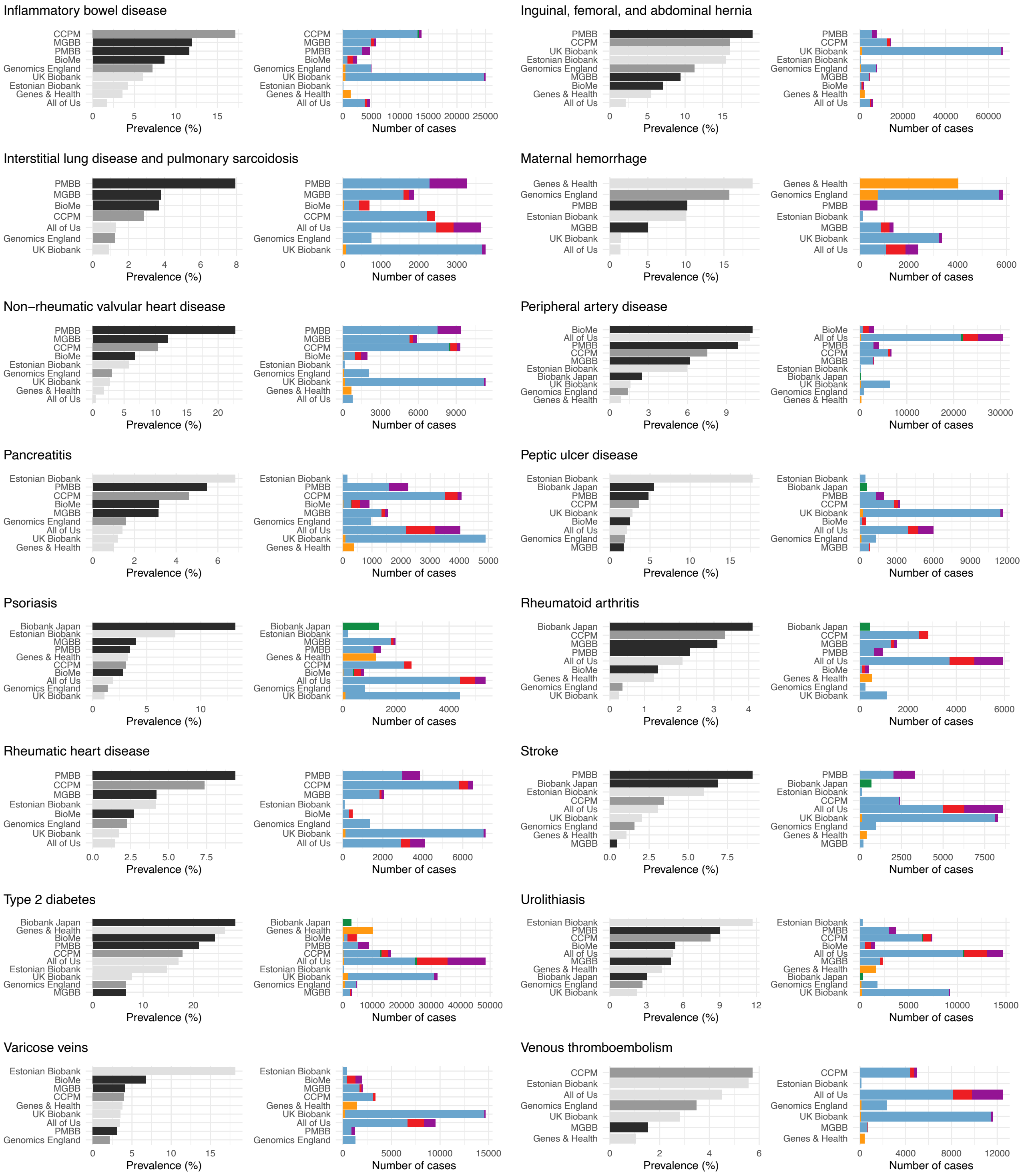

**Figure S3: Disease prevalence differs by biobank and recruitment strategy.** Sample prevalences for each biobank are displayed for each of the analyzed binary traits (left bar plots), together with case counts (right bar plots). For each phenotype, rows are ordered by prevalence, from most to least common. The right bar plots are colored by recruitment strategy according to the legend. The right stacked bar plots are stacked based on broad ancestry labels, detailed in the legend.

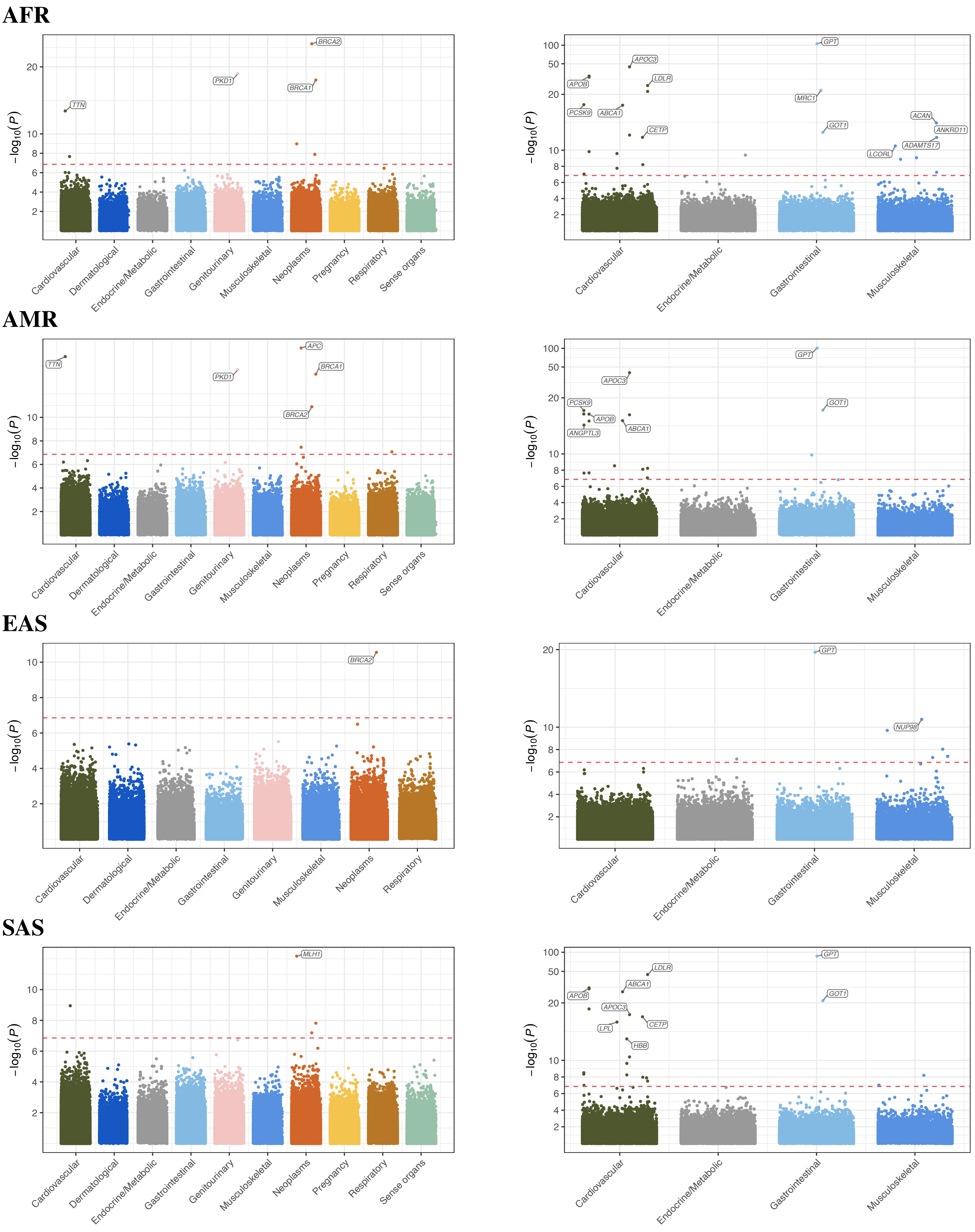

**Figure S4: AFR, AMR, EAS, SAS meta-analysis results.** Summary of gene-based analysis results split by ICD chapter, for continuous and binary traits within non-European broad continental ancestries. Throughout, we display the most significant association among all (MAF cutoff, annotation) pairs across test type (burden, SKAT, SKAT-O), where MAF cutoff $\in \{0.1\%, 0.01\%\}$, annotation $\in$ {pLoF, damaging missense or protein altering, pLoF or damaging missense or protein altering}. The red dotted horizontal lines denote the experiment-wise significance thresholds for gene-masks ($P = 1.4 \times 10^{-7}$).

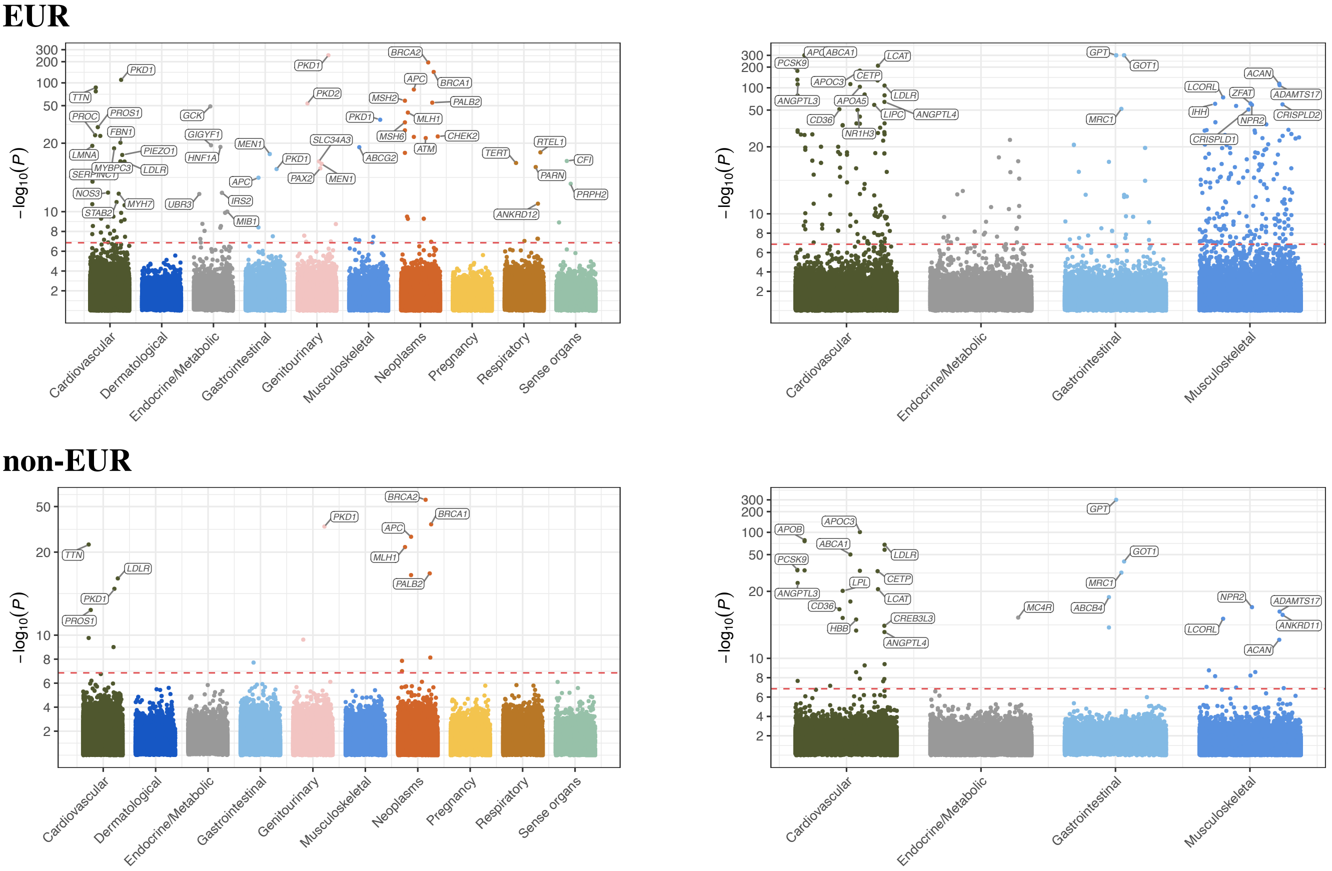

**Figure S5: EUR and non-EUR meta-analysis results.** Summary of gene-based analysis results split by ICD chapter, for continuous and binary traits, restricting meta-analysis to EUR and non-EUR subsets of the data, respectively. Throughout, we display the most significant association among all (MAF cutoff, annotation) pairs across test type (burden, SKAT, SKAT-O), where MAF cutoff $\in$ {0.1%, 0.01%}, annotation $\in$ {pLoF, damaging missense or protein altering, pLoF or damaging missense or protein altering}. The red dotted horizontal lines denote the experiment-wise significance thresholds for gene-masks ($P = 1.4 \times 10^{-7}$).

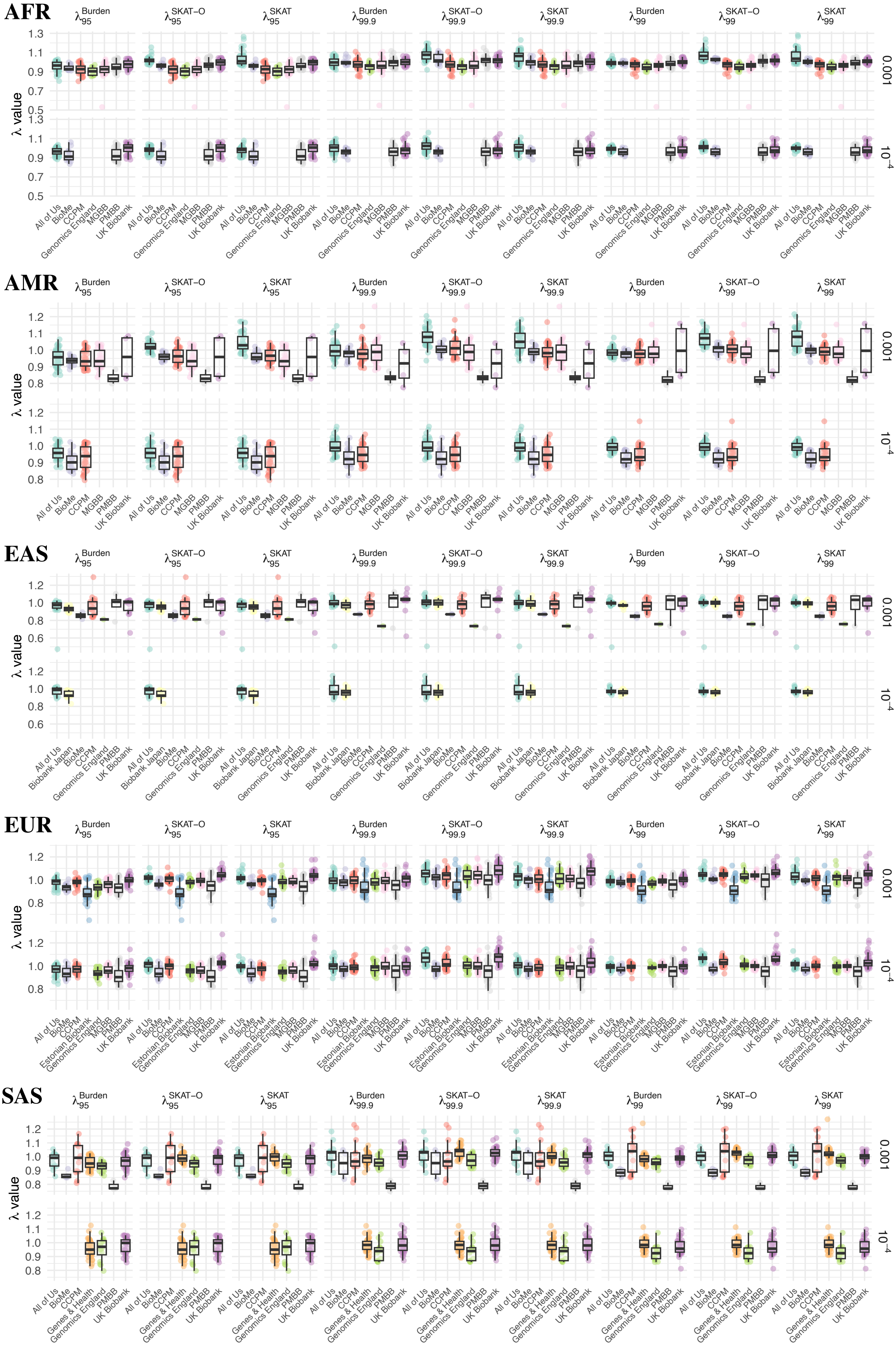
AFR
AMR
EAS
EUR
SAS
λ95 Burden
λ95 SKAT−O
λ95 SKAT
λ99.9 Burden
λ99.9 SKAT−O
λ99.9 SKAT
λ99 Burden
λ99 SKAT−O
λ99 SKAT
λ value
0.001
10−4
All of Us
BioMe
Biobank Japan
CCPM
Estonian Biobank
Genes & Health
Genomics England
MGBB
PMBB
UK Biobank

**Figure S6: Summary of inflation estimates across gene-based associations test statistics, split by (ancestry, biobank) pairs.** For each broad genetic ancestry, we show the genomic control factor ($\lambda$) at the 95th, 99th, and 99.9th percentile for gene-based tests of rare synonymous variation. We consider these upper percentiles, as many of the analyzed traits have relatively low case counts will lead to apparent deflation in test statistics if the median ($\lambda_{GC}$) is used. Facets split according the association tests carried out (burden, SKAT, SKAT-O). Each dot is a (phenotype, ancestry) pair. The first and second rows for each ancestry display the genomic inflation factors for 0.1% and 0.01% MAF masks. Box and whisker plots (box = interquartile range (IQR: [Q1, Q3]), middle = median, whiskers denote the range, except for outliers: defined as points outside [Q1 - 1.5 × IQR, Q3 + 1.5 × IQR]) are overlaid. Dots are jittered and colored by biobank to guide the eye.

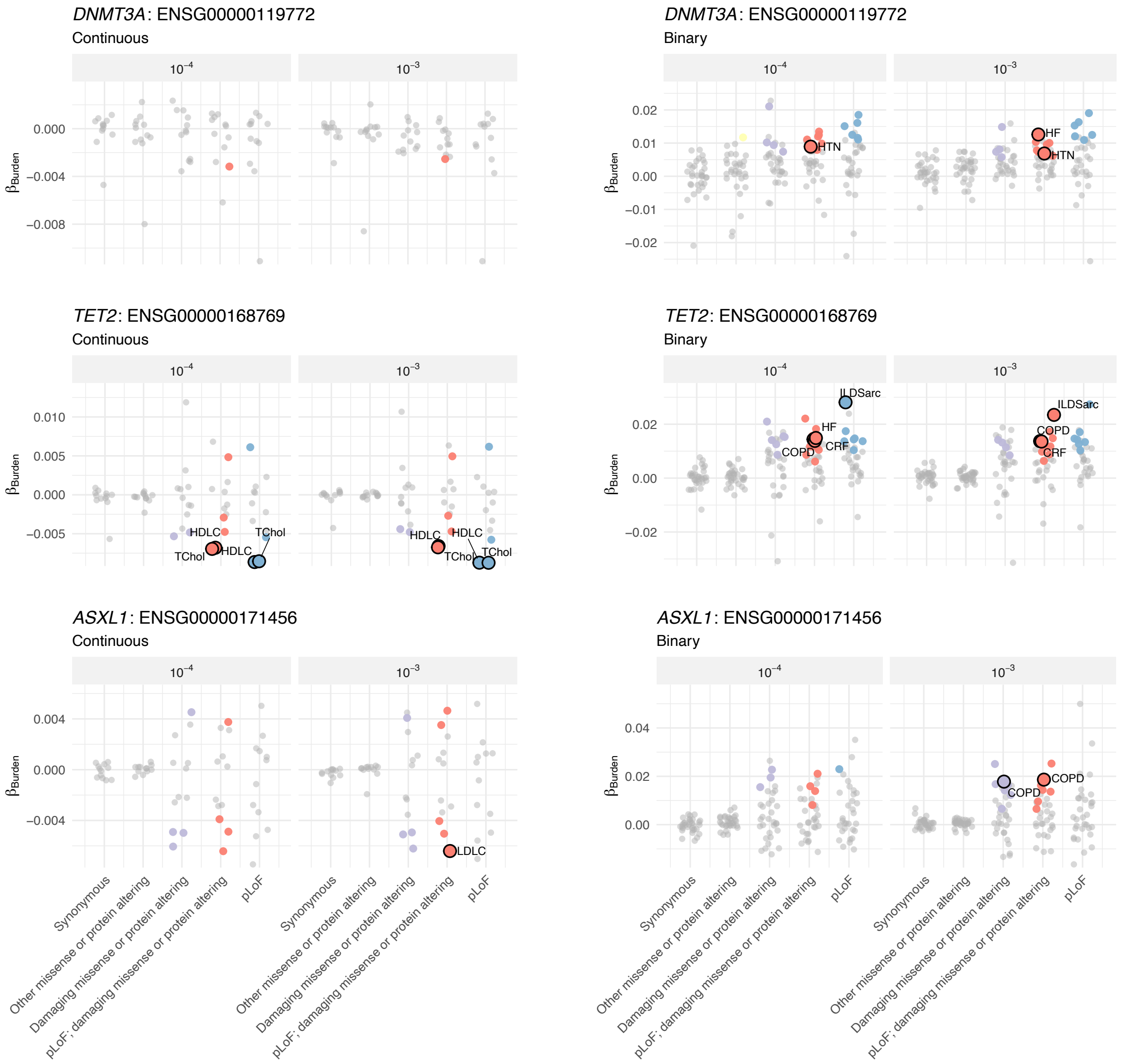

**Figure S7: Gene–trait associations across variant classes for *DNMT3A*, *TET2*, and *ASXL1*.** Scatter plots showing estimated effect burden effect sizes ($\beta_{\text{Burden}}$) from meta-analyzed gene-burden tests for three clonal hematopoiesis–associated genes (*DNMT3A*, *TET2*, and *ASXL1*) across variant annotation classes. Results are presented for continuous (left column) and binary (right column) traits, with two maximum MAF cutoffs (0.1% and 0.01%). Each point represents a trait-specific association, with larger outlined points indicating experiment-wise significant ($P < 1.39 \times 10^{-7}$) associations. Associations with $1.39 \times 10^{-7} < P < 1 \times 10^{-3}$ are colored but not outlined. Associations with $P > 1 \times 10^{-3}$ are displayed in gray.

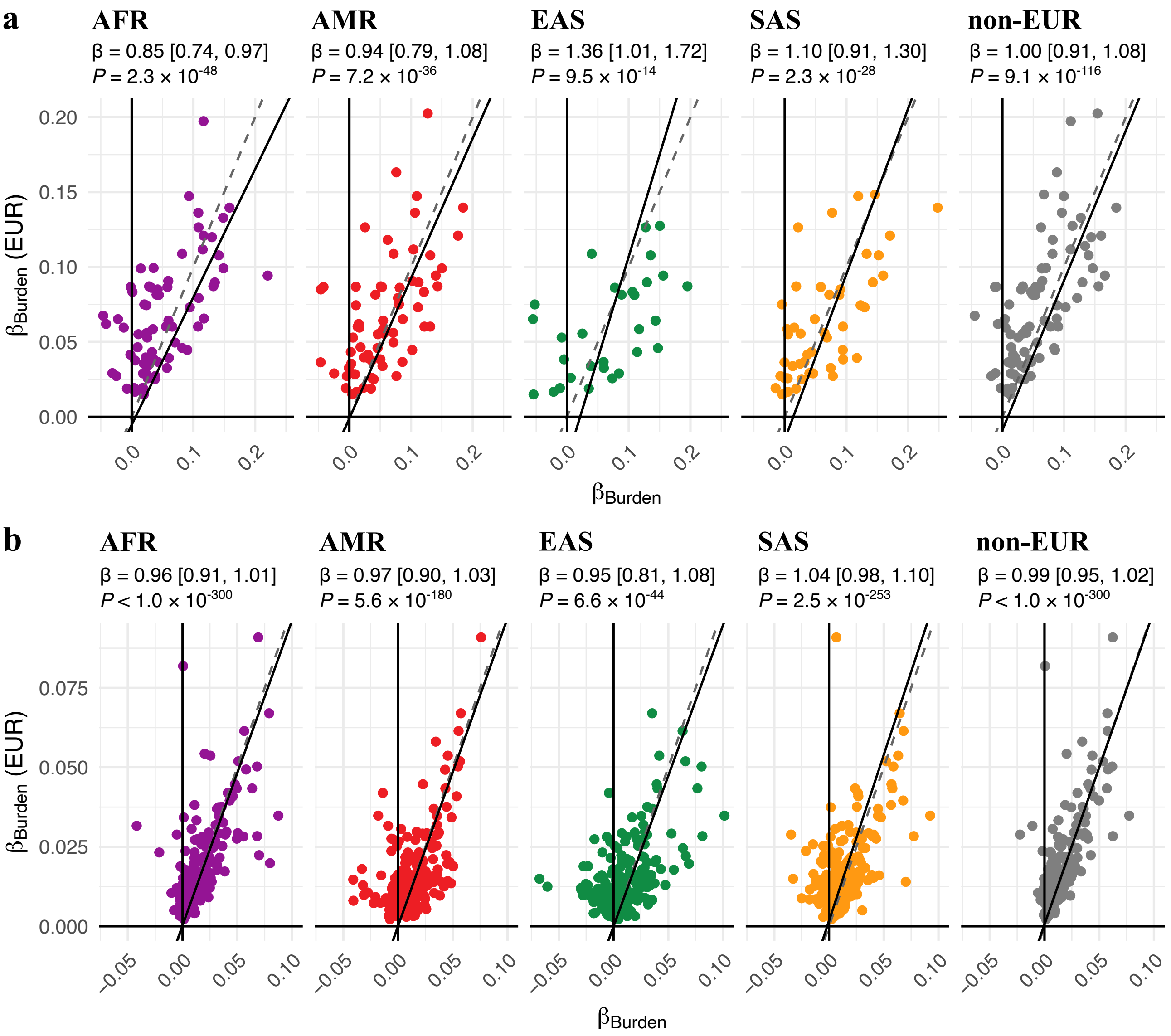

**Figure S8: Consistent effect size estimates across genetic ancestry labels.** Gene-level pLoF (MAF < 0.1%) burden effect sizes from European-ancestry meta-analyses using inverse-variance weighting are shown on the $y$-axis, with corresponding effect size estimates for other broad continental ancestries on the $x$-axes. The rightmost panel shows EUR against all non-European test statistics meta-analyzed together. In each panel, we subset to associations in EUR meta-analysis with $P < 0.05/(20,000 \times 3 \times 2) = 4.2 \times 10^{-7}$, and further subset to association test statistics with a standard error < 0.06 the other meta-analysis, plotting the resultant pairs. a and b display binary and continuous traits, respectively. The black solid lines show $y = x$, and the dashed gray lines show results from error-in-variable total-least-squares Deming regression. At the top of each panel we show results from Deming regression tests: estimated $\beta$ [95% CI] and associated $P$-value for the slope.

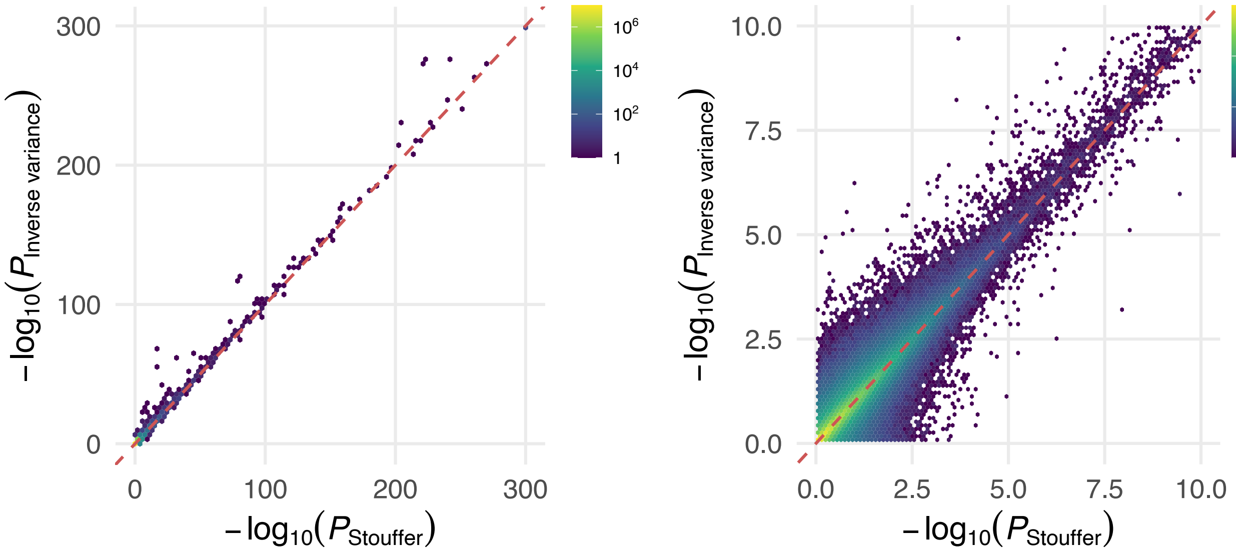

**Figure S9: Comparison of Stouffer and inverse-variance weighted meta-analysis $P$-values at the gene-level.** $-\log_{10}(P)$ **values evaluated Stouffer's method and inverse-variance weighting applied to burden associated test statistics at the gene-level**. Points are binned and colored according to the count of points within each bin according to the legend. The right plot zooms into the $[0, 10] \times [0, 10]$ region of the plot on the left.

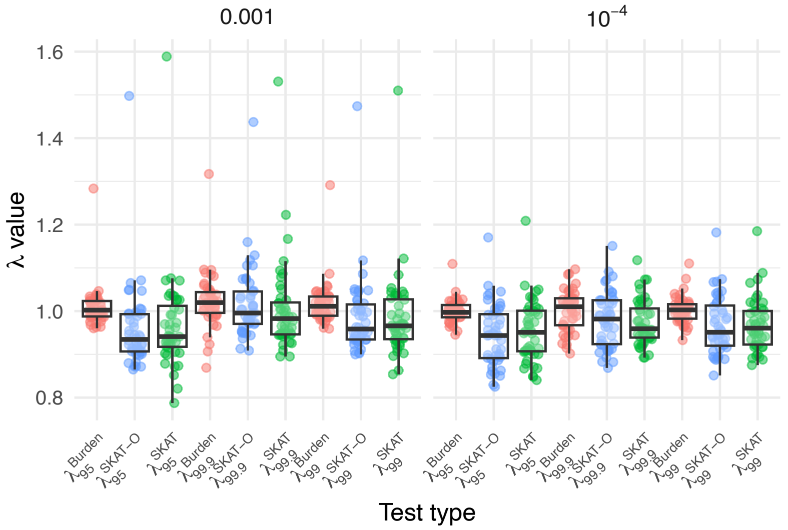

**Figure S10: Summary of inflation estimates across gene-based association test statistics following meta-analysis across (biobank, ancestry) tuples.** Following meta-analysis across all (genetic ancestry, biobank) tuples, we show the resultant genomic control factor ($\lambda$) at the 95th, 99th, and 99.9th percentile for meta-analyzed rare synonymous gene-based tests. Facets split according the association tests carried out (burden, SKAT, SKAT-O). Each dot is a (phenotype) pair. The first and second facets display genomic inflation factors for 0.1% and 0.01% MAF masks. Box and whisker plots (box = interquartile range (IQR: [Q1, Q3]), middle = median, whiskers denote the range, except for outliers: defined as points outside [Q1 - $1.5 \times$ IQR, Q3 + $1.5 \times$ IQR]) are overlaid. Dots are jittered and colored by test used (burden, SKAT, SKAT-O) to guide the eye.

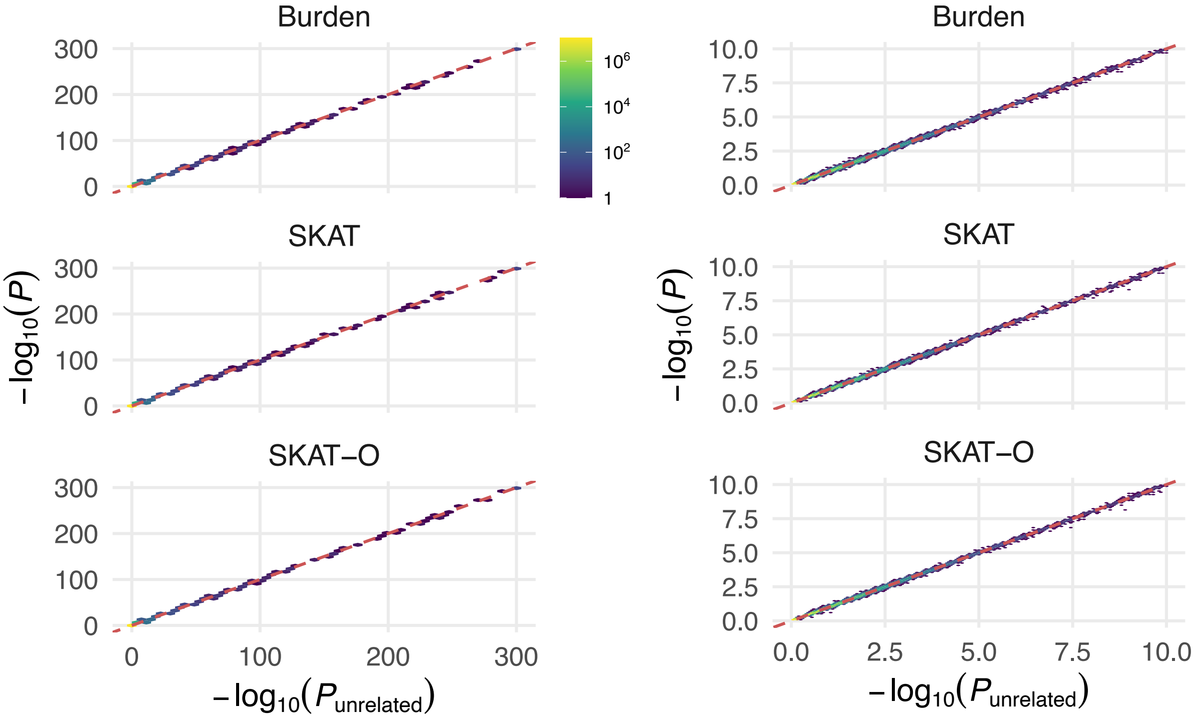

**Figure S11: Comparison of $P$-values when assuming unrelated samples to those estimated using the genetic relatedness matrix in each (biobank, ancestry) subset.** $-\log_{10}(P)$ values are plotted against each other following meta-analysis using Stouffer's method (methods). Points are binned and colored according to the count of points within each bin according to the legend. We plot each class of test (burden, variance-based (SKAT), and hybrid (SKAT-O)) on each row. The column of plots on the right zooms into the $[0, 10] \times [0, 10]$ region of the corresponding plots on the left.

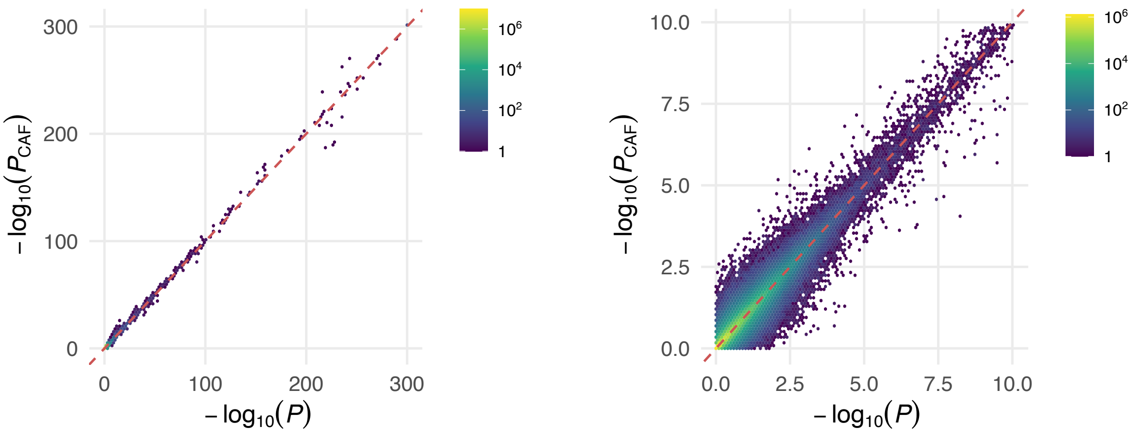

**Figure S12: Comparison of Stouffer based meta-analysis $P$-values at the gene-level with and without weighting by CAF.** $-\log_{10}(P)$ values evaluated Stouffer's method weighting by $\sqrt{N_{\text{eff}}}$ and $\sim \sqrt{N_{\text{eff}}\ 2\ \text{CAF}_{\text{mask}}(1 - \text{CAF}_{\text{mask}})}$ on the $x$ and $y$ axis respectively. Here, $\text{CAF}_{\text{mask}}$ is the cumulative allele frequency of variants within the analyzed mask, in that (biobank, genetic ancestry) sub-cohort. Points are binned and colored according to the count of points within each bin according to the legend. The right plot zooms into the $[0, 10] \times [0, 10]$ region of the plot on the left.

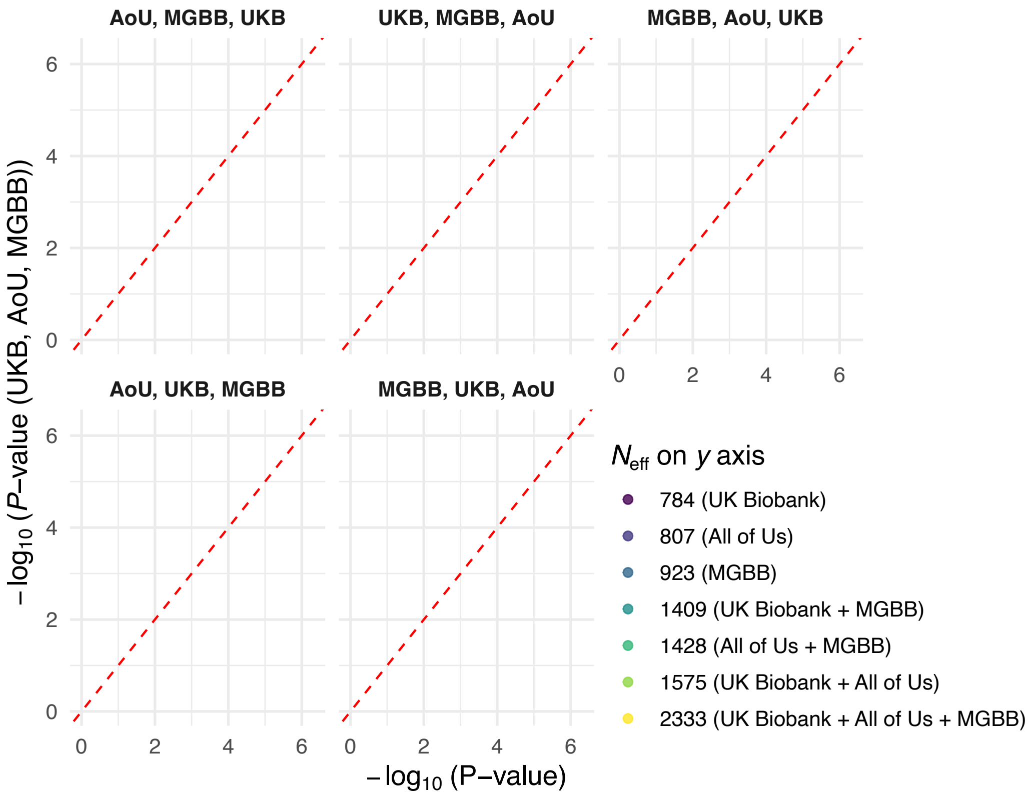

**Figure S13: Comparison of Stouffer based meta-analysis results when accounting overlap in METAL.** Here we compare resultant gene-level association $P$-values as a function of summary statistic input order for an example trait (abdominal aortic aneurysm) with moderate prevalence in three biobanks ($N_{\mathrm{eff}}$ is displayed in the legend), taken from biobank specific summary statistic data[27]. Facet titles display input order on the $x$-axis, compared to (UKB, AoU, MGBB) on the $y$-axis. We color by effective sample size, which demonstrates the source of the discrepancy. METAL evaluates correlations to evaluate meta-analyzed $Z$, sequentially adding studies. As a result, differential missingness leads to distinct collections of being included in evaluation of the sample correlation as a function of input order.

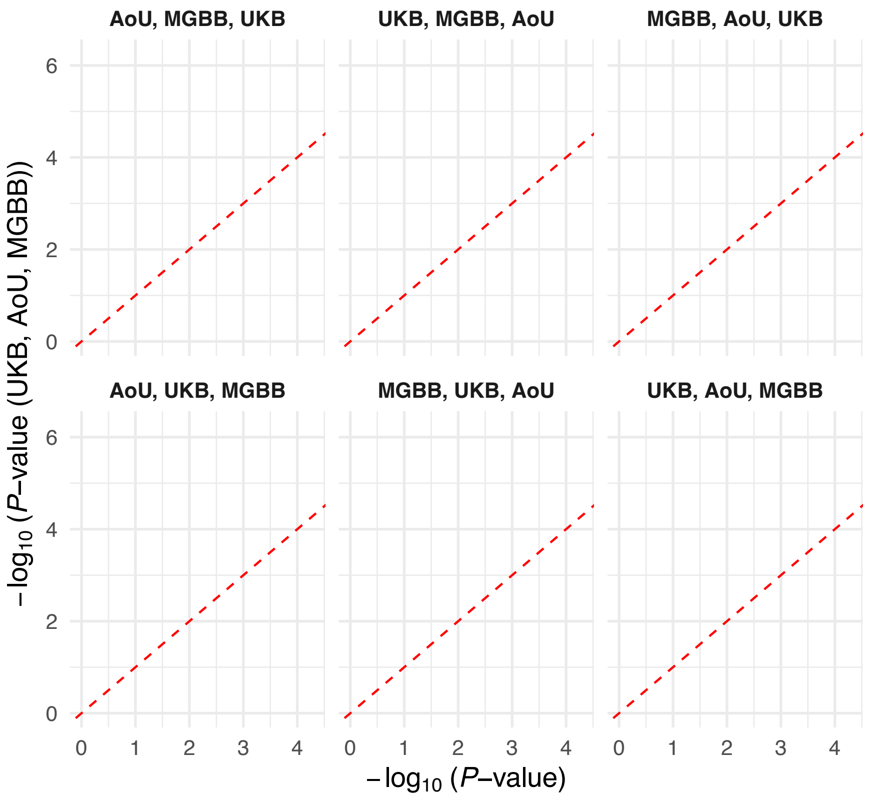

**Figure S14: Comparison of Stouffer based meta-analysis results when accounting overlap in METAL.** Here we compare resultant gene-level association $P$-values as a function of summary statistic input order for an example trait (acquired hemolytic anemias) with moderate prevalence in three biobanks ($N_{\text{eff}}$ is displayed in the legend) using biobank specific summary statistic data[27] as input. Facet titles display input order on the $x$-axis, compared to (UKB, AoU, MGBB) on the $y$-axis. We color by effective sample size, which demonstrates the source of the discrepancy. METAL evaluates correlations to evaluate meta-analyzed $Z$, sequentially adding studies. As a result, differential missingness leads to distinct collections of being included in evaluation of the sample correlation as a function of input order.

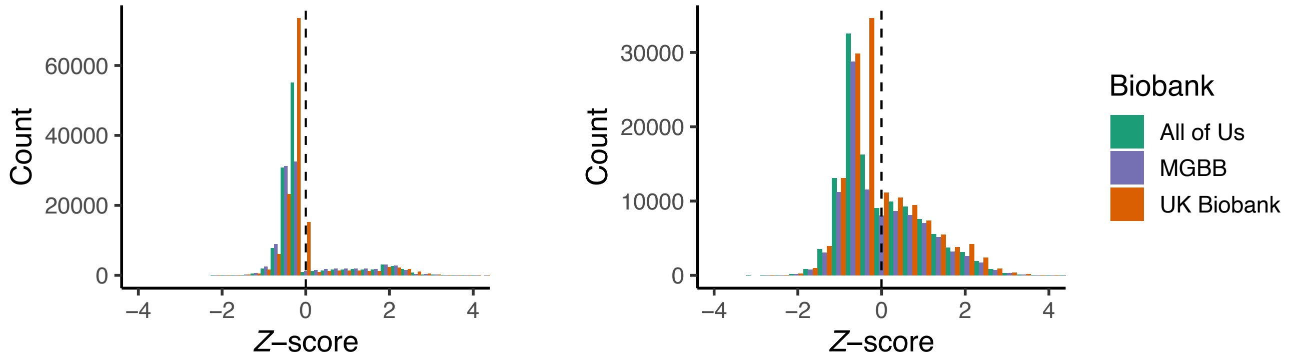

**Figure S15: Empirical $Z$-score statistic distribution among gene-based association tests in three biobanks for binary traits**. Here we display histograms of $Z$-scores taken from acquired hemolytic anemias (left) and abdominal aortic aneurysm (right), as exemplar traits with medium and low prevalence in European populations. Distributions of $Z$-scores for AoU, MGBB and UKB are displayed according to the legend. METAL's approach to account for sample overlap using summary level data relies on the assumption that null results follow a truncated normal distribution. This assumption breaks down for rare variation, and is exacerbated as disease prevalence reduces.

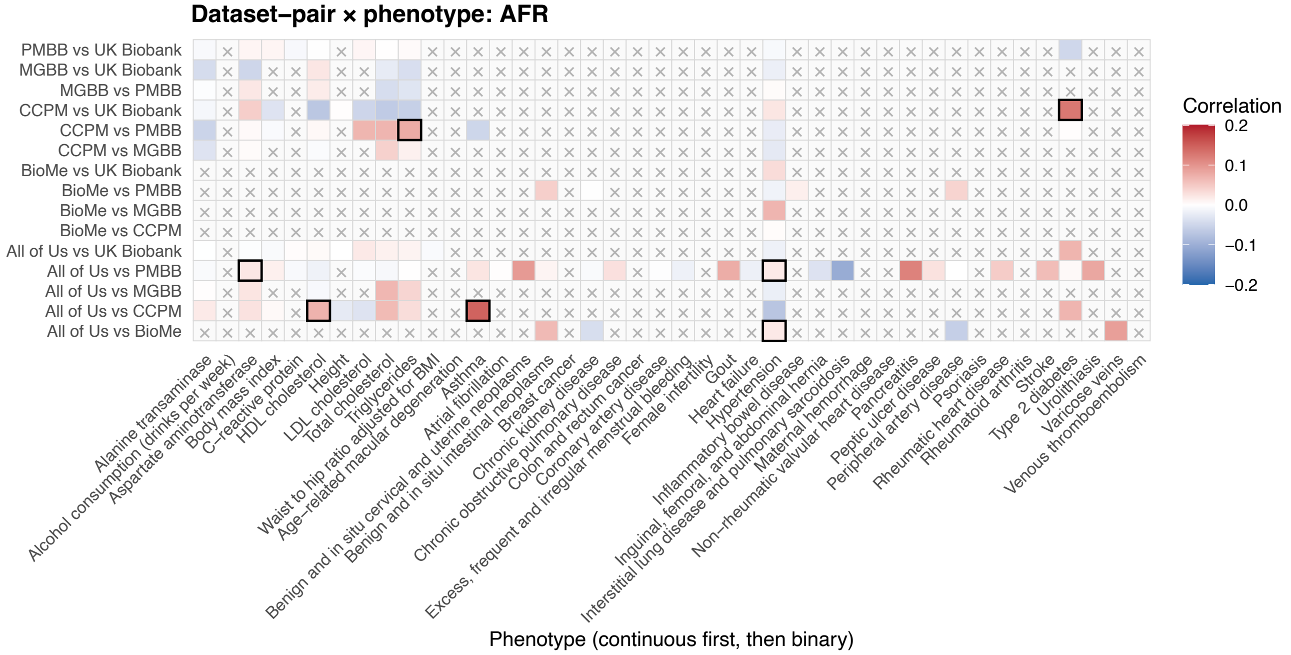

**Figure S16: Correlation between synonymous gene-level associations under the MAF $< 0.1\%$ mask in AFR.** For each trait, we compared gene-level synonymous burden test $Z$-scores (MAF $< 0.1\%$) between biobanks. Genes were excluded from correlation analyses if the cumulative minor allele count was $< 50$ for continuous traits or $< 30$ among cases for binary traits. Correlations were not computed when fewer than 100 genes met inclusion criteria, or when the phenotype was unavailable for a given (biobank, ancestry) pair; such comparisons are indicated by an $\times$. One-sided $P$-values were calculated to test for positive correlation, and nominally significant correlations ($P < 0.05$) are outlined in black. Correlation coefficients are colored according to the legend. Traits are grouped by continuous and binary phenotypes and ordered alphabetically along the $x$-axis. Biobank comparisons are ordered alphabetically from bottom to top along the $y$-axis.

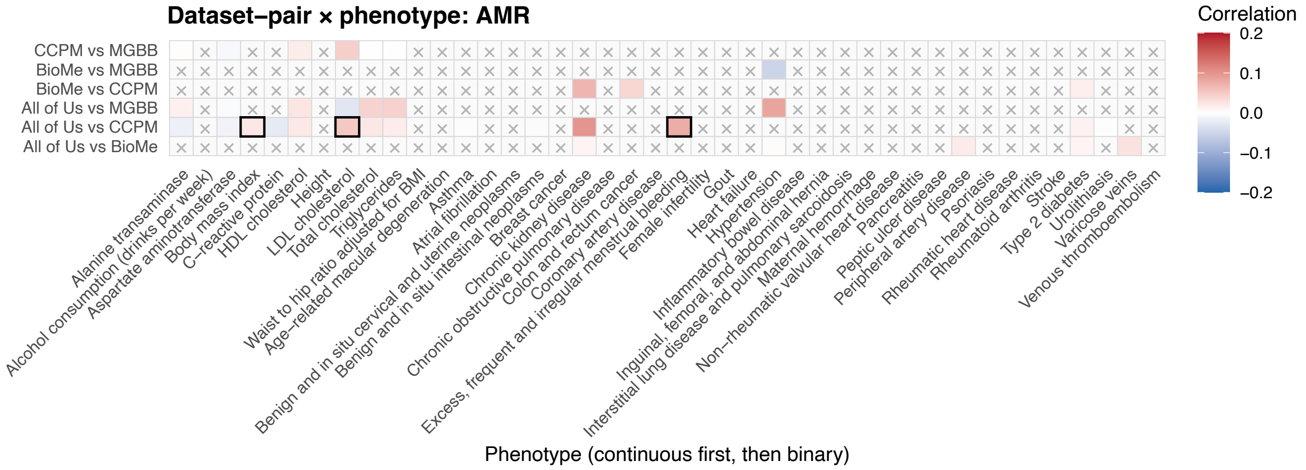

**Figure S17: Correlation between synonymous gene-level associations under the MAF $< 0.1\%$ mask in AMR.** For each trait, we compared gene-level synonymous burden test $Z$-scores (MAF $< 0.1\%$) between biobanks. Genes were excluded from correlation analyses if the cumulative minor allele count was $< 50$ for continuous traits or $< 30$ among cases for binary traits. Correlations were not computed when fewer than 100 genes met inclusion criteria, or when the phenotype was unavailable for a given (biobank, ancestry) pair; such comparisons are indicated by an $\times$. One-sided $P$-values were calculated to test for positive correlation, and nominally significant correlations ($P < 0.05$) are outlined in black. Correlation coefficients are colored according to the legend. Traits are grouped by continuous and binary phenotypes and ordered alphabetically along the $x$-axis. Biobank comparisons are ordered alphabetically from bottom to top along the $y$-axis.

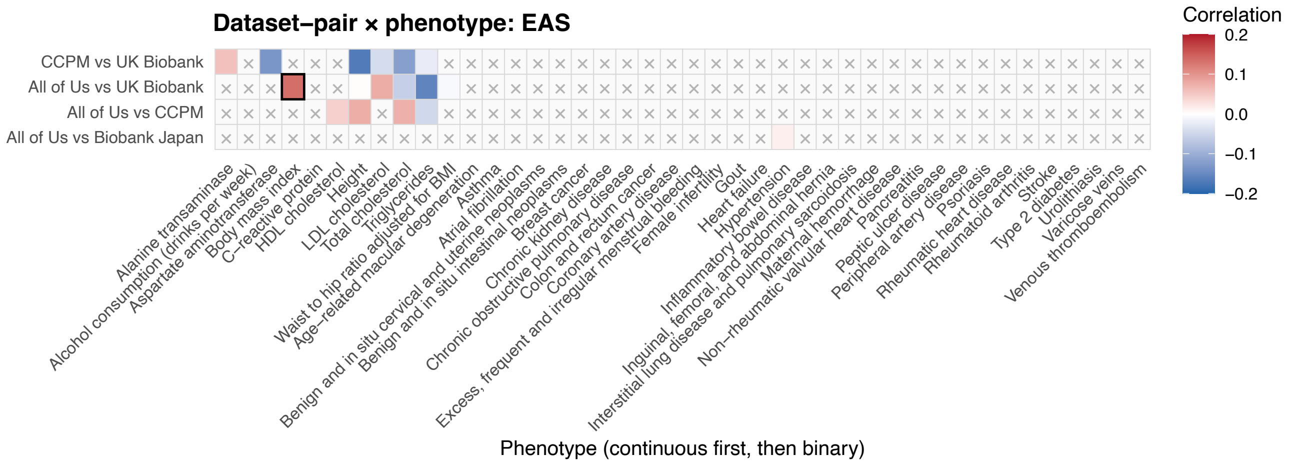

**Figure S18: Correlation between synonymous gene-level associations under the MAF $< 0.1\%$ mask in EAS.** For each trait, we compared gene-level synonymous burden test $Z$-scores (MAF $< 0.1\%$) between biobanks. Genes were excluded from correlation analyses if the cumulative minor allele count was $< 50$ for continuous traits or $< 30$ among cases for binary traits. Correlations were not computed when fewer than 100 genes met inclusion criteria, or when the phenotype was unavailable for a given (biobank, ancestry) pair; such comparisons are indicated by an $\times$. One-sided $P$-values were calculated to test for positive correlation, and nominally significant correlations ($P < 0.05$) are outlined in black. Correlation coefficients are colored according to the legend. Traits are grouped by continuous and binary phenotypes and ordered alphabetically along the $x$-axis. Biobank comparisons are ordered alphabetically from bottom to top along the $y$-axis.

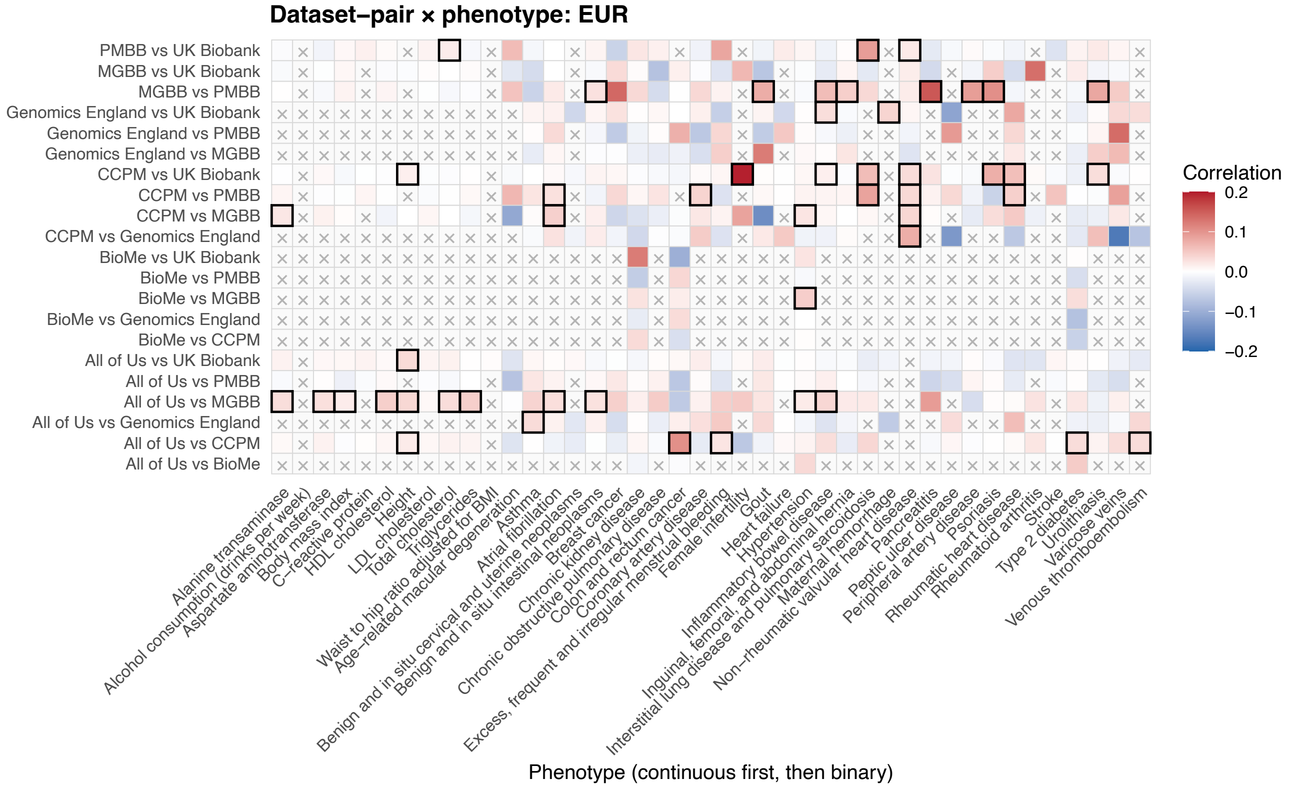

**Figure S19: Correlation between synonymous gene-level associations under the MAF $< 0.1\%$ mask in EUR.** For each trait, we compared gene-level synonymous burden test $Z$-scores (MAF $< 0.1\%$) between biobanks. Genes were excluded from correlation analyses if the cumulative minor allele count was $< 50$ for continuous traits or $< 30$ among cases for binary traits. Correlations were not computed when fewer than 100 genes met inclusion criteria, or when the phenotype was unavailable for a given (biobank, ancestry) pair; such comparisons are indicated by an $\times$. One-sided $P$-values were calculated to test for positive correlation, and nominally significant correlations ($P < 0.05$) are outlined in black. Correlation coefficients are colored according to the legend. Traits are grouped by continuous and binary phenotypes and ordered alphabetically along the $x$-axis. Biobank comparisons are ordered alphabetically from bottom to top along the $y$-axis.

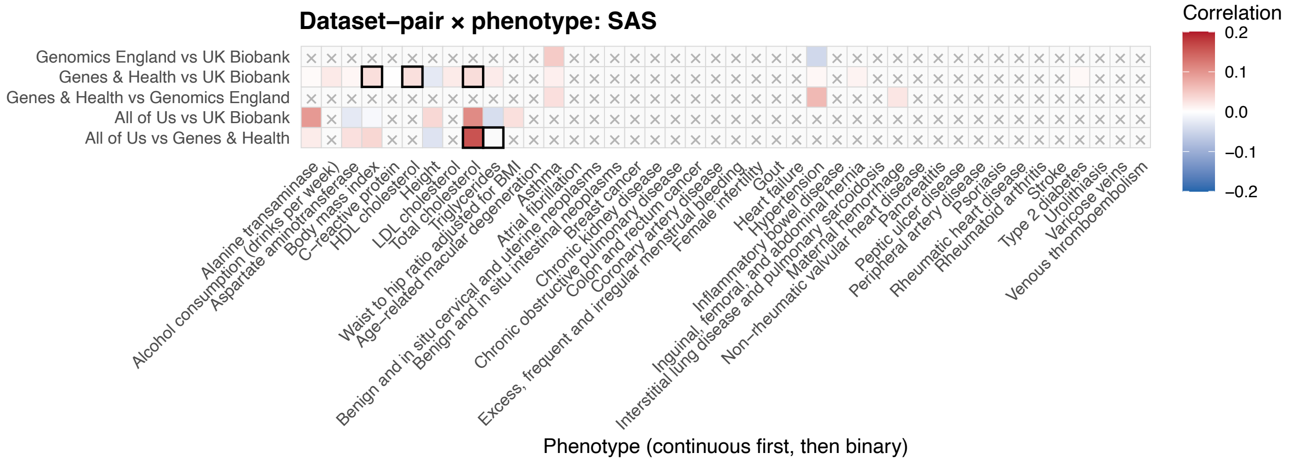

**Figure S20: Correlation between synonymous gene-level associations under the MAF $< 0.1\%$ mask in SAS.** For each trait, we compared gene-level synonymous burden test $Z$-scores (MAF $< 0.1\%$) between biobanks. Genes were excluded from correlation analyses if the cumulative minor allele count was $< 50$ for continuous traits or $< 30$ among cases for binary traits. Correlations were not computed when fewer than 100 genes met inclusion criteria, or when the phenotype was unavailable for a given (biobank, ancestry) pair; such comparisons are indicated by an $\times$. One-sided $P$-values were calculated to test for positive correlation, and nominally significant correlations ($P < 0.05$) are outlined in black. Correlation coefficients are colored according to the legend. Traits are grouped by continuous and binary phenotypes and ordered alphabetically along the $x$-axis. Biobank comparisons are ordered alphabetically from bottom to top along the $y$-axis.

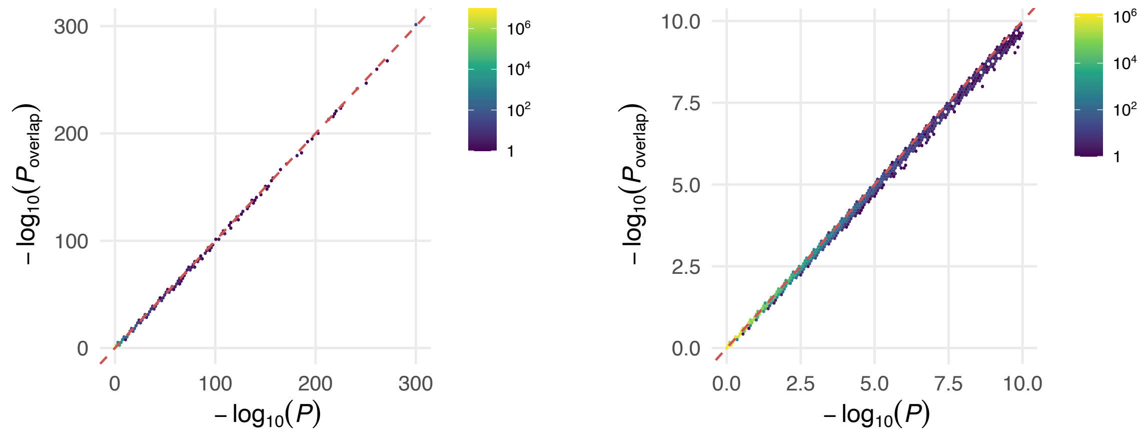

**Figure S21: Comparison of Stouffer based meta-analysis with and without accounting for sample overlap based on synonymous gene-level correlations** $-\log_{10}(P)$ values evaluated Stouffer's method with and without accounting for sample overlap according to the formula: $Z_{\text{meta}} = \frac{\sum_k \sqrt{N_{\text{eff},k}} Z_k}{\sqrt{\sum_k N_{\text{eff},k} + \sum_{k,l \neq k} \sqrt{N_{\text{eff},k} N_{\text{eff},l}} \hat{r}_{k,l}}}$ where $\hat{r}_{k,l}$ is the estimated correlation between the $Z$-scores of synonymous (MAF $< 0.1\%$) associations between the $k^{\text{th}}$ and $l^{\text{th}}$ studies under the null if the associated $P$-value for a positive association is below 0.05, otherwise $\hat{r}_{k,l}$ is set to 1. Points are binned and colored according to the count of points within each bin according to the legend. The right plot zooms into the $[0, 10] \times [0, 10]$ region of the plot on the left.

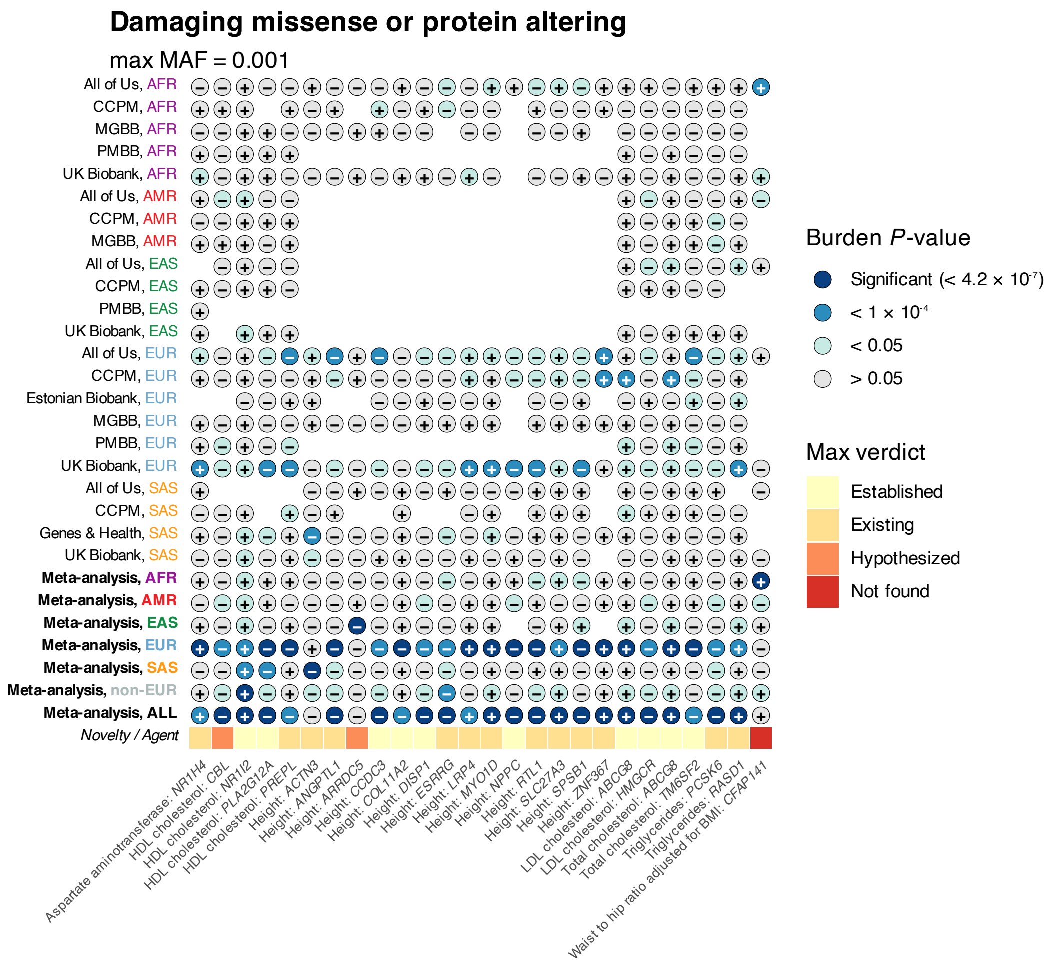

**Figure S22: Continuous, MAF $< 0.1\%$, 'damaging missense or protein altering' gene-level burden associations that are significant ($P < 0.05/(20,000 \cdot 3 \cdot 2) = 4.2 \times 10^{-7}$) through meta-analysis at the broad genetic ancestry level or across all (biobank, ancestry) tuples, but not significant in any single (biobank, ancestry) analysis.** Circles are colored according to significance of association for the corresponding (biobank, ancestry) or meta-analysis labeled on the $y$-axis, from light to dark according to the legend. The sign in the center of each circle displays the sign of the burden effect size (+ or −). The yellow to red squares in the final row shows the verdicts from an AI-agent (Lu et al., 2026) which indicates the level of evidence each association has been found previously.

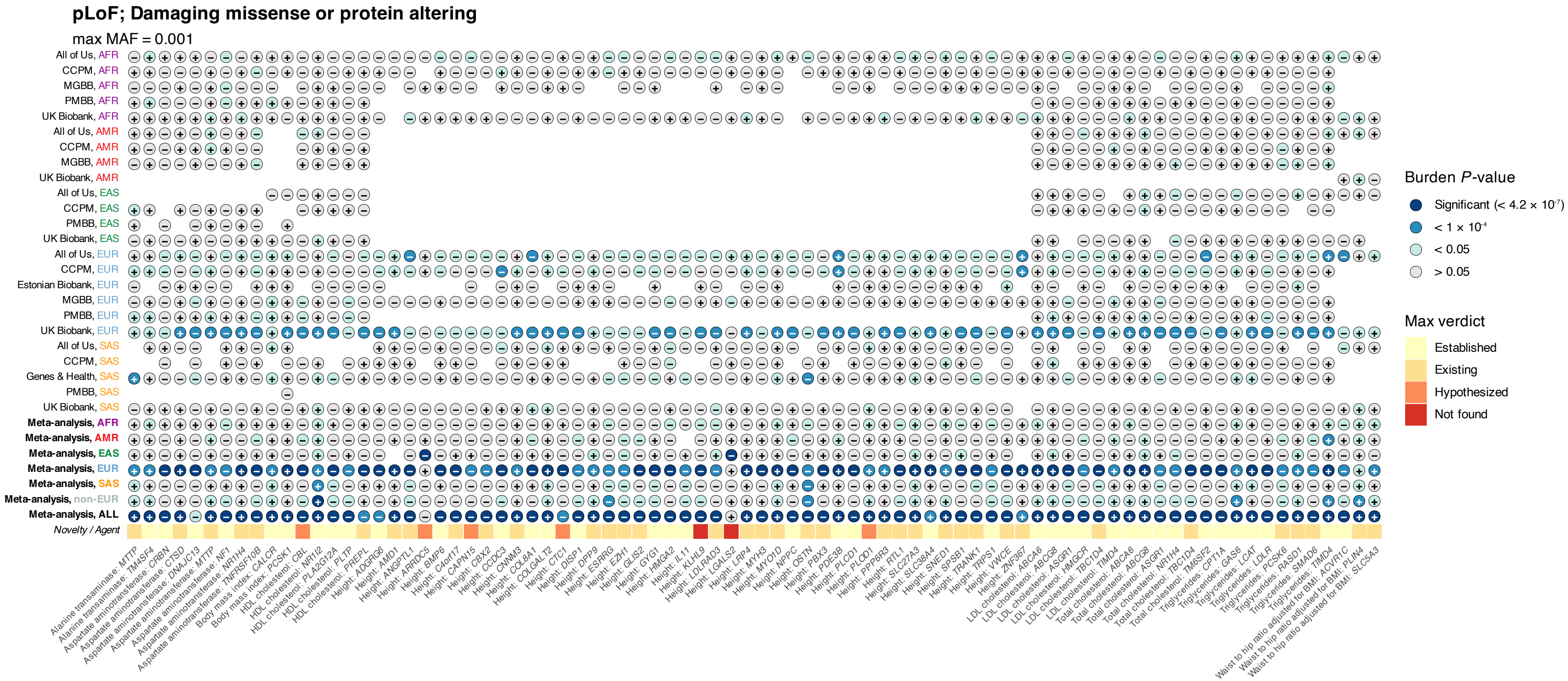

**Figure S23: Continuous, MAF < 0.1%, 'pLoF; damaging missense or protein altering' gene-level burden associations that are significant ($P < 0.05/(20,000 \cdot 3 \cdot 2) = 4.2 \times 10^{-7}$) through meta-analysis at the broad genetic ancestry level or across all (biobank, ancestry) tuples, but not significant in any single (biobank, ancestry) analysis.** Circles are colored according to significance of association for the corresponding (biobank, ancestry) or meta-analysis labeled on the $y$-axis, from light to dark according to the legend. The sign in the center of each circle displays the sign of the burden effect size (+ or −). The yellow to red squares in the final row shows the verdicts from an AI-agent (Lu et al., 2026) which indicates the level of evidence each association has been found previously.

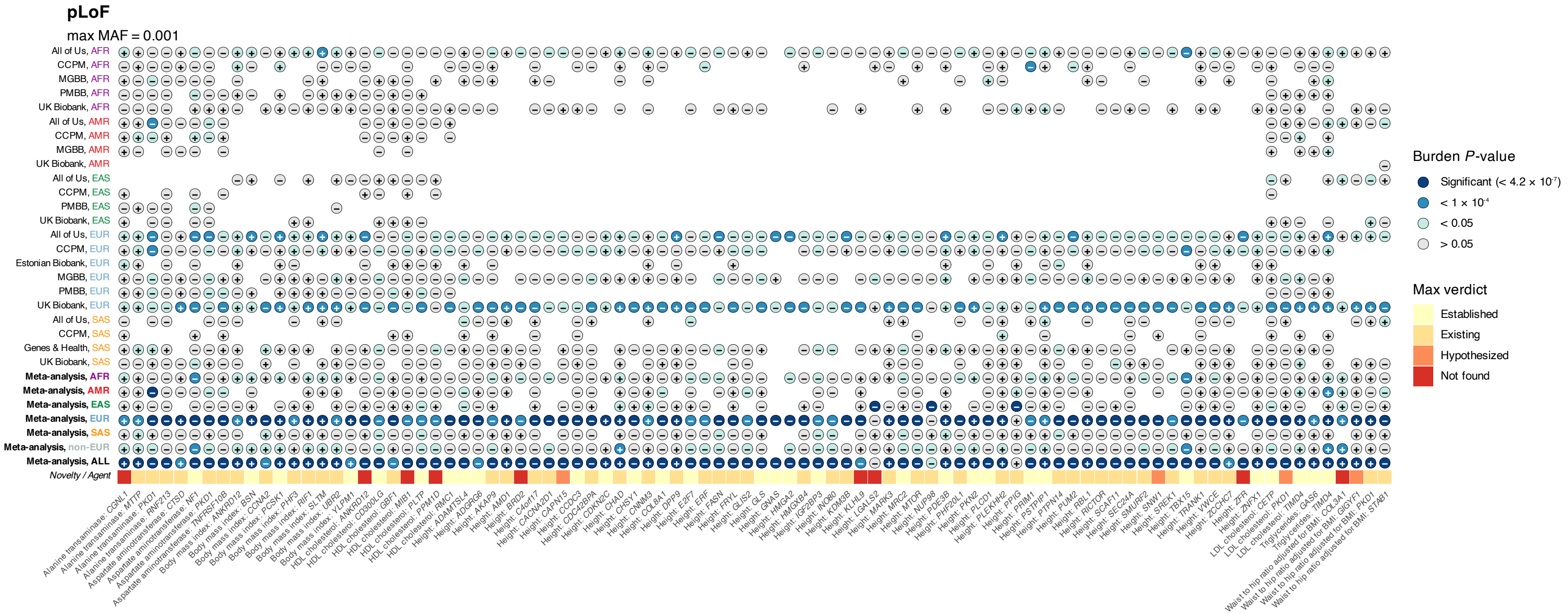

**Figure S24: Continuous, MAF $< 0.1\%$, pLoF gene-level burden associations that are significant ($P < 0.05/(20,000 \cdot 3 \cdot 2) = 4.2 \times 10^{-7}$) through meta-analysis at the broad genetic ancestry level or across all (biobank, ancestry) tuples, but not significant in any single (biobank, ancestry) analysis.** Circles are colored according to significance of association for the corresponding (biobank, ancestry) or meta-analysis labeled on the $y$-axis, from light to dark according to the legend. The sign in the center of each circle displays the sign of the burden effect size (+ or −). The yellow to red squares in the final row shows the verdicts from an AI-agent (Lu et al., 2026) which indicates the level of evidence each association has been found previously.

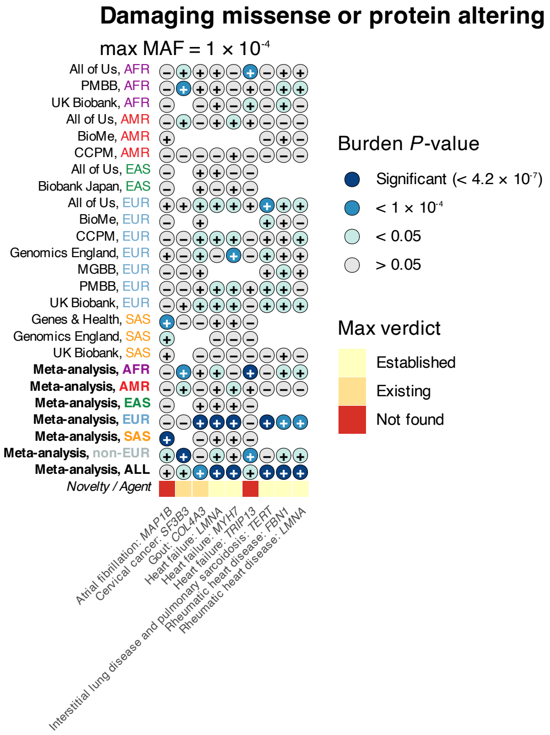

**Figure S25: Continuous, MAF $< 0.01\%$, 'damaging missense or protein altering' gene-level burden associations that are significant ($P < 0.05/(20,000 \cdot 3 \cdot 2) = 4.2 \times 10^{-7}$) through meta-analysis at the broad genetic ancestry level or across all (biobank, ancestry) tuples, but not significant in any single (biobank, ancestry) analysis.** Circles are colored according to significance of association for the corresponding (biobank, ancestry) or meta-analysis labeled on the $y$-axis, from light to dark according to the legend. The sign in the center of each circle displays the sign of the burden effect size (+ or −). The yellow to red squares in the final row shows the verdicts from an AI-agent (Lu et al., 2026) which indicates the level of evidence each association has been found previously.

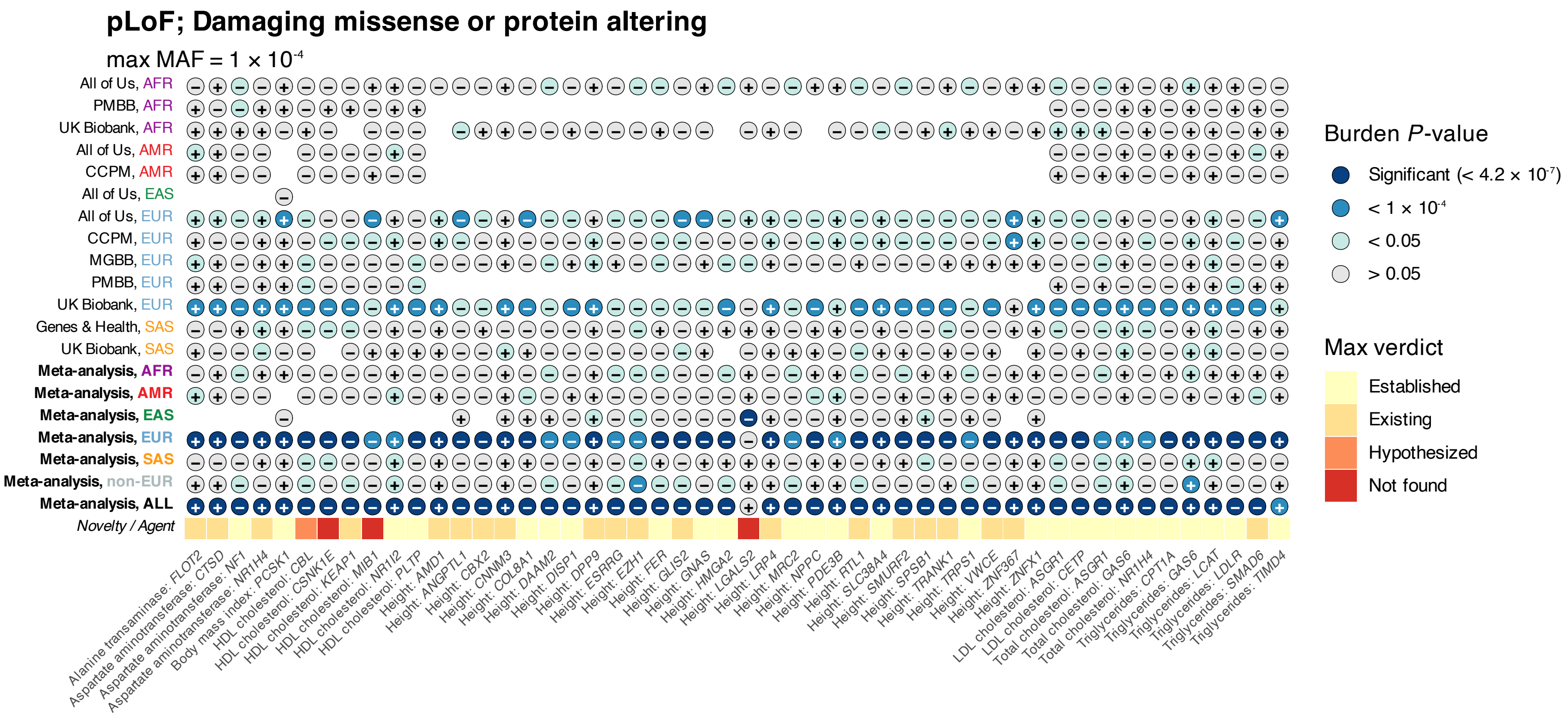

**Figure S26: Continuous, MAF $< 0.01\%$, 'pLoF; damaging missense or protein altering' gene-level burden associations that are significant ($P < 0.05/(20,000 \cdot 3 \cdot 2) = 4.2 \times 10^{-7}$) through meta-analysis at the broad genetic ancestry level or across all (biobank, ancestry) tuples, but not significant in any single (biobank, ancestry) analysis.** Circles are colored according to significance of association for the corresponding (biobank, ancestry) or meta-analysis labeled on the $y$-axis, from light to dark according to the legend. The sign in the center of each circle displays the sign of the burden effect size (+ or −). The yellow to red squares in the final row shows the verdicts from an AI-agent (Lu et al., 2026) which indicates the level of evidence each association has been found previously.

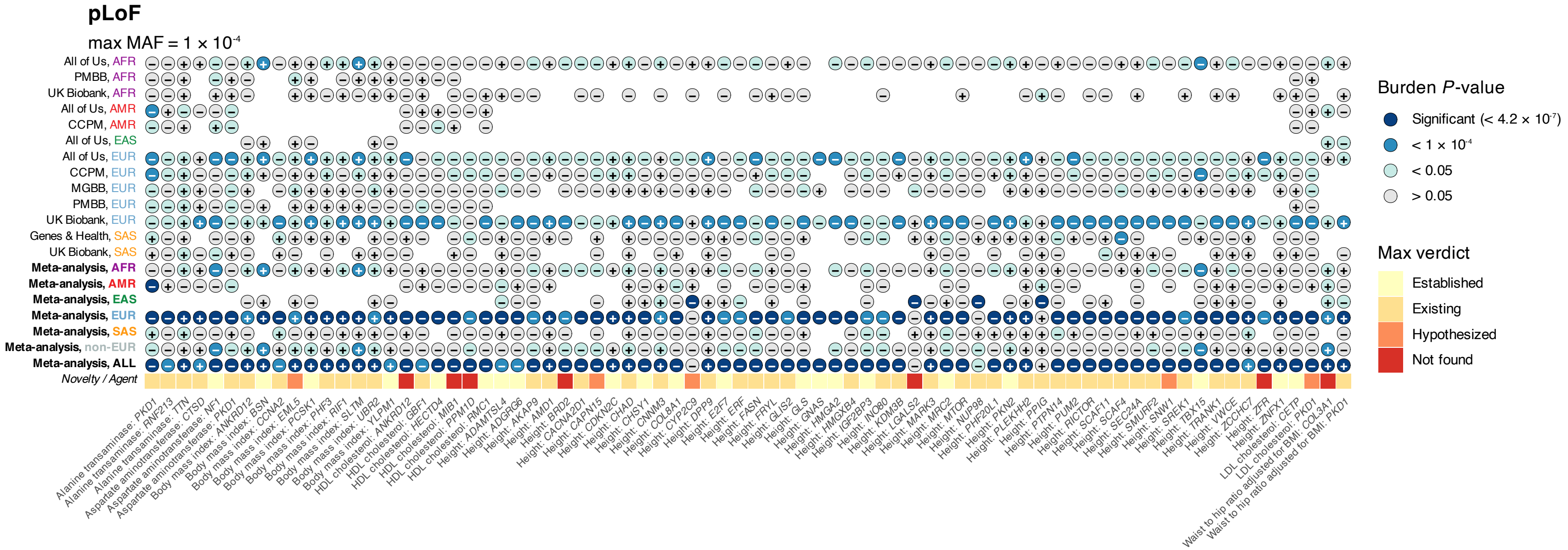

**Figure S27: Continuous, MAF $< 0.01\%$, pLoF gene-level burden associations that are significant ($P < 0.05/(20,000 \cdot 3 \cdot 2) = 4.2 \times 10^{-7}$) through meta-analysis at the broad genetic ancestry level or across all (biobank, ancestry) tuples, but not significant in any single (biobank, ancestry) analysis.** Circles are colored according to significance of association for the corresponding (biobank, ancestry) or meta-analysis labeled on the $y$-axis, from light to dark according to the legend. The sign in the center of each circle displays the sign of the burden effect size (+ or −). The yellow to red squares in the final row shows the verdicts from an AI-agent (Lu et al., 2026) which indicates the level of evidence each association has been found previously.

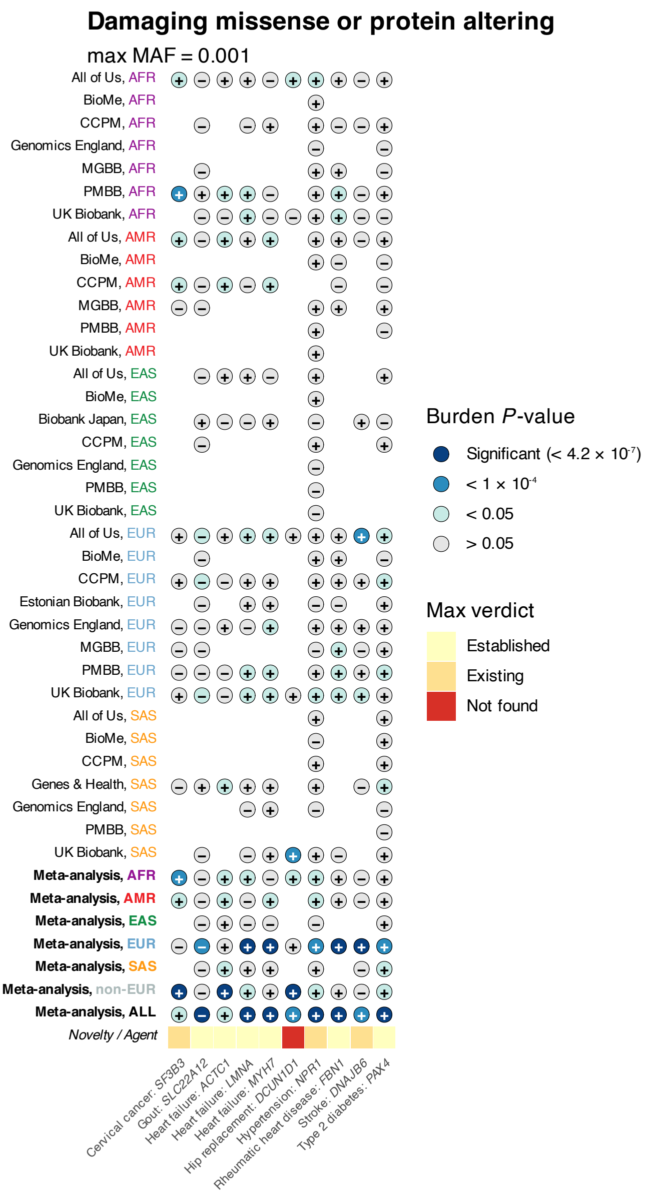

**Figure S28: Binary, MAF $< 0.1\%$, 'damaging missense or protein altering' gene-level burden associations that are significant ($P < 0.05/(20,000 \cdot 3 \cdot 2) = 4.2 \times 10^{-7}$) through meta-analysis at the broad genetic ancestry level or across all (biobank, ancestry) tuples, but not significant in any single (biobank, ancestry) analysis.** Circles are colored according to significance of association for the corresponding (biobank, ancestry) or meta-analysis labeled on the $y$-axis, from light to dark according to the legend. The sign in the center of each circle displays the sign of the burden effect size (+ or −). The yellow to red squares in the final row shows the verdicts from an AI-agent (Lu et al., 2026) which indicates the level of evidence each association has been found previously.

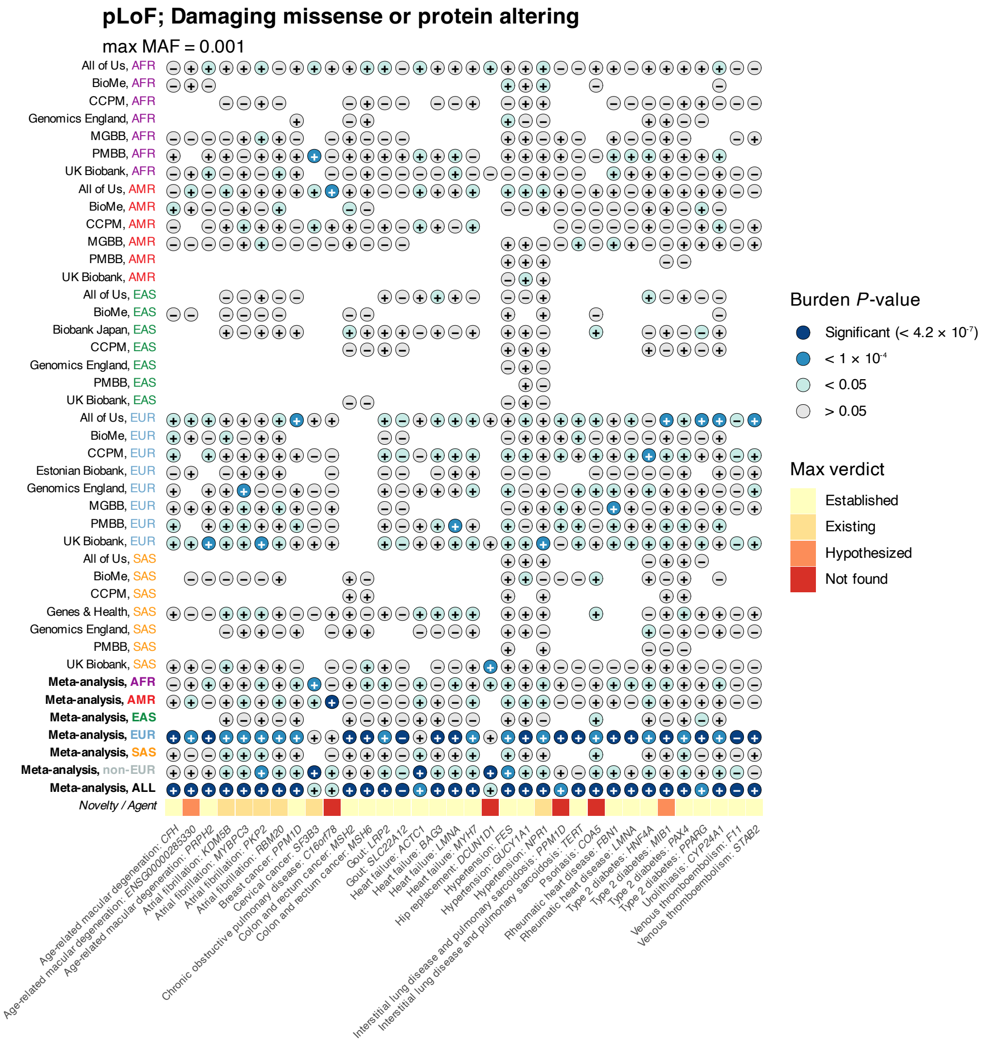

**Figure S29: Binary, MAF $< 0.1\%$, 'pLoF; damaging missense or protein altering' gene-level burden associations that are significant ($P < 0.05/(20,000 \cdot 3 \cdot 2) = 4.2 \times 10^{-7}$) through meta-analysis at the broad genetic ancestry level or across all (biobank, ancestry) tuples, but not significant in any single (biobank, ancestry) analysis.** Circles are colored according to significance of association for the corresponding (biobank, ancestry) or meta-analysis labeled on the $y$-axis, from light to dark according to the legend. The sign in the center of each circle displays the sign of the burden effect size (+ or −). The yellow to red squares in the final row shows the verdicts from an AI-agent (Lu et al., 2026) which indicates the level of evidence each association has been found previously.

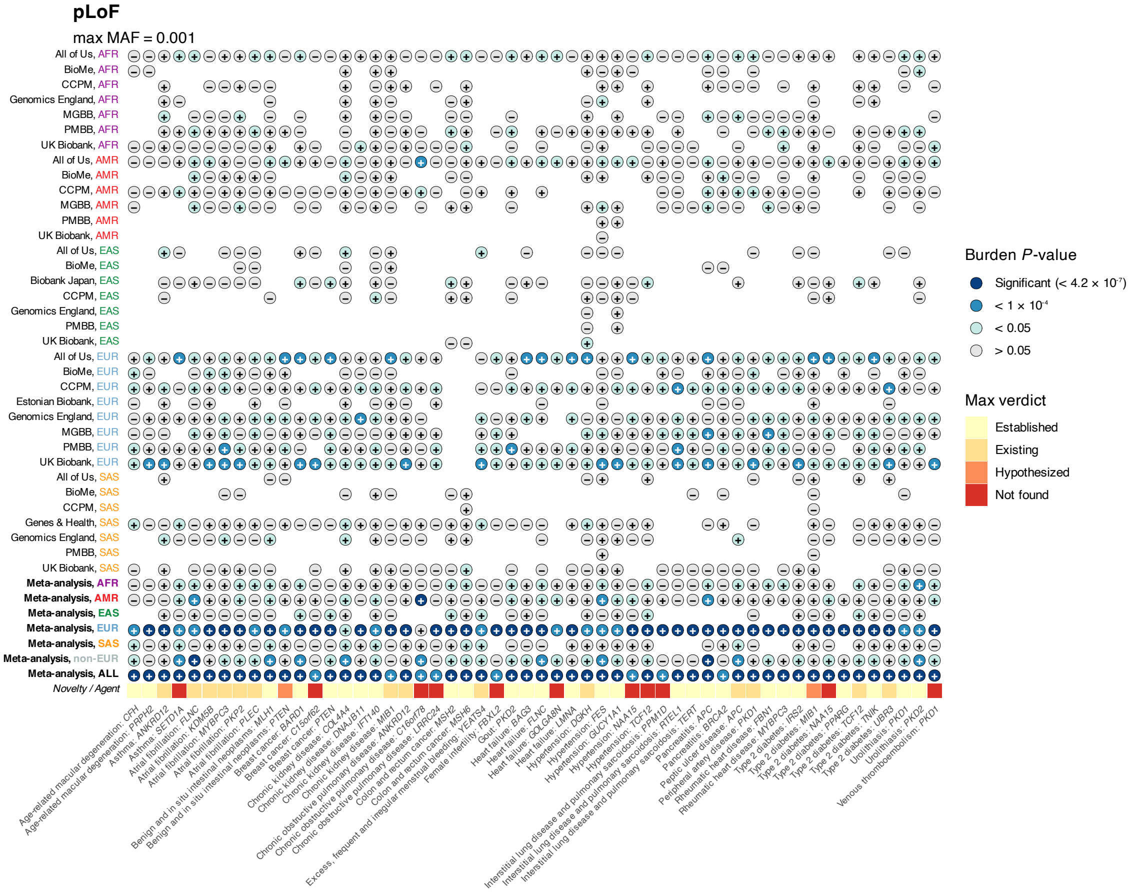

**Figure S30: Binary, MAF $< 0.1\%$, pLoF gene-level burden associations that are significant ($P < 0.05/(20,000 \cdot 3 \cdot 2) = 4.2 \times 10^{-7}$) through meta-analysis at the broad genetic ancestry level or across all (biobank, ancestry) tuples, but not significant in any single (biobank, ancestry) analysis.** Circles are colored according to significance of association for the corresponding (biobank, ancestry) or meta-analysis labeled on the $y$-axis, from light to dark according to the legend. The sign in the center of each circle displays the sign of the burden effect size (+ or −). The yellow to red squares in the final row shows the verdicts from an AI-agent (Lu et al., 2026) which indicates the level of evidence each association has been found previously.

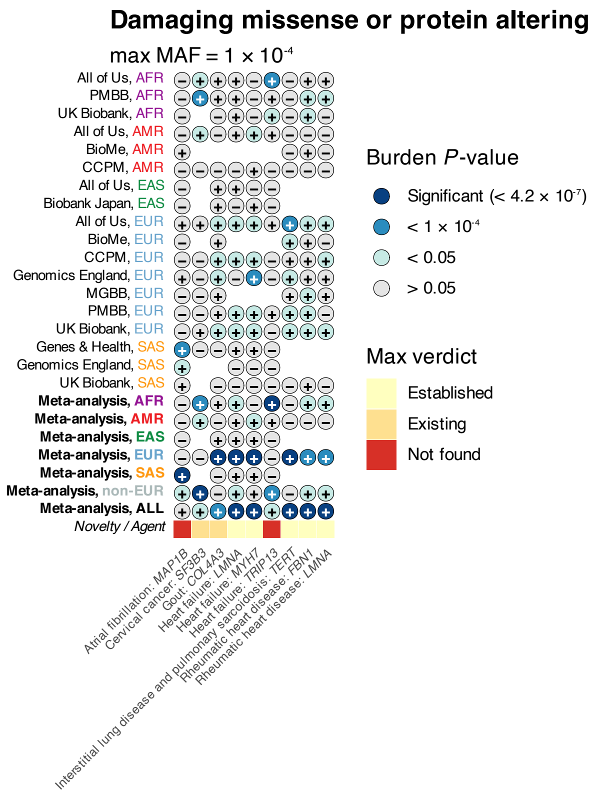

**Figure S31: Binary, MAF $< 0.01\%$, 'damaging missense or protein altering' gene-level burden associations that are significant ($P < 0.05/(20,000 \cdot 3 \cdot 2) = 4.2 \times 10^{-7}$) through meta-analysis at the broad genetic ancestry level or across all (biobank, ancestry) tuples, but not significant in any single (biobank, ancestry) analysis.** Circles are colored according to significance of association for the corresponding (biobank, ancestry) or meta-analysis labeled on the $y$-axis, from light to dark according to the legend. The sign in the center of each circle displays the sign of the burden effect size (+ or −). The yellow to red squares in the final row shows the verdicts from an AI-agent (Lu et al., 2026) which indicates the level of evidence each association has been found previously.

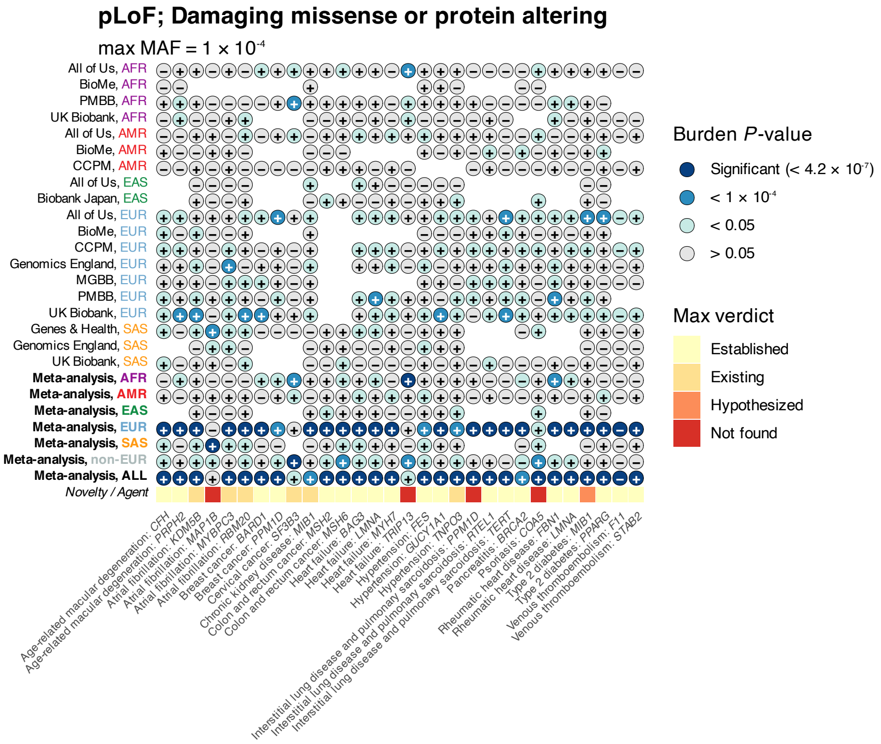

**Figure S32: Binary, MAF $< 0.01\%$, 'pLoF; damaging missense or protein altering' gene-level burden associations that are significant ($P < 0.05/(20,000 \cdot 3 \cdot 2) = 4.2 \times 10^{-7}$) through meta-analysis at the broad genetic ancestry level or across all (biobank, ancestry) tuples, but not significant in any single (biobank, ancestry) analysis.** Circles are colored according to significance of association for the corresponding (biobank, ancestry) or meta-analysis labeled on the $y$-axis, from light to dark according to the legend. The sign in the center of each circle displays the sign of the burden effect size (+ or −). The yellow to red squares in the final row shows the verdicts from an AI-agent (Lu et al., 2026) which indicates the level of evidence each association has been found previously.

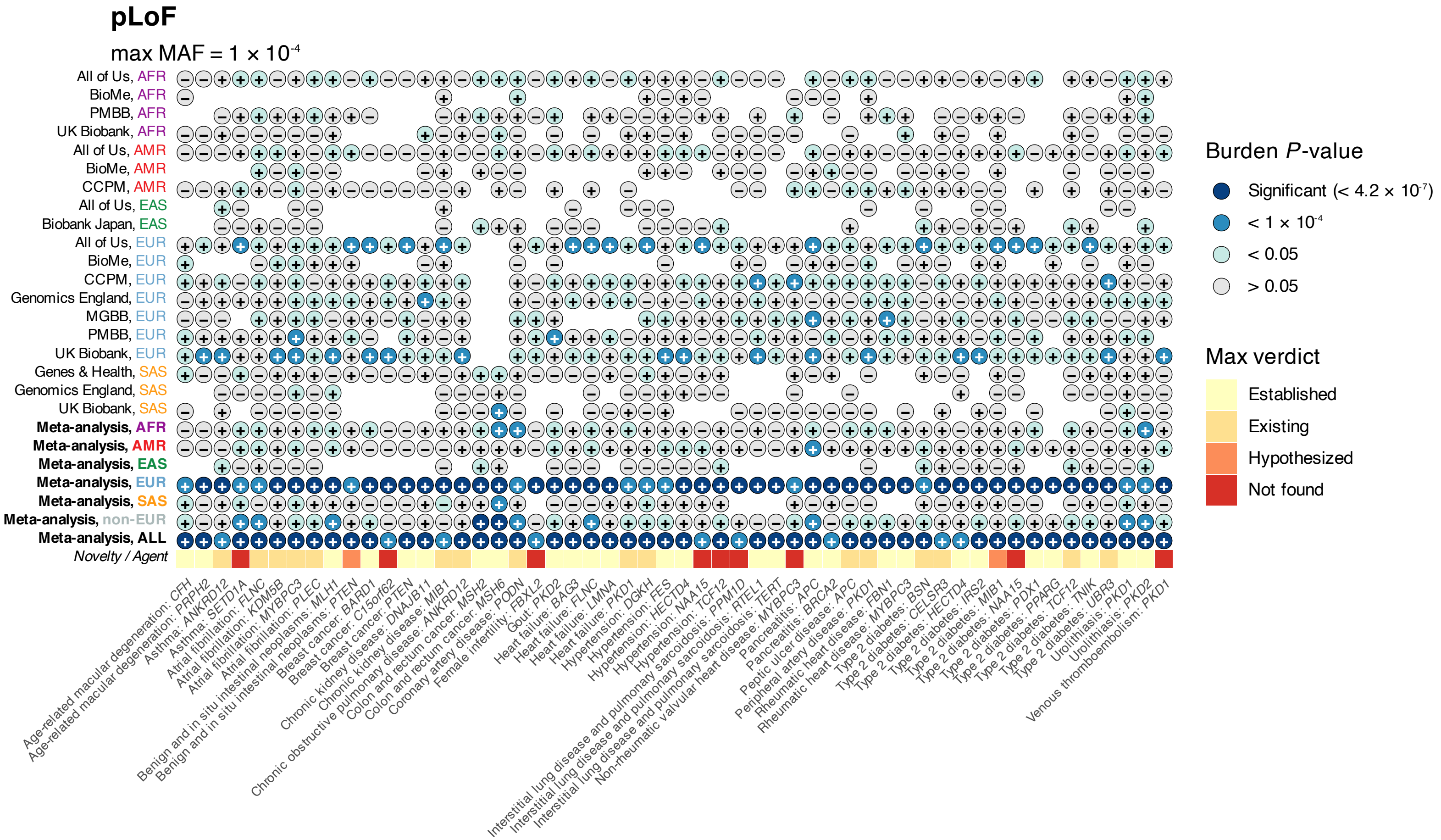

**Figure S33: Binary, MAF $< 0.01\%$, pLoF gene-level burden associations that are significant** ($P < 0.05/(20,000 \cdot 3 \cdot 2) = 4.2 \times 10^{-7}$) **through meta-analysis at the broad genetic ancestry level or across all (biobank, ancestry) tuples, but not significant in any single (biobank, ancestry) analysis.** Circles are colored according to significance of association for the corresponding (biobank, ancestry) or meta-analysis labeled on the $y$-axis, from light to dark according to the legend. The sign in the center of each circle displays the sign of the burden effect size (+ or −). The yellow to red squares in the final row shows the verdicts from an AI-agent (Lu et al., 2026) which indicates the level of evidence each association has been found previously.

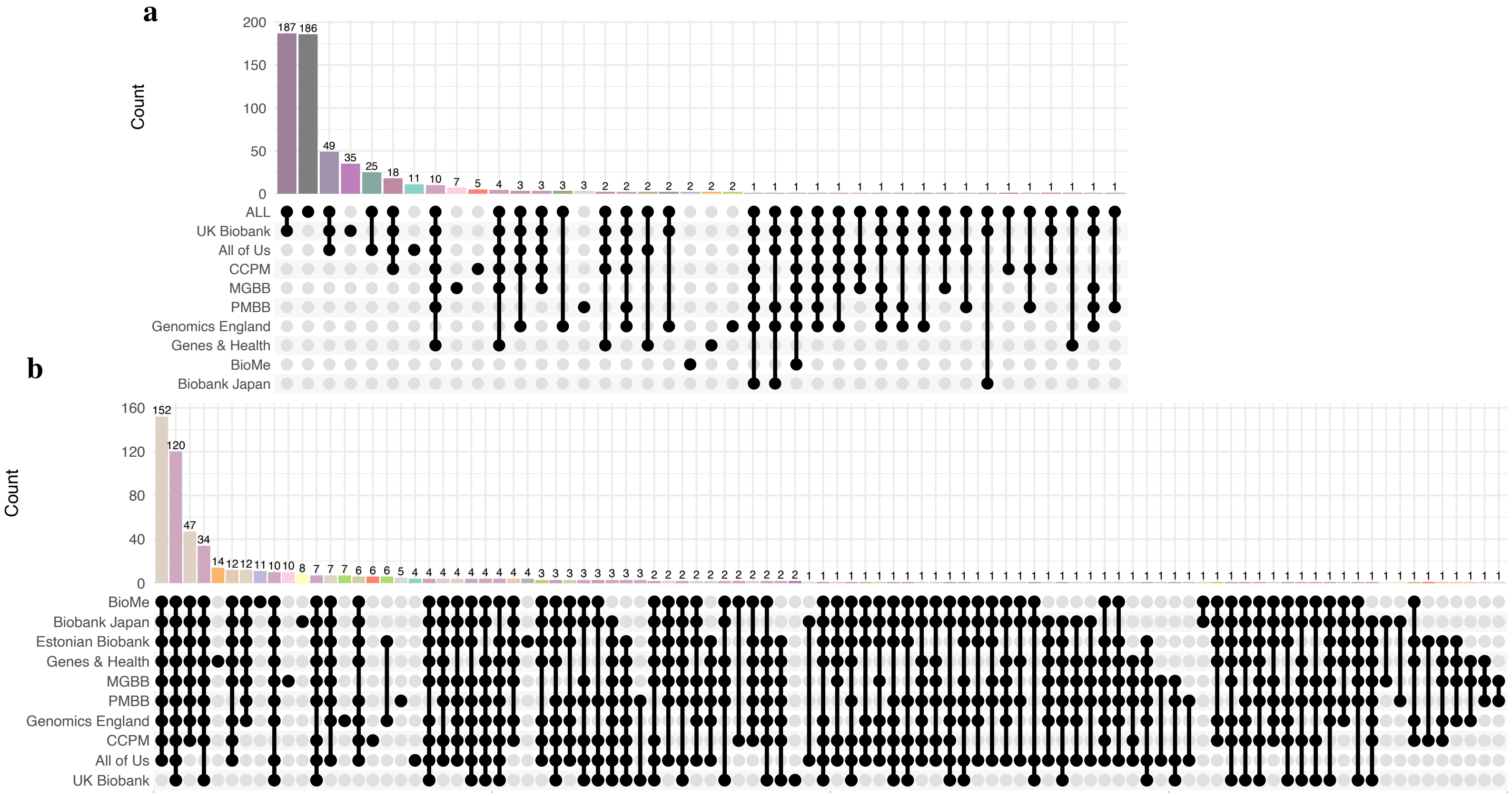

**Figure S34: Biobank-specific and leave-one-biobank-out meta-analysis results.** a, Upset plot showing counts of unique experiment-wise significant gene-trait associations identified in biobank-specific meta-analyses, performed by meta-analyzing across genetic ancestry groups within each biobank and aggregating evidence across tests for each gene using the Cauchy combination test (Cauchy $P < 2.5 \times 10^{-6}$). 'ALL' denotes the meta-analysis across all (biobank, ancestry) pairs. b, Corresponding upset plot showing counts of unique experiment-wise significant gene-trait associations identified in leave-one-biobank-out meta-analyses across all (biobank, ancestry) pairs, followed by Cauchy combination (Cauchy $P < 2.5 \times 10^{-6}$). Vertical bars indicate the number of associations unique to or shared among the indicated meta-analyses.

### AFR

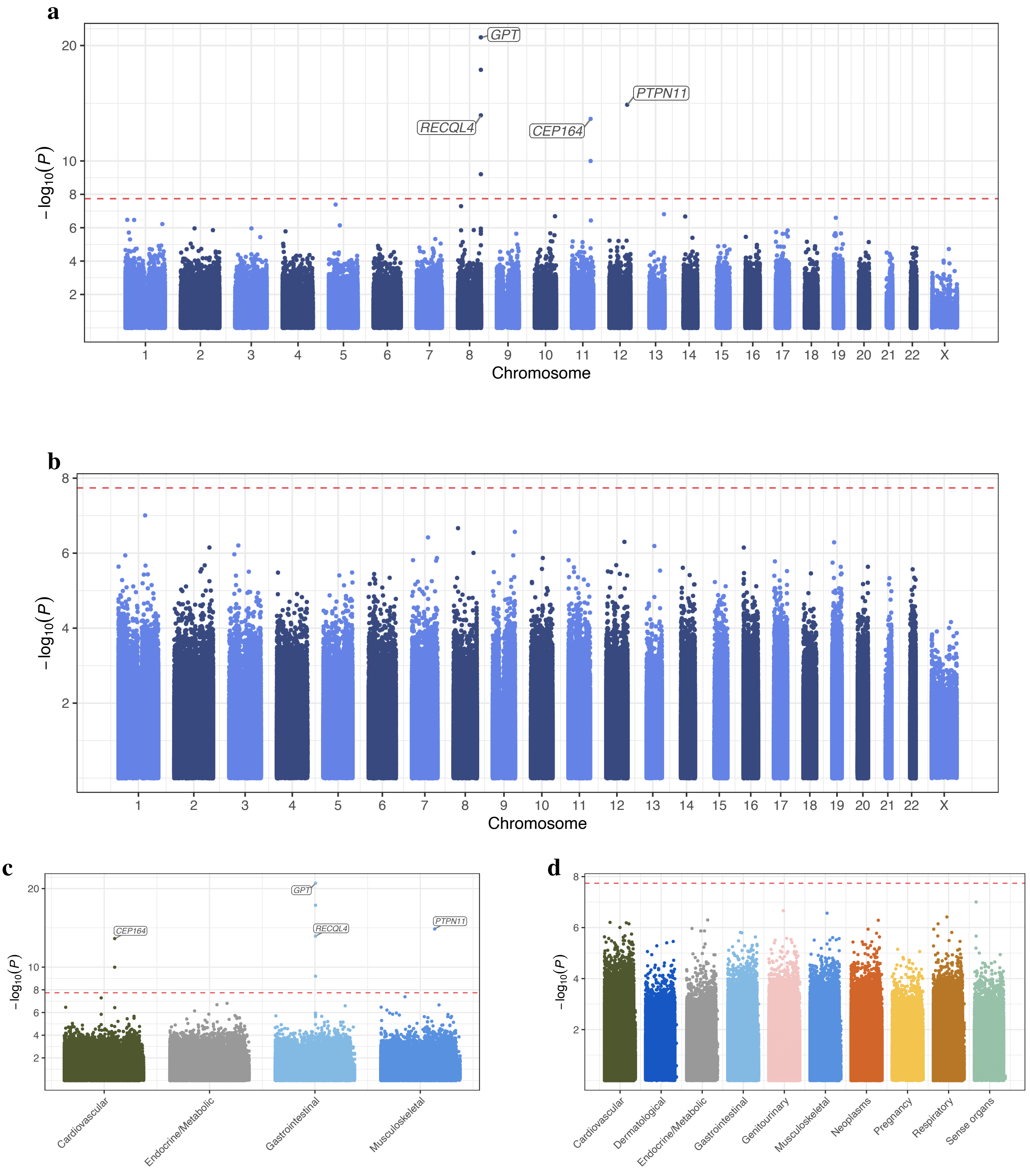

**Figure S35: Meta-analysis of AFR genetic ancestry individuals across 10 global biobanks: variant level, MAF $< 0.1\%$.** a-b, Summary of variant level analysis results for continuous and binary traits, respectively. The red dotted horizontal line denotes the experiment-wise significance threshold for variant-level association testing ($P = 1.8 \times 10^{-8}$). c-d, display the corresponding plots, split by ICD chapter on the $x$-axis.

#### AMR

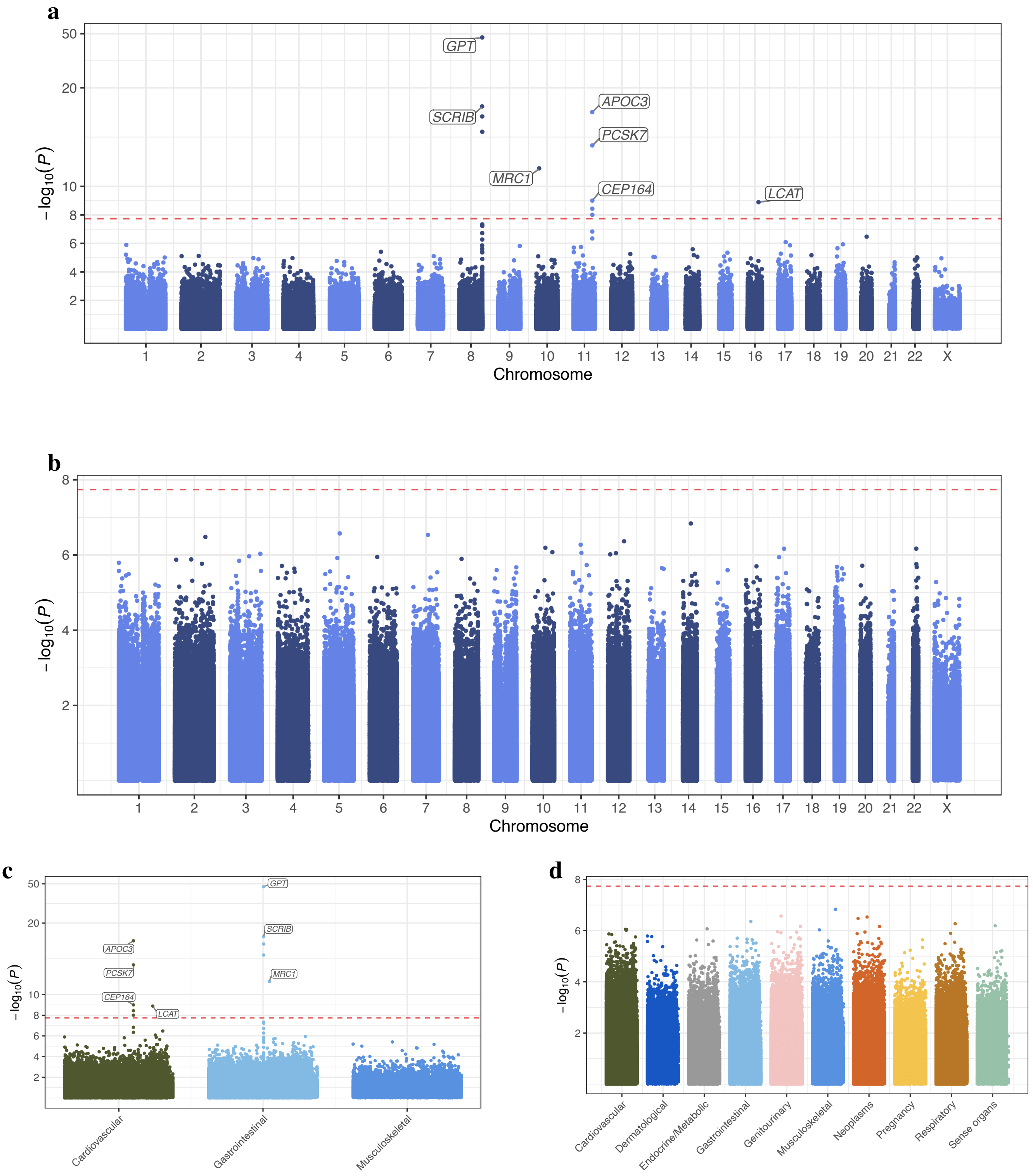

**Figure S36: Meta-analysis of AMR genetic ancestry individuals across 10 global biobanks: variant level, MAF < 0.1%.** a-b, Summary of variant level analysis results for continuous and binary traits, respectively. The red dotted horizontal line denotes the experiment-wise significance threshold for variant-level association testing ($P = 1.8 \times 10^{-8}$). c-d, display the corresponding plots, split by ICD chapter on the $x$-axis.

### EAS

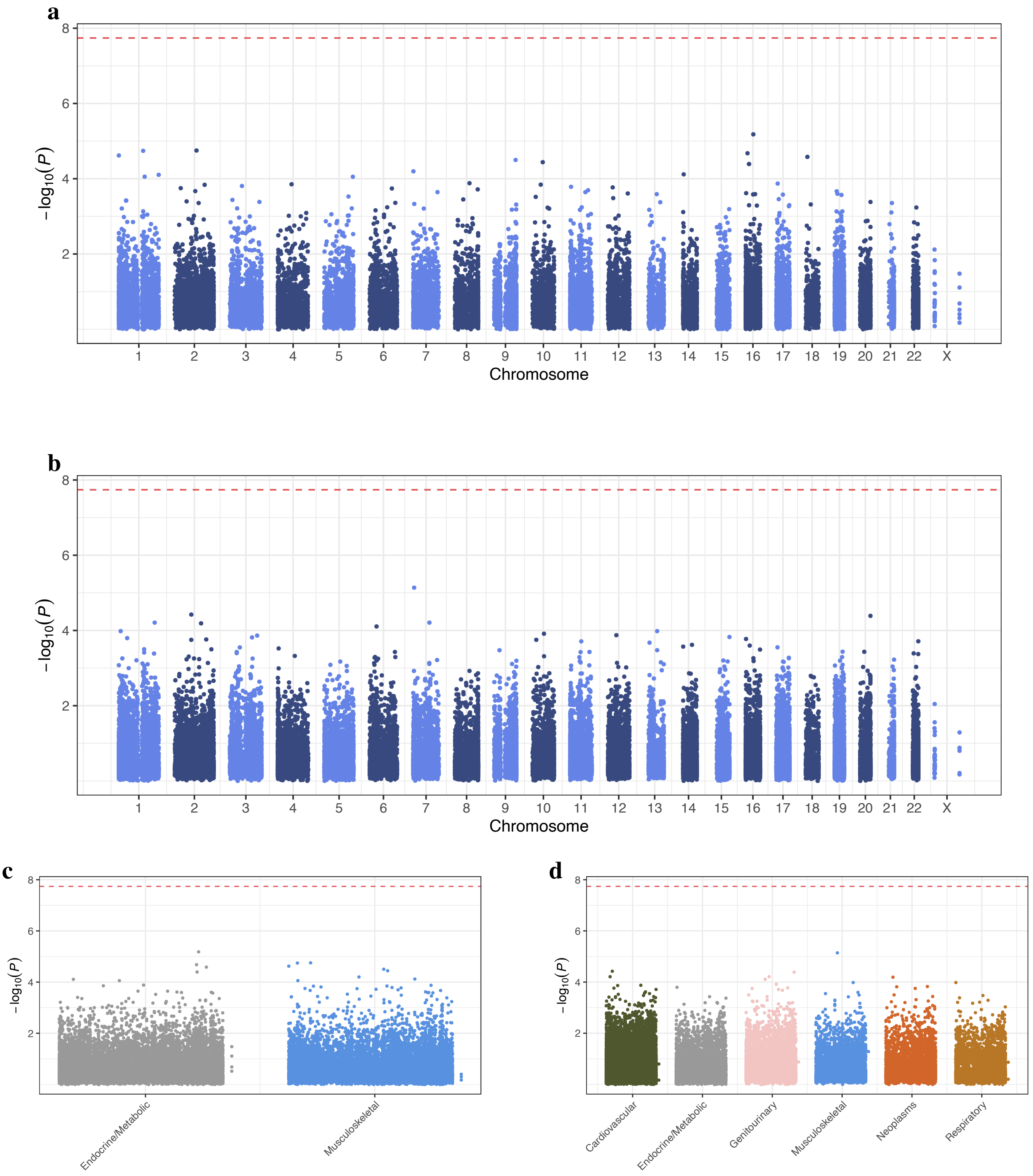

**Figure S37: Meta-analysis of EAS genetic ancestry individuals across 10 global biobanks: variant level, MAF $< 0.1\%$.** a-b, Summary of variant level analysis results for continuous and binary traits, respectively. The red dotted horizontal line denotes the experiment-wise significance threshold for variant-level association testing ($P = 1.8 \times 10^{-8}$). c-d, display the corresponding plots, split by ICD chapter on the $x$-axis.

### EUR

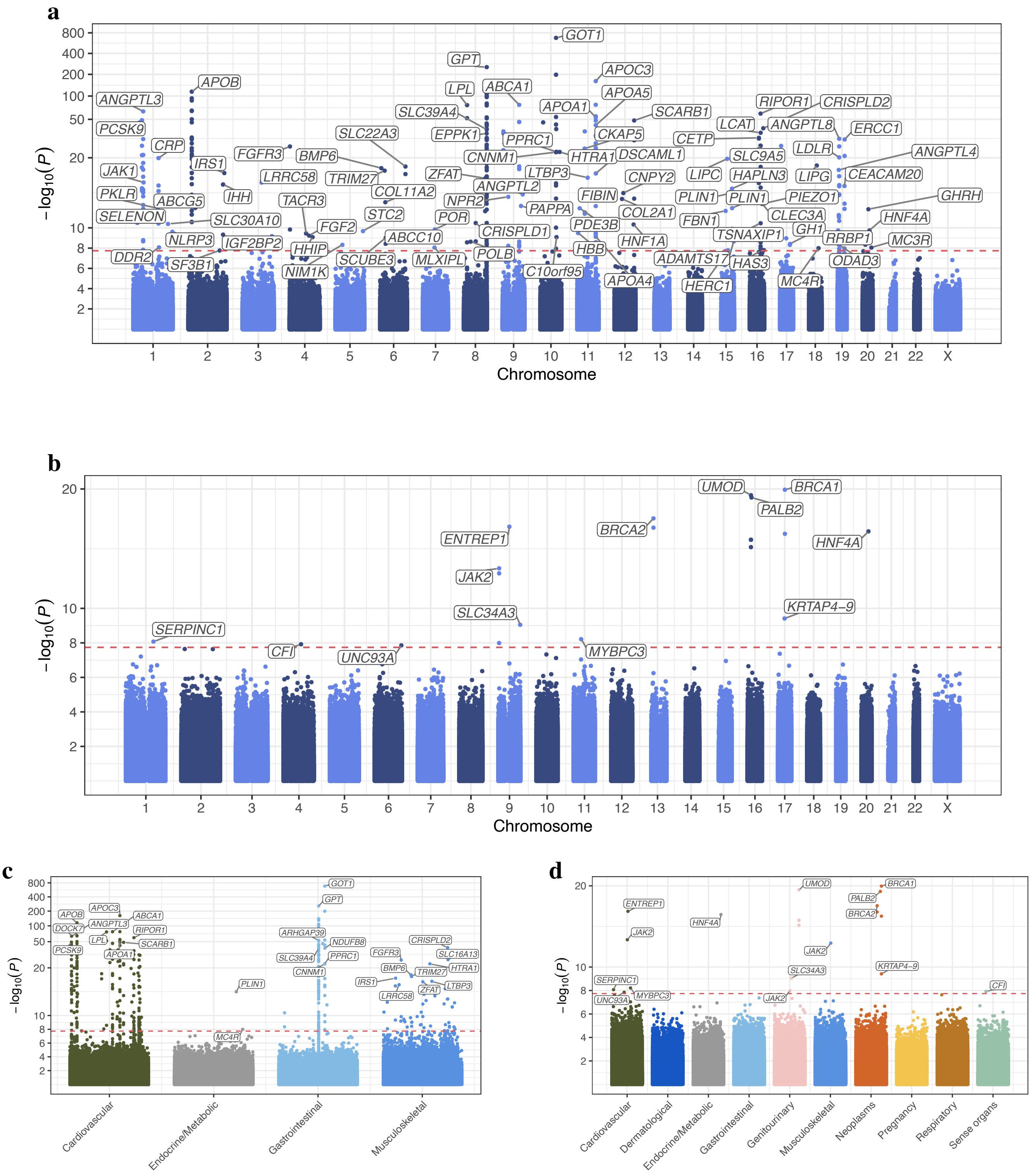

**Figure S38: Meta-analysis of EUR genetic ancestry individuals across 10 global biobanks: variant level, MAF $< 0.1\%$.** a-b, Summary of variant level analysis results for continuous and binary traits, respectively. The red dotted horizontal line denotes the experiment-wise significance threshold for variant-level association testing ($P = 1.8 \times 10^{-8}$). c-d, display the corresponding plots, split by ICD chapter on the $x$-axis.

#### SAS

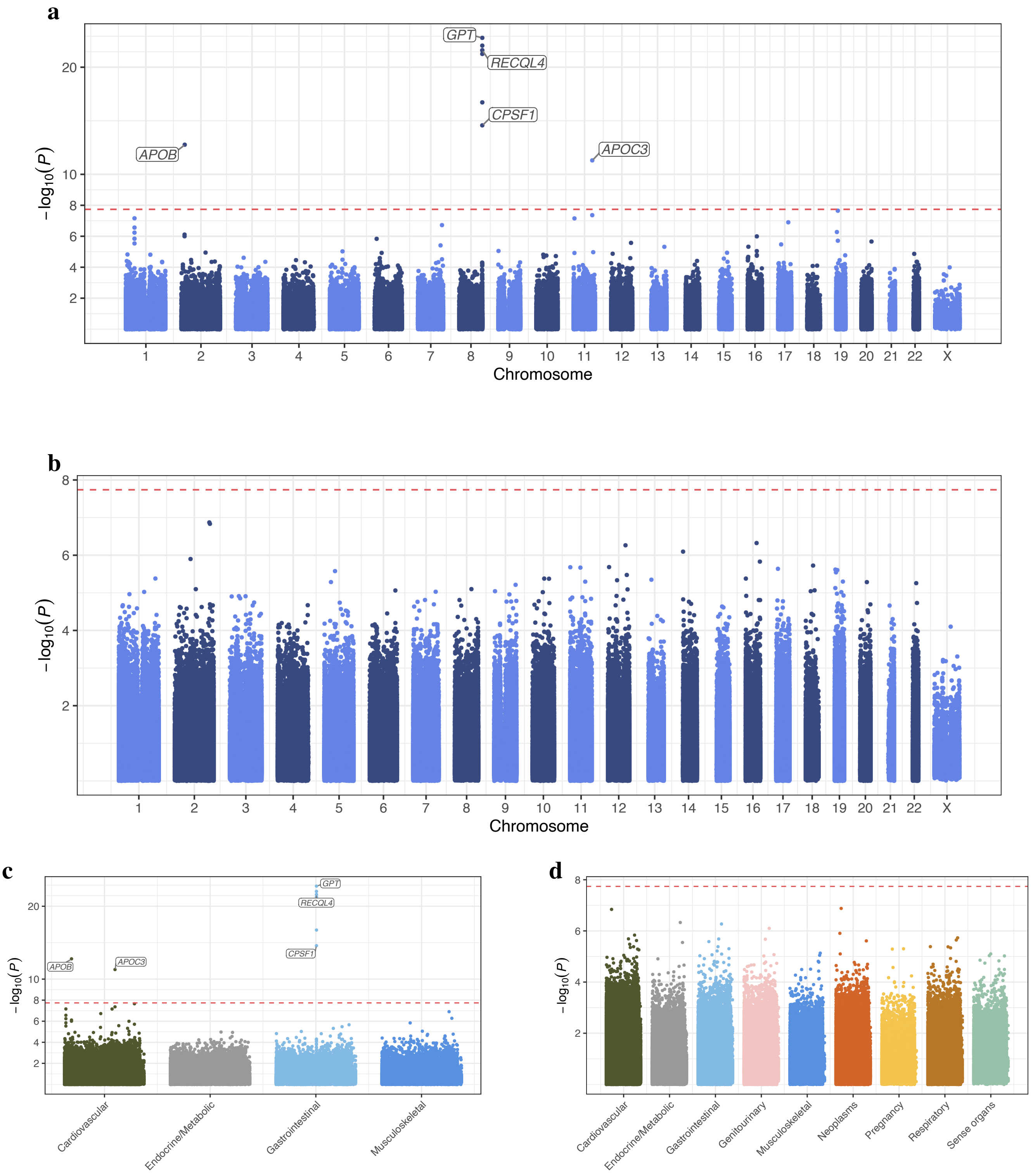

**Figure S39: Meta-analysis of SAS genetic ancestry individuals across 10 global biobanks: variant level, MAF $< 0.1\%$.** a-b, Summary of variant level analysis results for continuous and binary traits, respectively. The red dotted horizontal line denotes the experiment-wise significance threshold for variant-level association testing ($P = 1.8 \times 10^{-8}$). c-d, display the corresponding plots, split by ICD chapter on the $x$-axis.

#### non-EUR

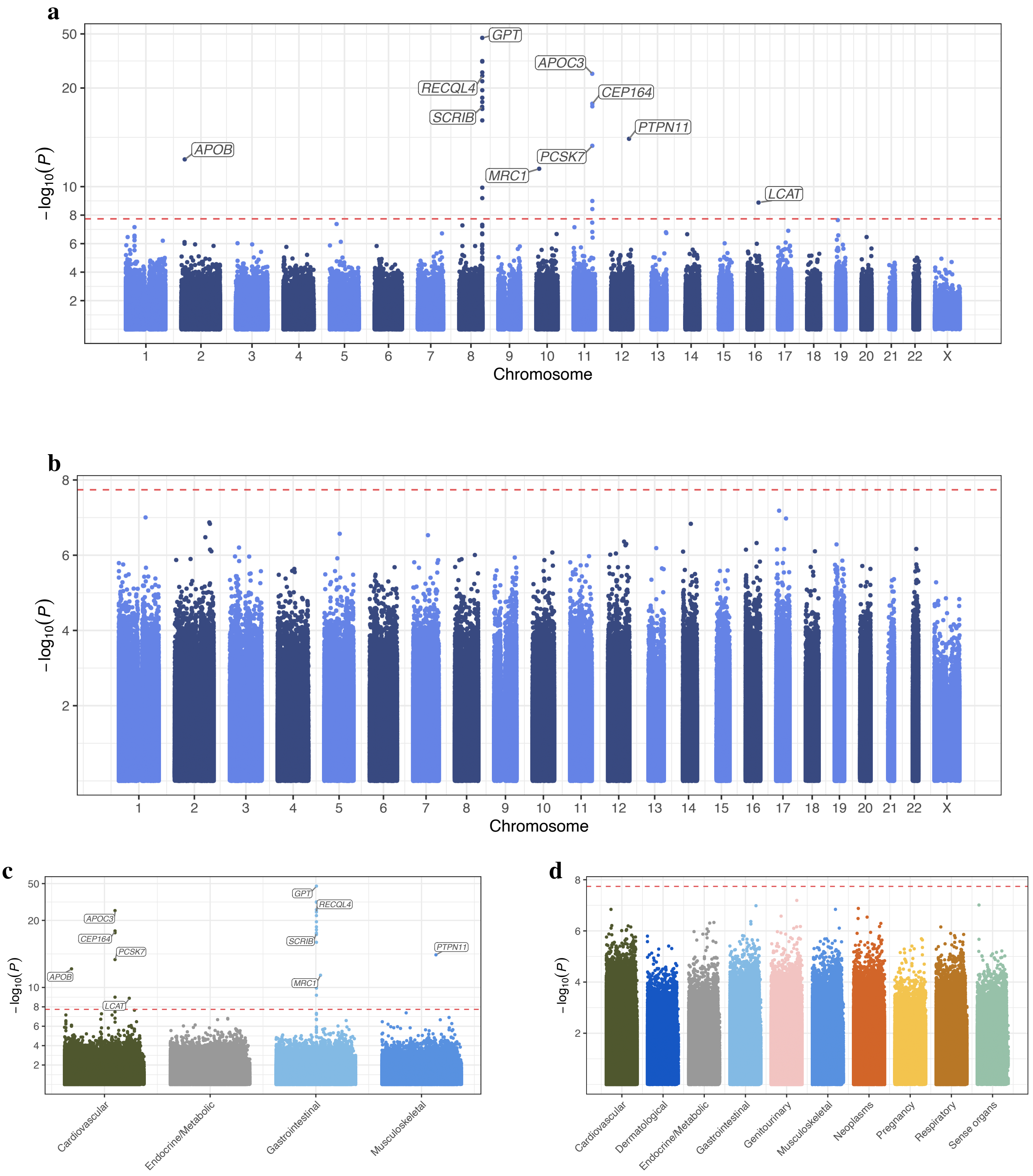

**Figure S40: Meta-analysis of non-EUR genetic ancestry individuals across 10 global biobanks: variant level, MAF < 0.1%.** a-b, Summary of variant level analysis results for continuous and binary traits, respectively. The red dotted horizontal line denotes the experiment-wise significance threshold for variant-level association testing ($P = 1.8 \times 10^{-8}$). c-d, display the corresponding plots, split by ICD chapter on the $x$-axis.

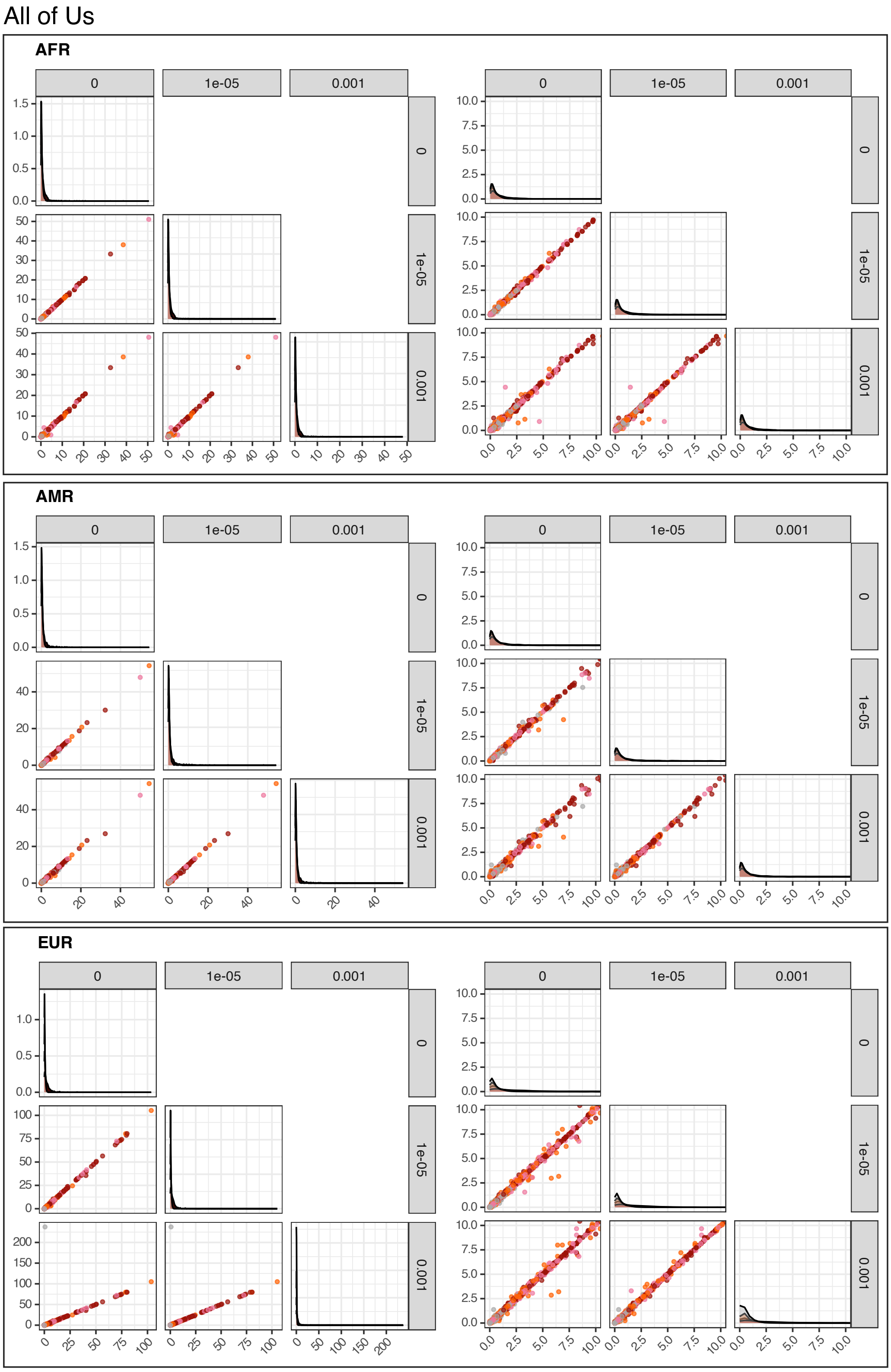

**Figure S41: Examining effect of choice of conditioning $P$-value threshold on results.** $-\log_{10}(P)$ values are plotted against each other after performing iterative conditioning for two $P$-value thresholds $(0.001, 1 \times 10^{-5})$, and excluding conditioning ($P$-value threshold of 0). Histograms on the diagonal show the distribution of association $P$-values for each $P$-value threshold. Dots are colored by annotation according to the legend. For (biobank, ancestry) pairs with at least one association $P < 1 \times 10^{-5}$ we zoom in on the $[0, 10] \times [0, 10]$ region of the plot on the left. Boxes delineate genetic ancestry subsets of the data.

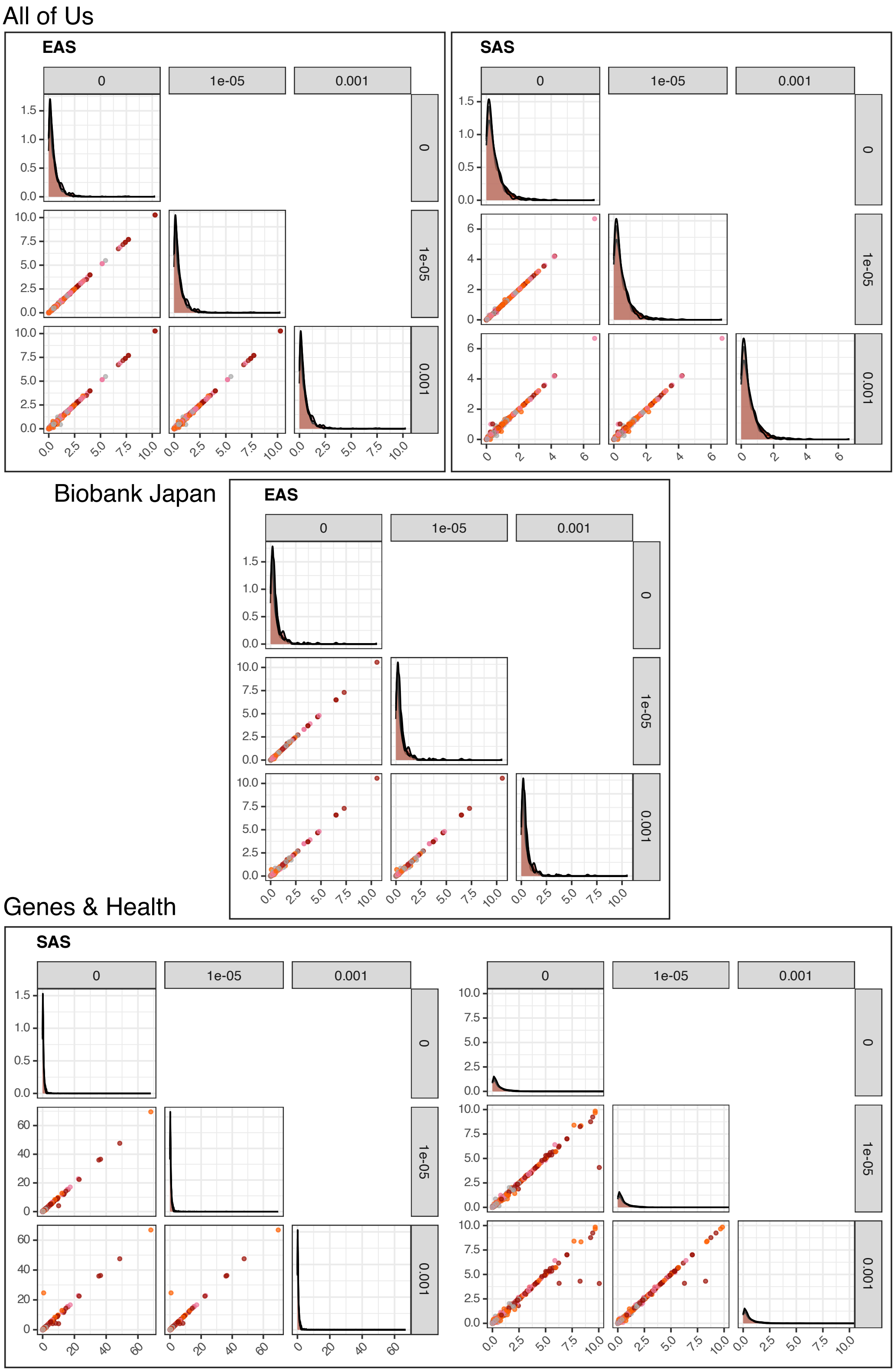

**Figure S42: Examining effect of choice of conditioning $P$-value threshold on results.** $-\log_{10}(P)$ values are plotted against each other after performing iterative conditioning for two $P$-value thresholds $(0.001, 1 \times 10^{-5})$, and excluding conditioning ($P$-value threshold of 0). Histograms on the diagonal show the distribution of association $P$-values for each $P$-value threshold. Dots are colored by annotation according to the legend. For (biobank, ancestry) pairs with at least one association $P < 1 \times 10^{-5}$ we zoom in on the $[0, 10] \times [0, 10]$ region of the plot on the left. Boxes delineate genetic ancestry subsets of the data.

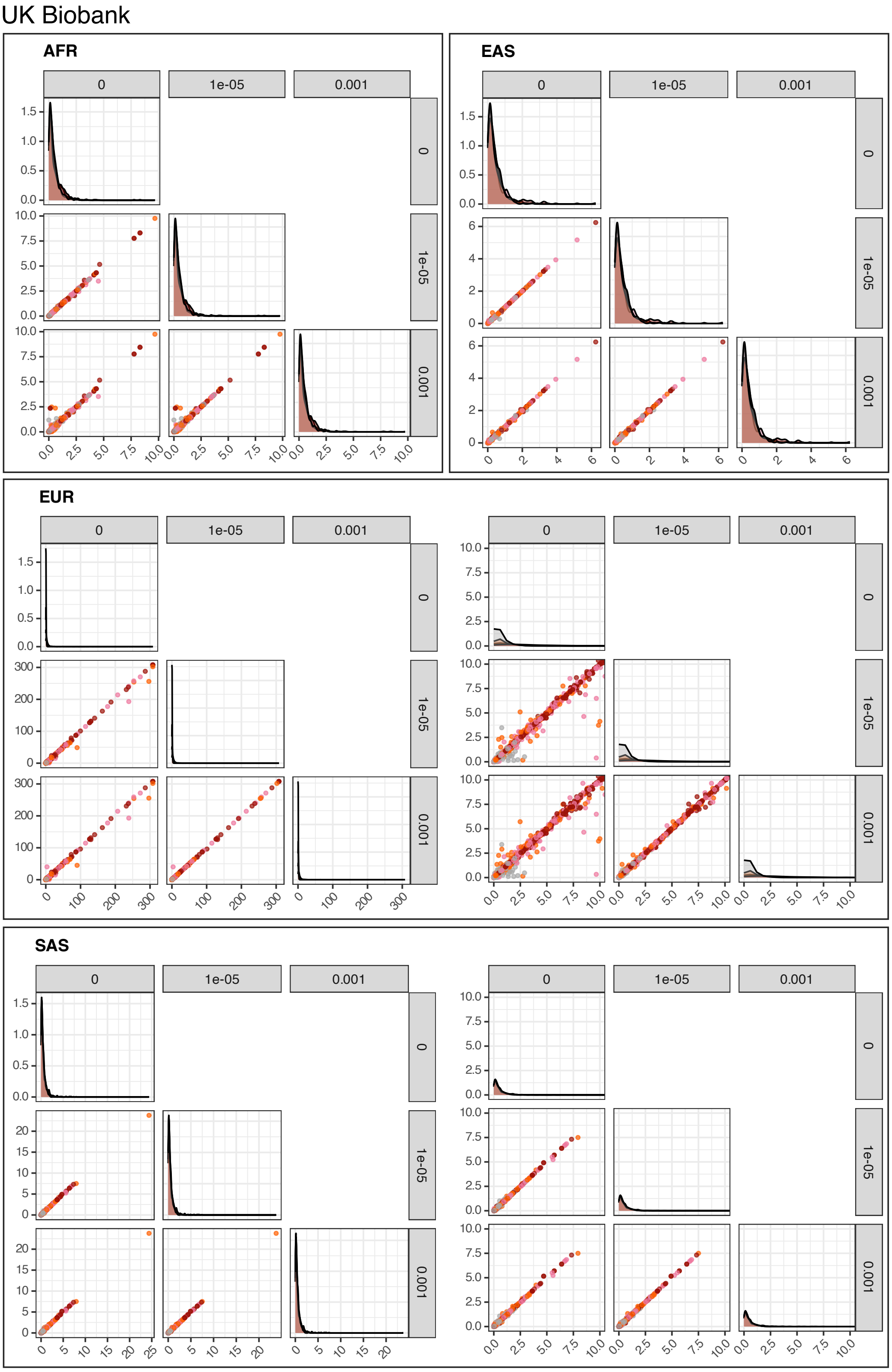

**Figure S43: Examining effect of choice of conditioning $P$-value threshold on results.** $-\log_{10}(P)$ values are plotted against each other after performing iterative conditioning for two $P$-value thresholds $(0.001, 1 \times 10^{-5})$, and excluding conditioning ($P$-value threshold of 0). Histograms on the diagonal show the distribution of association $P$-values for each $P$-value threshold. Dots are colored by annotation according to the legend. For (biobank, ancestry) pairs with at least one association $P < 1 \times 10^{-5}$ we zoom in on the $[0, 10] \times [0, 10]$ region of the plot on the left. Boxes delineate genetic ancestry subsets of the data.

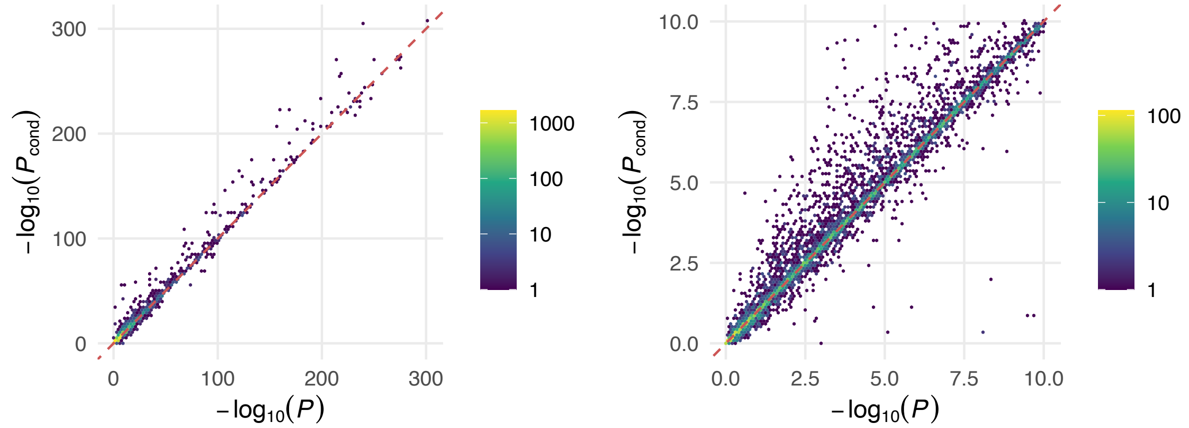

**Figure S44: Comparison of mask-level association $P$-values before and after conditioning on nearby common variation.** $-\log_{10}(P)$ values from the primary association analysis are compared against corresponding $-\log_{10}(P_{cond})$ values obtained after conditioning on nearby common variants. Points are binned and colored according to the number of points within each bin, as indicated by the legend. The right plot zooms into the $[0, 10] \times [0, 10]$ region of the plot on the left.

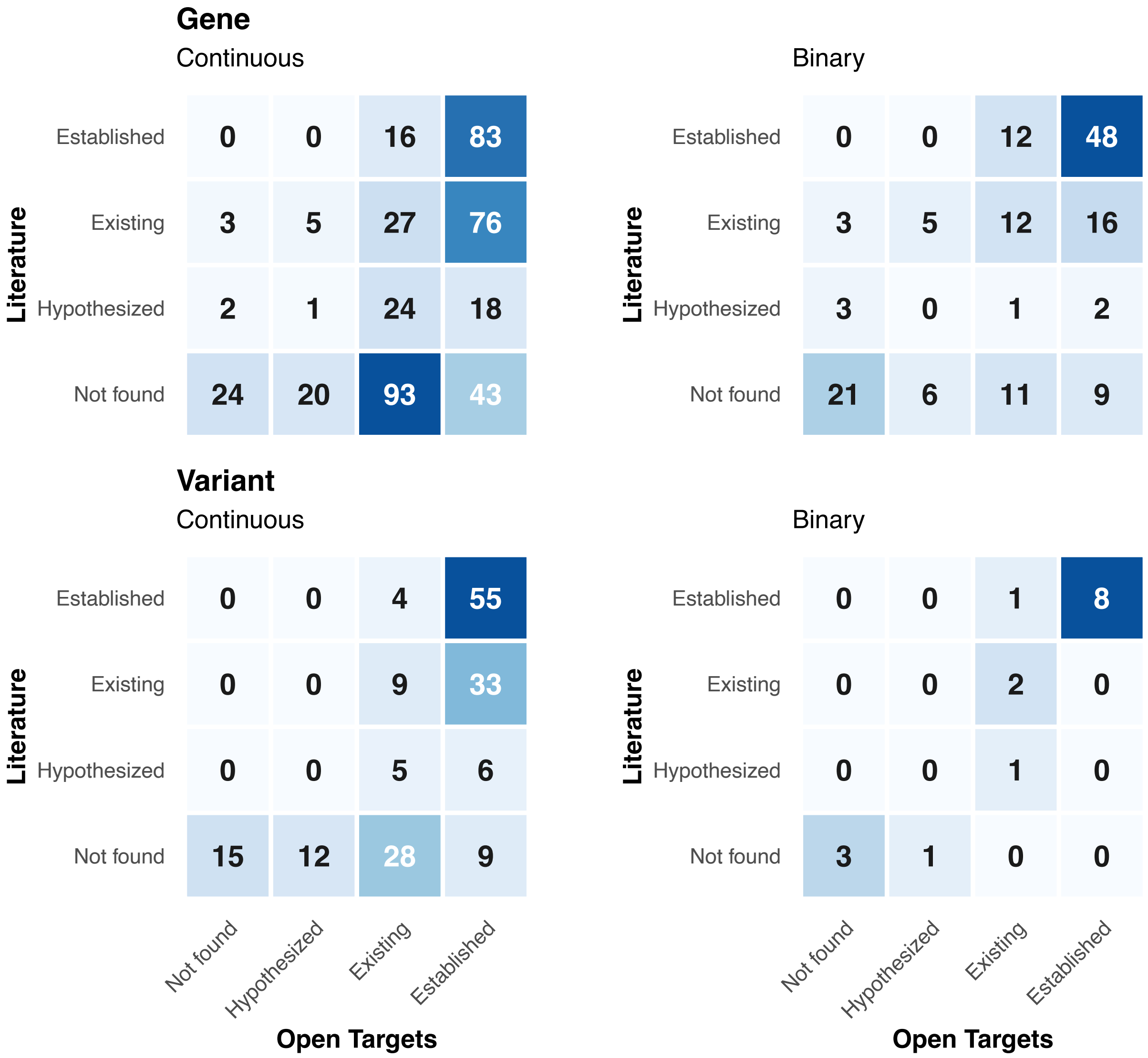

**Figure S45: Confusion matrices determined using AI-based literature curation agent.** The confusion matrices compare assessments using the abstract literature search against those determined from a search of Open Targets[28] for all unique gene-trait associations found to be experiment-wise significant in at least one meta-analysis (ALL, AFR, AMR, EAS, EUR, SAS, non-EUR). The first row display results for the gene-level tests for continuous and binary traits, respectively. The second row shows results for experiment-wise significant variant-level results.

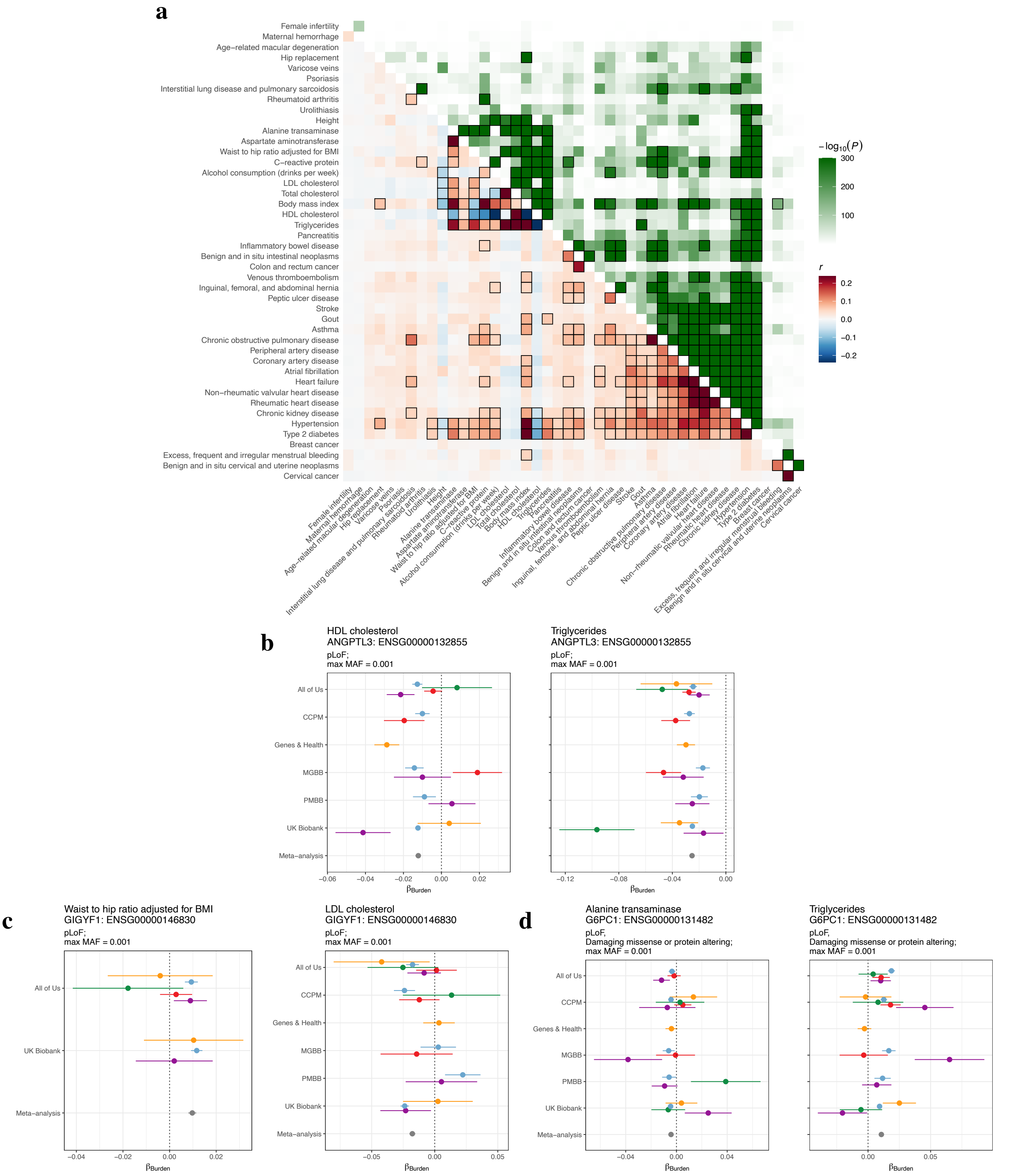

**Figure S46: Phenotypic correlations and discordant gene-level effects across traits.** a, Pairwise correlation matrix across quantitative traits and disease outcomes. For each trait pair, the lower triangle shows Pearson correlation coefficients ($r$), colored by direction and magnitude (red, positive; blue, negative, according to the legend - we clip the scale, such that 98% of correlations lie within the scale in the legend, any correlations exceeding the scale are colored according to the maximum [or minimum] of the scale), while the upper triangle shows corresponding $-\log_{10}(P)$. Squares are outlined when correlations pass Bonferroni significance and have $|r| > 0.05$. Phenotypic correlations were calculated using residuals after regressing out covariates: $age, sex, age \times sex, age^2, age^2 \times sex$, and 10 PCs for traits measured in both sexes, and $age, age^2$, and 10 PCs for sex-specific traits (breast cancer, female infertility, excess or frequent menstrual bleeding, maternal hemorrhage, cervical cancer, and benign cervical or uterine neoplasms). For lipid traits, individuals on statin therapy were excluded prior to evaluation of correlation. b-d, Forest plots illustrating examples where gene-level burden effects are not concordant with phenotypic correlations between traits. Points represent effect estimates ($\beta_{\text{Burden}}$) with standard errors across multiple cohorts (labeled on the $y$-axes, and colored by genetic ancestry according to the legend, with the result following meta-analysis colored in gray. b, *GIGYF1* (pLoF variants; MAF < 0.1%) across waist-to-hip ratio adjusted for BMI and LDL cholesterol. c, *ANGPTL3* (pLoF variants; MAF < 0.1%) for HDL cholesterol and triglycerides. d, *G6PC1* (pLoF and damaging missense or protein-altering variants; MAF < 0.1%) for alanine transaminase and triglycerides.

1601 # Supplementary Tables

**Table S1: Pilot phenotypes, binary.** Phenotype definitions are included based on ICD case inclusion and control exclusion criteria for both ICD9 and ICD10 through comma delimited ICD codes, and mapped to SNOMED codes. Regular expressions are then defined for ICD codes, taking advantage of the inherent tree structure of the ICD encodings. Phenotype definitions were defined using a combination of phecodes (shown in the Phecode column), mappings provided by GHDx, and definitions put forward by analysts in the consortium. Phenotypes for which samples were restricted to a single sex are detailed in the 'sex' column.

**Table S2: Pilot phenotypes, continuous.** ICD based phenotype exclusion criteria are provided for both ICD9 and ICD10 through comma delimited ICD codes. Regular expressions are then defined for ICD codes, taking advantage of the inherent tree structure of the ICD encodings. Prior to analysis, all continuous phenotypes are inverse rank normalized.

**Table S3: Biobank summaries.** Constituent members of BRaVa are listed, together with approximate number of individuals with paired genetic sequencing (exome or genome) and phenotypic data, location of sampling, and sampling strategy of the biobank/cohort.

**Table S4: Sample sizes by (phenotype, genetic ancestry, biobank); binary traits.** For each of the 33 clinical endpoints, we show the number of cases and controls, split by ancestry, for each of the contributing biobanks. Note that sample sizes are only shown for those with at least 100 cases, the cutoff we enforce for inclusion of test statistics within the meta-analysis.

**Table S5: Sample sizes by (phenotype, genetic ancestry, biobank); continuous traits.** For each of the 11 continuous traits, we show the count of individuals with phenotype and sequence data available, split by ancestry, for each of the contributing biobanks.

**Table S6: Sample size summaries for binary phenotypes.** Sums of total case and control counts across (biobank, ancestry) pairs for each binary phenotype. We also display the number of unique biobanks and genetic ancestries represented in the downstream meta-analysis.

**Table S7: Sample size summaries for continuous phenotypes.** Sums of sample counts across (biobank, ancestry) pairs for each continuous phenotype, together with counts of the number of unique biobanks and genetic ancestries represented in the downstream meta-analysis for each trait.

**Table S8: Sample sizes by genetic ancestry group.** For each (biobank, genetic ancestry) pair, we provide a count of the maximum number of available samples for association testing.

**Table S9: Disease prevalence of analyzed traits by biobank.** For each (biobank, trait) pair, we provide and control counts, and associated prevalence within the cohort.

**Table S10: Genomic inflation factors from gene-based association tests of synonymous variation.** Each row corresponds to a specific combination of genetic ancestry, biobank, phenotype, and MAF threshold. Columns report the ancestry group, data source, phenotype, maximum variant MAF included in the gene-based test, and the upper-tail genomic inflation metric evaluated (e.g. $\lambda_{95}$), together with its estimated value. All results are based on gene-based association tests of rare synonymous variation.

**Table S11: Results of Deming regressions comparing effect size estimates across broad continental genetic ancestries.** To evaluate the consistency and transferability of genetic associations across diverse populations, we performed Deming regression on the estimated burden effect sizes between pairs of continental genetic ancestries. The table provides the slope and intercept estimates, along with their respective standard errors and $P$-values, for these cross-ancestry comparisons. Results are systematically stratified by trait type, variant filtering criteria (masks), and maximum MAF thresholds.

**Table S12: Experiment-wise significant associations for the analyzed traits at the gene-level: continuous traits.** For each (gene, phenotype) pair with at least one experiment-wise significant association, we provide summaries of experiment-wise significant associations, and existing evidence in the literature, using a combination of an AI agent (methods), and results from gene-bass.

**Table S13: Experiment-wise significant associations for the analyzed traits at the gene-level: binary traits.** For each (gene, phenotype) pair with at least one experiment-wise significant association, we provide summaries of experiment-wise significant associations, and existing evidence in the literature, using a combination of an AI agent (methods), results from gene-bass, and OMIM data.

**Table S14: Detailed meta-analysis association statistics: continuous traits.** Provided in a long (melted) format, this table expands upon the summarized findings by detailing the specific statistical outputs for significant gene-phenotype associations. For each continuous trait, we report the analyzed ancestry group, the specific variant filtering criteria applied (Mask), and the maximum minor allele frequency (max MAF). Additional columns specify the test class (e.g., Burden) and the meta-analysis method used (e.g., Inverse variance), alongside the $P$-value, burden effect size (BETA Burden), and corresponding standard error (SE Burden).

**Table S15: Detailed meta-analysis association statistics.** Provided in a long (melted) format, this table expands upon the summarized findings by detailing the specific statistical outputs for significant associations. For each gene-phenotype pair, we report the analyzed ancestry group, the specific variant filtering criteria applied (Mask), and the maximum minor allele frequency (max MAF). Additional columns specify the test class (e.g., Burden) and the meta-analysis method used (e.g., Inverse variance), alongside the $P$-value, burden effect size (BETA Burden), and corresponding standard error (SE Burden).

**Table S16: Experiment-wise significant associations for the analyzed traits at the variant-level: continuous traits.** For each (gene, phenotype) pair with at least one experiment-wise significant association, we provide summaries of experiment-wise significant associations, and existing evidence in the literature, using a combination of an AI agent (methods), and results from gene-bass.

**Table S17: Experiment-wise significant associations for the analyzed traits at the variant-level: binary traits.** For each (gene, phenotype) pair with at least one experiment-wise significant association, we provide summaries of experiment-wise significant associations, and existing evidence in the literature, using a combination of an AI agent (methods), results from gene-bass, and OMIM data.

**Table S18: Gene-level associations discordant with phenotypic correlations for continuous traits.** This table highlights specific genetic associations where the relationship between the burden effect sizes (BETA Burden) across pairs of continuous traits contradicts their overall phenotypic correlation. Associations were included if the baseline phenotypic correlation between the trait pair was Bonferroni significant $\left(P < \frac{0.05}{(44\cdot 43)/2} = 5.3 \times 10^{-5}\right)$ with an absolute magnitude ($|r| > 0.05$), and the direction of the genetic burden effects was discordant with this phenotypic correlation. For each identified instance, we report the compared phenotypes, the gene involved, variant filtering criteria (Mask and max MAF), the parallel association statistics ($P$-value and BETA Burden) for both traits, and their observed phenotypic correlation.

**Table S19: Logistic regression analysis of enrichment of CES gene sets among BRaVa gene-level associations.** For each gene set, we fit a logistic regression model with gene-level association status (BRaVa hit vs non-hit) as the outcome and gene set membership as the predictor of interest, adjusting for gene length ($\log_{10}$-transformed), evolutionary conservation (mean phyloP across MANE transcript exons), and CRISPR-based gene essentiality (DepMap median gene effect score). Reported values include log-odds coefficients, standard errors, odds ratios, 95% confidence intervals, and two-sided Wald test $P$-values.

### References

1. All of Us Research Program Investigators *et al.* The “All of Us” Research Program. en. *N. Engl. J. Med.* **381,** 668–676 (15 8 2019).

2. *BRaVa data curation github repository* `https://github.com/BRaVa-genetics/BRaVa_curation`.

3. Jacobs, B. M. *et al.* Genetic architecture of routinely acquired blood tests in a British South Asian cohort. en. *Nat. Commun.* **15,** 8929 (16 10 2024).

4. *Pipeline-Standardization* en.

5. Regier, A. A. *et al.* Functional equivalence of genome sequencing analysis pipelines enables harmonized variant calling across human genetics projects. en. *Nat. Commun.* **9,** 4038 (Oct. 2018).

6. Li, H. *bwa: Burrow-Wheeler Aligner for short-read alignment (see minimap2 for long-read alignment)* en.

7. *Index of /vol1/ftp/technical/reference/GRCh38_reference_genome* `http://ftp.1000genomes.ebi.ac.uk/vol1/ftp/technical/reference/GRCh38_reference_genome/`. Accessed: 2025-12-19.

8. The SAM/BAM Format Specification Working Group. *Sequence alignment/map optional fields specification* https://samtools.github.io/hts-specs/SAMtags.pdf. Accessed: 2025-12-19.

9. *picard: A set of command line tools (in Java) for manipulating high-throughput sequencing (HTS) data and formats such as SAM/BAM/CRAM and VCF* en.

10. *Google Cloud console* en. https://console.cloud.google.com/storage/browser/genomics-public-data/resources/broad/hg38/v0/. Accessed: 2025-12-19.

11. *hail 0.2 documentation* https://hail.is/docs/0.2/.

12. Chang, C. C., Chow, C. C., Tellier, L. C., Vattikuti, S., Purcell, S. M. & Lee, J. J. Second-generation PLINK: rising to the challenge of larger and richer datasets. en. *Gigascience* **4,** 7 (Feb. 2015).

13. McLaren, W. *et al.* The Ensembl Variant Effect Predictor. en. *Genome Biol.* **17,** 122 (June 2016).

14. Karczewski, K. J. *et al.* The mutational constraint spectrum quantified from variation in 141,456 humans. en. *Nature* **581,** 434–443 (May 2020).

15. Rentzsch, P., Schubach, M., Shendure, J. & Kircher, M. CADD-Splice-improving genome-wide variant effect prediction using deep learning-derived splice scores. en. *Genome Med.* **13,** 31 (Feb. 2021).

16. Ioannidis, N. M. *et al.* REVEL: An ensemble method for predicting the pathogenicity of rare missense variants. en. *Am. J. Hum. Genet.* **99,** 877–885 (Oct. 2016).

17. Liu, X., Li, C., Mou, C., Dong, Y. & Tu, Y. dbNSFP v4: a comprehensive database of transcript-specific functional predictions and annotations for human nonsynonymous and splice-site SNVs. en. *Genome Med.* **12,** 103 (Feb. 2020).

18. Jaganathan, K. *et al.* Predicting Splicing from Primary Sequence with Deep Learning. en. *Cell* **176,** 535–548.e24 (Jan. 2019).

19. Pejaver, V. *et al.* Calibration of computational tools for missense variant pathogenicity classification and ClinGen recommendations for PP3/BP4 criteria. en. *Am. J. Hum. Genet.* **109,** 2163–2177 (Jan. 2022).

20. *BRaVa variant annotation github repository* http://github.com/BRaVa-genetics/variant-annotation.

21. *universal-saige github repository* https://github.com/BRaVa-genetics/universal-saige.

22. National Genomic Research Library, Genomics England (2024). https://doi.org/10.6084/m9.figshare.4530893.

23. Morales, J. *et al.* A joint NCBI and EMBL-EBI transcript set for clinical genomics and research. en. *Nature* **604,** 310–315 (Apr. 2022).

24. Cunningham, F. *et al.* Ensembl 2022. en. *Nucleic Acids Res.* **50,** D988–D995 (July 2022).

25. Pollard, K. S., Hubisz, M. J., Rosenbloom, K. R. & Siepel, A. Detection of nonneutral substitution rates on mammalian phylogenies. en. *Genome Res.* **20,** 110–121 (Jan. 2010).

26. Arafeh, R., Shibue, T., Dempster, J. M., Hahn, W. C. & Vazquez, F. The present and future of the Cancer Dependency Map. en. *Nat. Rev. Cancer* **25,** 59–73 (Jan. 2025).

27. Jurgens, S. J. *et al.* Rare coding variant analysis for human diseases across biobanks and ancestries. en. *Nat. Genet.* **56,** 1811–1820 (Sept. 2024).

28. Ghoussaini, M. *et al.* Open Targets Genetics: systematic identification of trait-associated genes using large-scale genetics and functional genomics. en. *Nucleic Acids Res.* **49,** D1311–D1320 (Aug. 2021).
